# Multi-organ metabolome biological age implicates cardiometabolic conditions and mortality risk

**DOI:** 10.1101/2025.03.27.25324801

**Authors:** Junhao Wen, Christos Davatzikos, Sarah Ko, Mehrshad Saadatinia, Filippos Anagnostakis, Jingyue Wang, the MULTI consortium

## Abstract

Biological aging clocks across organs and omics data, including clinical phenotypes, neuroimaging, proteomics, and epigenetics, have proven instrumental in advancing our understanding of human aging and disease. Here, we expand this aging clock framework to plasma metabolomics by developing 5 organ-specific metabolome-based biological age gaps (MetBAGs) using 107 plasma non-derived metabolites from 274,247 UK Biobank participants. Our multi-organ MetBAGs were trained using Lasso regression and neural networks, achieving a mean absolute error of approximately 6 years (0.25<*r*<0.42) and comparable with literature for non-organ specific MetBAGs. Critically, including composite metabolites (e.g., sums or ratios of the original metabolites) reduced model generalizability to independent test data due to the high collinearity (or multicollinearity). Genome-wide associations identified 405 MetBAG-locus pairs (P<5×10□□/5). Using SBayesS, we estimated the SNP-based heritability (0.09<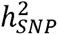<0.18), negative selection signatures (-0.93<*S*<-0.76), and polygenicity (0.001<*Pi*<0.003) for the 5 MetBAGs. Genetic correlation and Mendelian randomization analyses revealed potential causal links between the 5 MetBAGs and cardiometabolic conditions, such as metabolic disorders with the endocrine MetBAG and hypertension with the immune MetBAG. Integrating multi-organ and multi-omics features improved disease category and mortality predictions. The 5 MetBAGs expand upon existing biological aging clocks, providing an enriched framework for studying aging and disease across multiple biological scales.

## Main

Multi-organ biological age gaps (BAG), derived from clinical phenotypes, neuroimaging, and proteomics^1,2,3,4,5,6,7,8,9,10,8,11^ via artificial intelligence and machine learning (AI/ML), have become pivotal in understanding the mechanisms of human aging, disease, and mortality. These aging clocks provide a holistic perspective, capturing functional and structural changes across various organs. Despite the recent advancements, plasma metabolomics – a critical layer of molecular data – remains underexplored in this framework. Unlike proteomics, which focuses on proteins and post-translational modifications, metabolomics examines the downstream products of cellular processes, encompassing the biochemical intermediates and byproducts of metabolic pathways^12,13^.

Developing plasma metabolome-based BAGs is essential for enhancing the granularity and coverage of multi-organ aging clocks, building upon prior research by our group and others ^1,2,3,4,5,6,7,8,9,10,8^. Metabolome-based BAGs may provide new insights and pose unique challenges compared to plasma proteomics and phenotype-based BAGs due to the complex and dynamic nature of the metabolome. Metabolomics data provide a dynamic and highly responsive view of an individual’s biological state. Its sensitivity to factors such as diet, microbiome composition, environmental exposures, and immediate physiological changes makes it a powerful tool for capturing real-time metabolic shifts. Additionally, metabolomics encompasses a diverse array of small molecules, many of which extend beyond direct gene products, offering unique biological insights. While integrating metabolic signatures with genetic and proteomic data requires thoughtful approaches, doing so within a multi-organ framework holds great promise for uncovering novel aging and disease mechanisms. Unlike phenotype-based BAGs, which rely on observable clinical traits or imaging-derived metrics, metabolome-based BAGs capture aging at a deeper molecular level. This offers a more precise and dynamic reflection of an individual’s metabolic state, providing unique insights into aging processes. While translating molecular data into higher-order outcomes like cognitive decline, disease onset, or lifespan prediction requires sophisticated modeling, this depth of information holds great potential for advancing precision aging biomarkers and therapeutic strategies. By integrating metabolomics data, researchers can achieve a more holistic understanding of aging processes, linking molecular pathways to organ-specific aging and systemic health outcomes^14,15,16^. This molecular-level insight is crucial for capturing the metabolic state of an organism and bridging the gap between the causal pathway from underlying genotypes to exo-phenotypes, such as cognitive decline, clinical symptoms, or disease onset.

In our previous studies, we developed 9 phenome-wide BAGs^3,2^ (PhenoBAG) and 11 proteome-wide BAGs (ProtBAG^17^, using a similar approach as in Oh et al.^8^, https://labs-laboratory.com/medicine/hepatic_protbag), representing two essential aspects of human aging and disease causal pathways. Through genome-wide association studies (GWAS) and subsequent analyses, including genetic correlation^18^, polygenic risk score^19^, and causal inference^4^, we thoroughly evaluated the clinical applicability of these AI/ML-derived biomarkers. Here, this study used 107 plasma non-derived metabolites from 274,247 UK Biobank participants (**UKBB**, **Method 1**, and **Supplementary eTable 1**) to develop 5 organ-specific metabolome-based BAG (MetBAG), including the digestive, hepatic, immune, endocrine, and metabolic systems (**Method 2**). Subsequently, we examine their genetic architecture (i.e., SNP-based heritability, natural selection signature, and polygenicity), genetic correlation, and causal relationships with 525 disease endpoints (DEs) from FinnGen^20^ and Psychiatric Genomics Consortium (PGC)^21^ (**Method 3**). Finally, we assessed the clinical promise of the 5 MetBAGs and their PRSs for predicting 14 disease categories and mortality (**Method 4**). All results and pre-trained AI/ML models are publicly disseminated at the MEDICINE portal: https://labs-laboratory.com/medicine/.

## Results

### Biological age prediction performance of the 5 MetBAGs

To rigorously evaluate the performance of biological age prediction models, we partitioned the 34,354 healthy control (CN, without any pathologies) participants into the CN training/validation/test (*N*=29,354) and independent test (ind. test; *N*=5000) datasets (**Method 2e**). **Extended Data Fig. 1** details this study’s population selection and overall workflow. The CN training set was used for model development and nested cross-validation (Lasso regression) for the two AI/ML models (Lasso regression and neural network (NN)), while the independent test set provided an unbiased assessment of model performance (**Supplementary eTable 1**).

When fitting the organ-specific metabolites (**Fig. 1a**, **Method 2b**, and **Supplementary eTable 2**), the two AI/ML models showed marginal variability in model performance, with no single model consistently outperforming the others (**Fig. 1b**). The optimal model for each organ-specific MetBAG was selected based on a higher degree of generalizability (i.e., a smaller Cohen’s D, as denoted by the # symbol in **Fig. 1b**). The optimal models of the 5 MetBAGs showed moderate Pearson’s *r* coefficients (0.25<*r<*0.42) in the independent test dataset (**Fig. 1c**). The metrics after correcting the age bias^22,23^ are shown in **Extended Data Fig. 2**. For all subsequent analyses, we used the age bias-corrected^23^ MetBAGs. **Supplementary eTable 3** presents detailed statistics for the age prediction tasks before and after the age bias correction.

**Figure 1:**
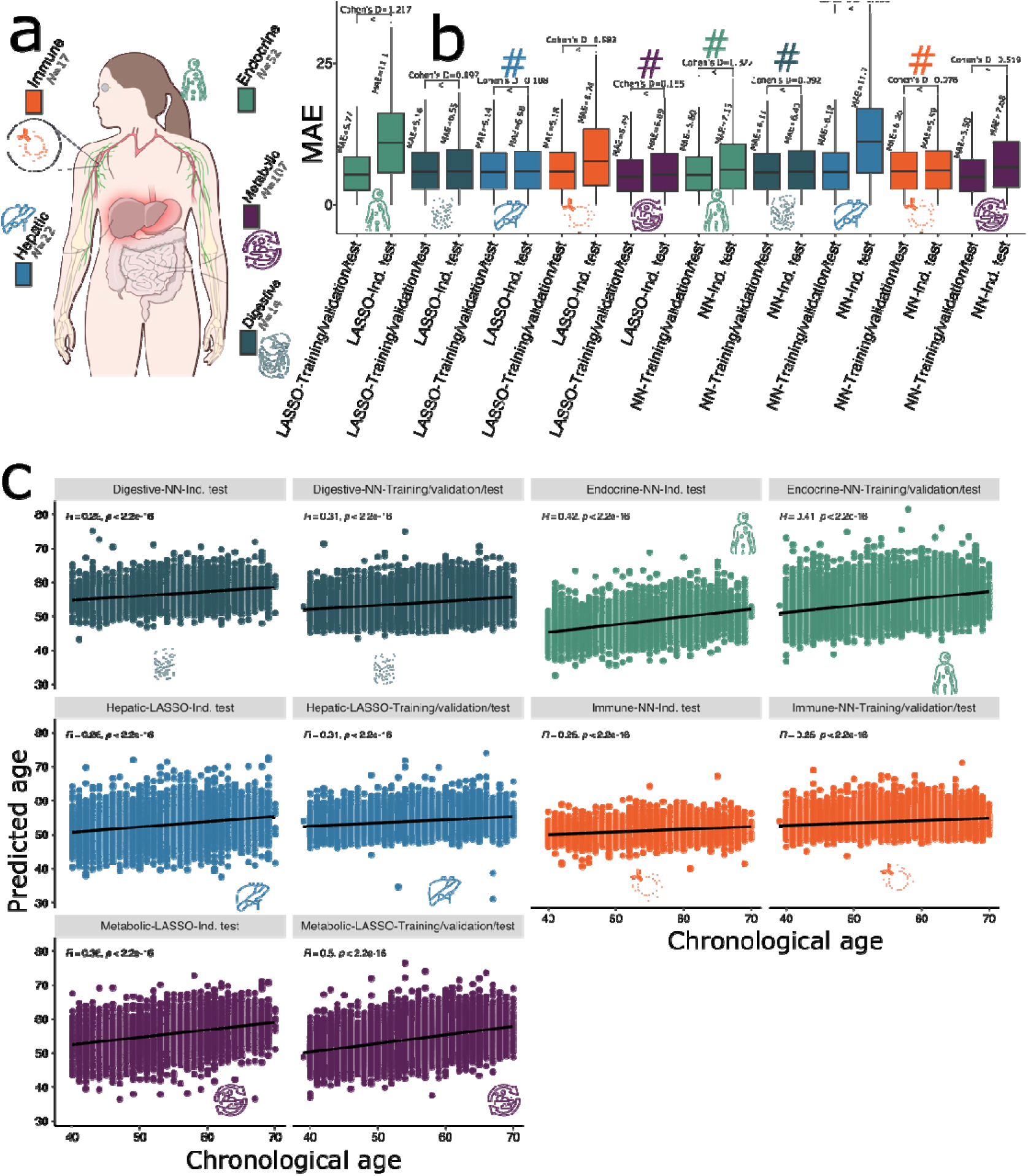
Two AI/ML models to derive the 5 multi-organ MetBAGs. **a**) We identified organ-specific metabolites for the five human organ systems by mapping the 107 non-derived metabolites to their corresponding plasma proteins using a linear regression model. The most strongly associated protein for each metabolite was then linked to their organ-specific RNA expression profiles using data from the Human Protein Atlas (e.g., S_HDL_P→LCAT→liver: https://www.proteinatlas.org/ENSG00000213398-LCAT/tissue). **Method 2b** and **Supplementary eTable 2** detail the metabolite-protein-organ annotations. An interactive graph visualization for this annotation is also accessible at https://labs-laboratory.com/medicine/metabolite_organ_annotation. **b**) We evaluated age prediction performance using the mean absolute error (MAE) on independent test (ind. test) data, employing 2 AI/ML models across 5 organ systems with 107 metabolites. The "#" symbol indicates the model that generalized best to the independent test data (i.e., smallest Cohen’s D). **c**) The scatter plot between the AI/ML-derived biological age and chronological age without applying the age bias correction^23^.

In our previous study^17^, we systematically assessed model performance of key modeling components (e.g., the pathology profile of the training population and the degree of organ specificity of the proteins) from both methodological and clinical perspectives, using 11 ProtBAGs as a demonstration. Here, leveraging nested cross-validation and an independent test dataset, we further investigated factors contributing to overfitting and limited generalizability in developing the 5 MetBAGs. Specifically, we identified: *i*) poor generalizability to independent test datasets in certain models, and *ii*) substantial overfitting and reduced generalizability when incorporating composite metabolite biomarkers derived from sums or ratios of the 107 non-derived metabolites (**Supplementary eFigure 1**). A detailed discussion is presented in **Supplementary eNote 1**. We also discussed sex-stratified analyses via the metabolic MetBAG in **Supplementary eNote 2** and **eFigure 2**.

### The genetic architecture of the 5 MetBAGs

We conducted GWAS (**Method 3a**) for the 5 MetBAGs and identified 405 (P-value<5×10^-8^/5) genomic locus-BAG pairs. We denoted the genomic loci using their top lead SNPs defined by FUMA (**Supplementary eNote 3**) considering linkage disequilibrium (LD); the genomic loci are presented in **Supplementary eTable 4**. We visually present the shared genomic loci annotated by cytogenetic regions based on the GRCh37 cytoband (**Fig. 2a**). Manhattan and QQ plots, as well as the genomic inflation factor (*λ*) of the 5 MetBAG GWASs, are presented in the MEDICINE portal (e.g., the hepatic MetBAG: https://labs-laboratory.com/medicine/hepatic_metbag). The LDSC^18^ intercept (LDSC*_b_*=1.05 [1.03, 1.08]) of the 5 MetBAG GWASs was close to 1, and the LDSC ratio (an indication of inflation vs. true polygenicity; LDSC*_r_*=0.10 [0.09, 0.13])) was small, indicating that no severe inflation due to population stratification was observed. **Extended Data Fig. 3** presents the trumpet plots of the effective allele frequency vs. the *β* coefficients of the 5 MetBAG GWASs.

**Figure 2:**
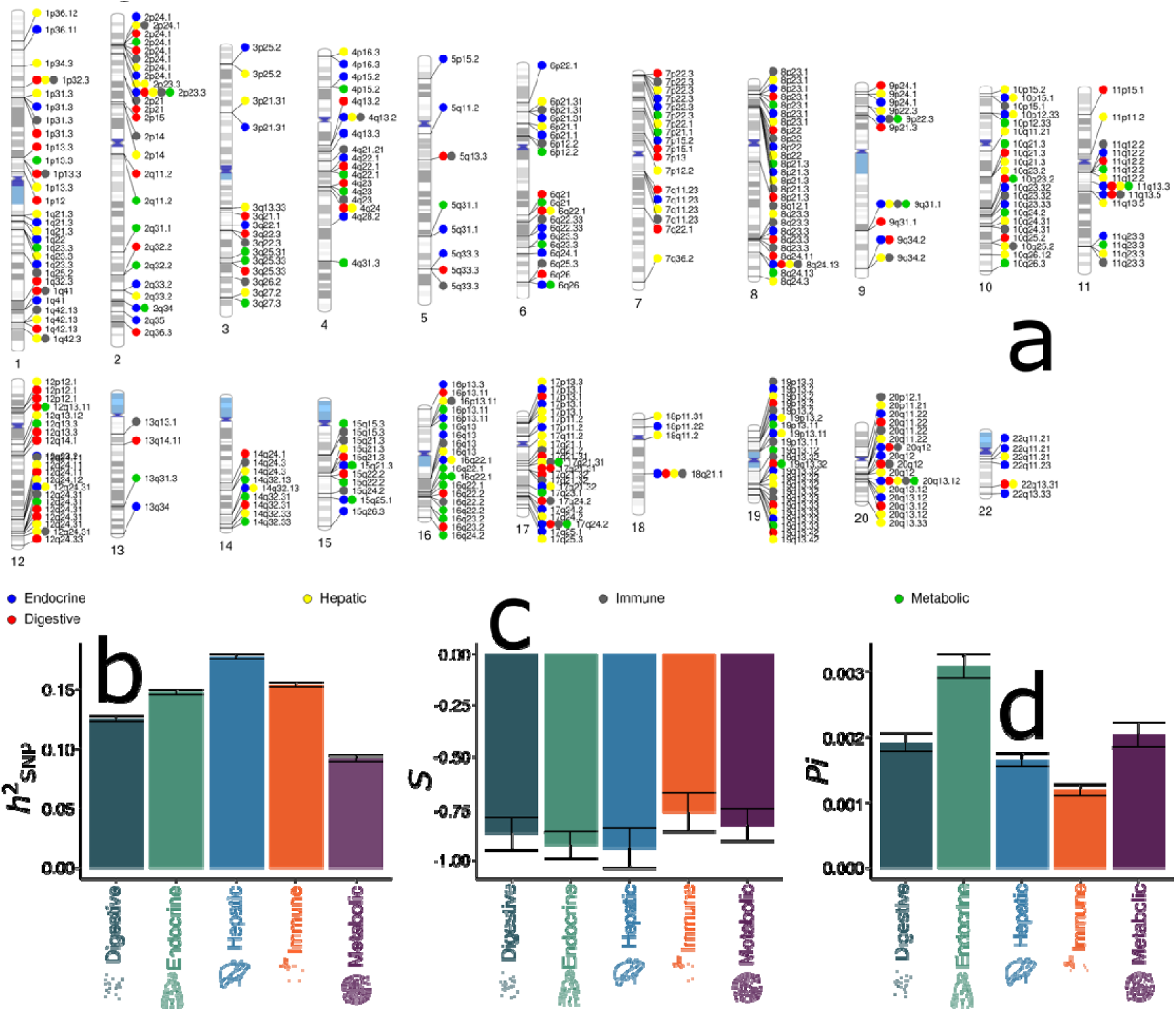
The genetic architecture of the 5 MetBAGs. **a)** Cytogenetic regions where the genomic region was linked to the 5 MetBAGs. Bonferroni correction was applied to denote significant genomic loci associated with PhenoBAG (P-value<5×10^-8^/5). SBayesS estimates three genetic parameters to delineate the genetic architecture of the 5 MetBAGs, including **b)** the SNP-based heritability ( ), **c)** the relationship between MAF and effect size for the selection signature (*S*), and **d)** polygenicity ( ).

We then computed three key genetic parameters of the 5 MetBAGs, including SNP-based heritability estimate (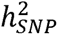), natural selectional signature (*S*), and polygenicity *(Pi*) (**Method 3b**). We observed small to moderate 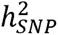, ranging from 0.09 to 0.18 (**Fig. 2b**); a detailed comparison for the 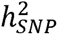 estimate between the 5 MetBAGs, 11 ProtBAGs, and 9 PhenoBAGs are presented in **Supplementary eNote 4** and **eFigure 3**. The 5 MetBAGs showed strong negative selection signatures, exemplified by the hepatic MetBAG (-0.94±0.09) (**Fig. 2c**). The highest polygenic estimate was obtained by the endocrine MetBAG (0.003±0.0002) (**Fig. 2d**). **Supplementary eTable 5** presents detailed statistics.

### The genetic correlation between the 5 MetBAGs, 107 non-derived metabolites, 9 PhenoBAGs, 11 ProtBAGs, and 525 DEs

We used LDSC to estimate the genetic correlation (*g_c_*) of the 5 MetBAGs with three sets of traits (**Method 3c**). We observed strong genetic correlations among the 5 MetBAGs (0.17 <*g_c_*< 0.78), with the highest correlation between the immune and hepatic MetBAGs (*g_c_*=0.78±0.04). Additionally, the phenotypic correlations (*p_c_*) among the 5 MetBAGs closely reflected the genetic correlations (**Fig. 3a**). Subsequently, we also observed strong genetic correlations between the 5 MetBAGs and the 107 non-derived metabolites (-0.44<*g_c_*<0.91; P-value<0.05/327) (**Fig. 3b**). We presented two cases where the genetic correlations between organ MetBAGs and metabolites either align or do not align with the metabolite-protein-organ annotations (https://labs-laboratory.com/medicine/metabolite_organ_annotation). For example, we found a negative genetic correlation between the gly metabolite and the endocrine BAG (*g_c_*=-0.20±0.03); our metabolite-protein-organ annotation indeed assigned the gly metabolite to the endocrine system (gly→LPL→endocrine) (**Fig. 3c**). Another example is the acetoacetate metabolite, which was annotated to the endocrine system (**Fig. 3c**) but showed a positive association with the hepatic MetBAG (*g_c_*=0.24±0.06). Using RNA-seq data from the Human Protein Atlas (HPA), we found that the ANGPTL4 protein was also highly expressed in liver tissue (nTPM = 179.2), slightly lower than its expression in adipose tissue of the endocrine system (nTPM = 189.6). Finally, we also found 18 significant genetic associations (P-value<0.05/545) between the 5 MetBAGs and 545 traits, including 9 PhenoBAGs^3^, 11 ProtBAGs, and 525 DEs. For instance, the endocrine MetBAG was associated with the metabolic and hepatic PhenoBAGs, the hepatic ProtBAG, and several cardiometabolic conditions, such as metabolic diseases (FinnGen code: E4_METABOLIA) and hypercholesterolemia (E4_HYPERCHOL). The immune MetBAG was connected to various types of hypertension (**Fig. 3d**). **Supplementary eTable 6** presents detailed statistics.

**Figure 3:**
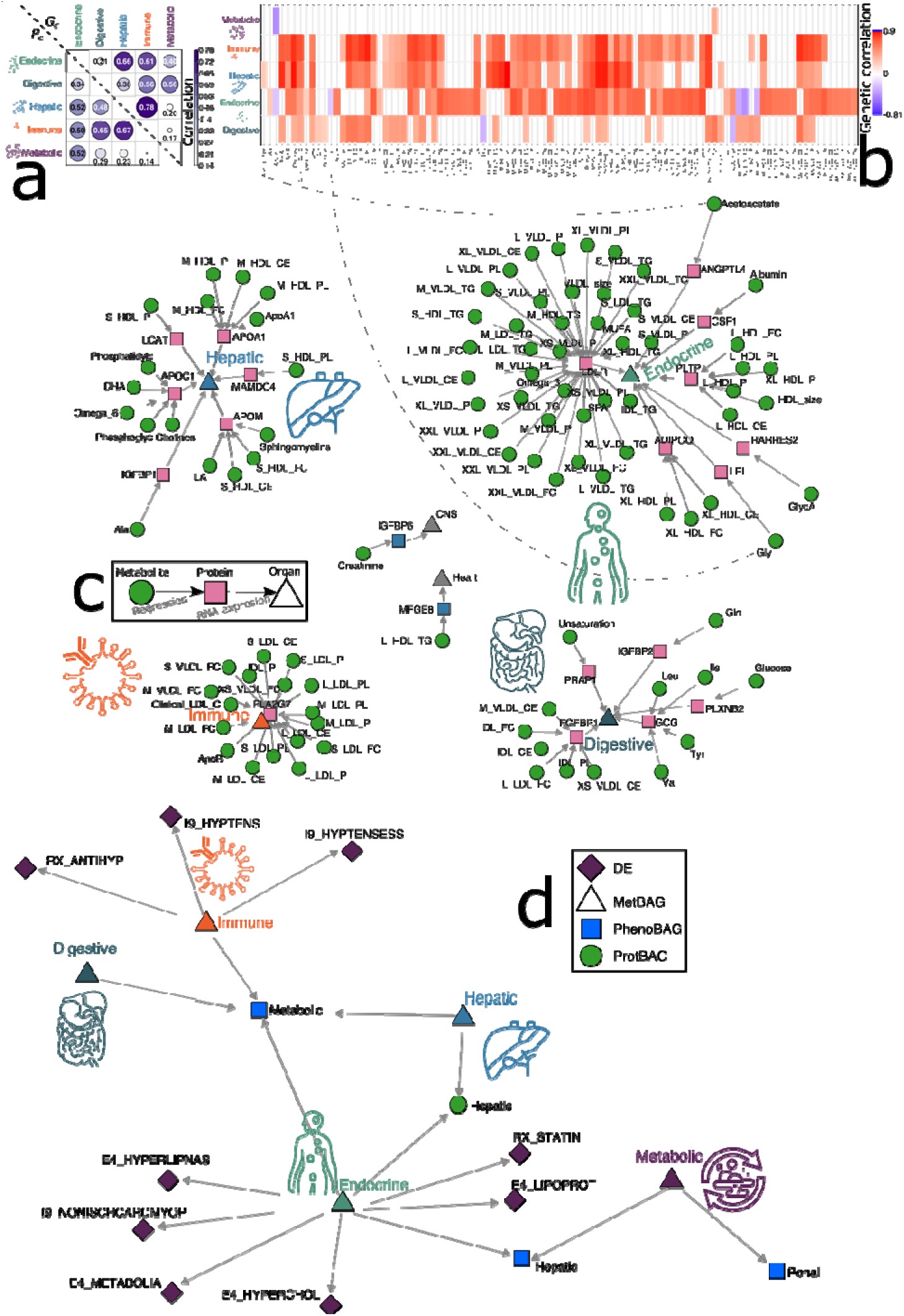
Genetic correlation between the 5 MetBAGs, 9 PhenoBAGs, 11 ProtBAGs, 107 metabolites, and 525 DEs. **a**) Genetic and phenotypic correlation between the 5 MetBAGs. **b**) Genetic correlation between the 5 MetBAGs and the 107 non-derived NMR metabolites. Bonferroni correction (P-value<0.05/327) was applied to denote the significant metabolites. **c**) The metabolite-protein-organ annotations for the significant metabolites identified in **b**). We exemplified cases where the MetBAG-metabolite genetic correlation partially supports this organ-specificity annotation (e.g., gly→LPL→endocrine). **d**) Genetic correlation between the 5 MetBAGs, 11 ProtBAGs, 9 PhenoBAGs^3^, and 525 DEs. Bonferroni correction (P-value<0.05/545) was applied to denote statistical significance. An interactive network visualization is also available at https://labs-laboratory.com/medicine/metbag_gc.

### The causal relationship between the 5 MetBAGs and 525 DEs

Using bi-directional, two-sample Mendelian randomization (**Method 3d**), we subsequently established two causal networks that linked the 5 MetBAG and 525 DEs (**Fig. 4**). The *MetBAG2DE* network found 27 significant causal relationships (P-value<0.05/500). As anticipated, 4 MetBAGs (immune, hepatic, digestive, and endocrine) were predominantly causally associated with metabolic diseases (FinnGen codes beginning with E4), including the endocrine MetBAG and metabolic diseases (E4_METABOLIA) [P-value= 1.39×10^-8^; OR (95% CI)=1.30 (1.18, 1.41); number of IVs=90], which aligns with the genetic correlation observed in **Fig. 3d**. In addition, cardiovascular diseases (FinnGen codes start with I9) were also largely causally linked with the MetBAGs. For example, the digestive MetBAG was causally linked to heart valve disease (I9_VHD) [P-value= 6.00×10^-6^; OR (95% CI)=1.25 (1.13, 1.37); number of IVs=79].

**Figure 4:**
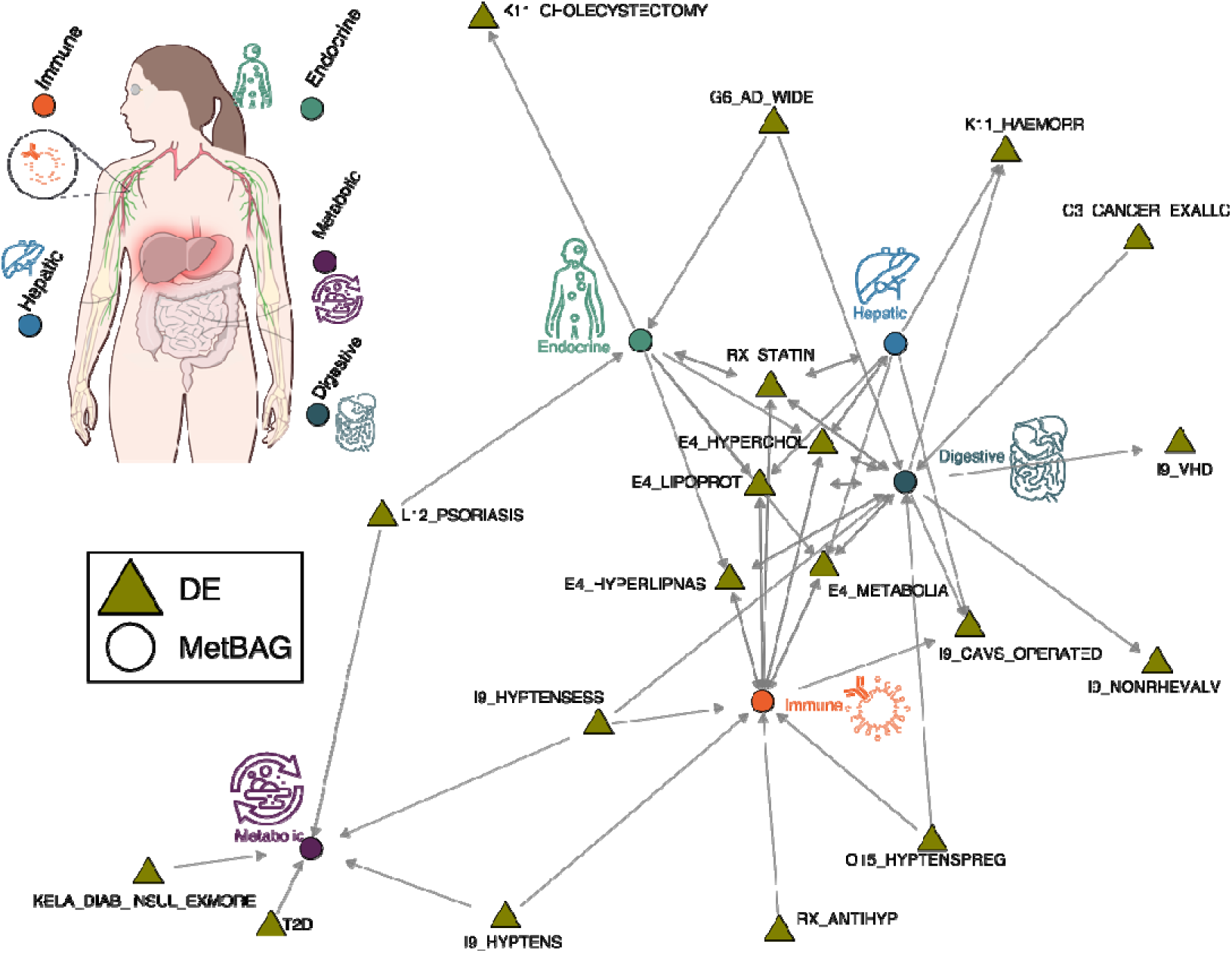
Causal networks between the 5 MetBAGs and 525 DEs. We constructed a causal network by employing bi-directional two-sample Mendelian randomization, following a rigorous quality control procedure to select exposure and instrumental variables (number of IVs>7), corrected for multiple comparisons (based on the number of DEs), and performed sensitivity analyses (e.g., horizontal pleiotropy) to scrutinize the robustness of our results. Two causal networks were analyzed: i) *MetBAG2DE* and ii) *DE2MetBAG*. The arrows indicate the direction of the established causal relationship from the exposure variable to the outcome variable. An interactive network visualization is also available at https://labs-laboratory.com/medicine/metbag_mr. It is crucial to approach the interpretation of these potential causal relationships with caution despite our thorough efforts in conducting multiple sensitivity checks to assess any potential violations of underlying assumptions.

For the *DE2MetBAG* causal network, we found 28 significant signals (P-value<0.05/179). The involvement of cardiometabolic diseases persisted, exemplified by the causal relationship from hypertension (I9_HYPTENS) to the immune MetBAG and from disorders of lipoprotein metabolism and other lipidaemias (E4_LIPOPROT) to the digestive MetBAG. In addition, Alzheimer’s disease was causally linked to the digestive [P-value= 2.16×10^-10^; OR (95% CI)=1.08 (1.05, 1.10); number of IVs=8] and endocrine [P-value= 1.12×10^-6^; OR (95% CI)= 1.04 (1.03, 1.05); number of IVs=8] MetBAGs (**Fig. 4**).

Mendelian randomization relies on stringent assumptions that can sometimes be violated. We conducted comprehensive sensitivity analyses for the significant signals identified to scrutinize this. **Extended Data Fig. 4** provides an example of our sensitive check analyses, with a detailed discussion in **Supplementary eNote 5**. Detailed statistics for all five estimators are presented in **Supplementary eTable 7**, and the results of the sensitivity analyses are presented in **Supplementary eFolder 1**.

### The clinical promise of the 5 MetBAGs and 5 MetBAG-PRSs

We demonstrate the clinical promise of the 5 MetBAGs and 5 MetBAG-PRSs in predicting various clinical outcomes through binary classification and survival analysis: *i*) the classification of 14 systemic disease categories and *ii*) the risk of mortality (**Method 4a-b**).

We evaluated the predictive ability of a neural network (NN) at the individual level to classify 14 disease categories (**Method 4a**). Due to the large sample size, we selected the NN over support vector machines (SVM), as SVM does not scale well with large sample-sized datasets. We report the balanced accuracy (BA) from the independent test dataset (*N*=10,000), with additional metrics and results from the training/test datasets provided in **Supplementary eTable 8**. The MetBAG-PRS alone did not outperform random chance (BA<0.5). However, combining the MetBAGs with their PRS resulted in a slight increase in classification accuracy, as illustrated by the genitourinary disease category (ICD codes: N) and circulative disease category (ICD codes: I) (**Fig. 5a**). Adding age and sex further enhanced the classification accuracy (**Supplementary eFigure 4**). **Supplementary eFigure 5** shows the loss of training and validation (at the end of each epoch) during the model training.

**Figure 5:**
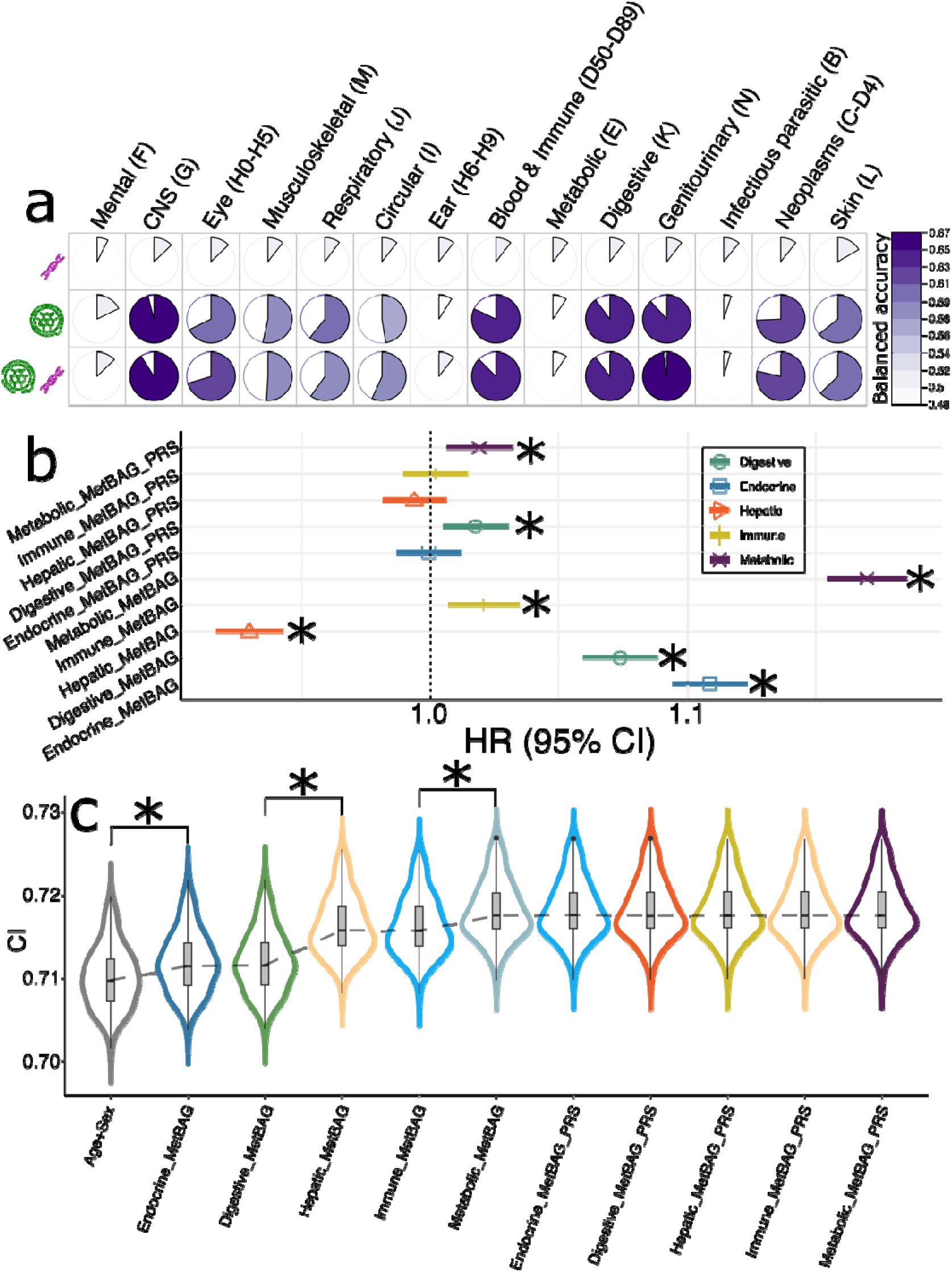
MetBAG and their PRS predict systemic disease categories and mortality. **a)** A neural network evaluated the classification balanced accuracy (BA) for 14 ICD-based disease categories using the 5 MetBAGs and MetBAG-PRSs as features. Balanced accuracy (BA) from the independent test dataset (*N*=10,000) is presented, with additional metrics provided in the Supplement and training/test dataset results. **b**) The 5 MetBAGs and MetBAG-PRSs show significant associations with the risk of mortality. Age and sex were included as covariates in the Cox proportional hazard model. The symbol * indicates significant results that survived the Bonferroni correction (<0.05/5). **c**) The 5 MetBAGs and MetBAG-PRSs were cumulatively included as features for mortality risk prediction. The * symbol indicates statistical significance (<0.05) from a two-sample t-test comparing results between two Cox models. HR: hazard ratio; CI: concordance index.

We also used the 5 MetBAGs and their PRSs to predict mortality risk (**Method 4b**). Our analysis revealed that all the 5 MetBAGs, digestive MetBAG-PRS and metabolic MetBAG-PRS, showed significant associations (P-value<0.05/5) with mortality. The metabolic MetBAG showed the highest mortality risks [HR (95% CI)=1.16 (1.15, 1.18); P-value=1.00×10^-116^], followed by the endocrine MetBAG [HR (95% CI)=1.11 (1.09, 1.12); P-value=6.77×10^-53^]. The hepatic MetBAG showed a protective association with mortality [HR (95% CI)=0.93 (0.92, 0.94); P-value=3.23×10^-24^] (**Fig. 5b**). We then conducted a cumulative prediction analysis based on the substantial associations. This analysis demonstrated that combining these features provided additional predictive power beyond age and sex, achieving an average concordance index of 0.72 (**Fig. 5c**). Comprehensive statistics, including HRs, P-values, and sample sizes, are available in **Supplementary eTable 9**. We also assessed the predictive performance of the 5 MetBAGs in predicting the incidence of ICD-based single disease entities (**Supplementary eFigure 6** and **eTable 10**, and **Method 4c**).

## Discussion

This study explores the clinical potential of 5 multi-organ metabolome-based biological aging clocks, extending previously established aging clocks derived from clinical phenotypes, neuroimaging, and proteomics data. The 5 MetBAGs were developed for 5 key organ systems – immune, endocrine, digestive, hepatic, and metabolic – reflecting the systemic nature of plasma metabolites circulating throughout the body. Given their integral role in these interconnected physiological systems, the 5 MetBAGs were strongly linked to cardiometabolic conditions. Comprehensive genetic analyses illuminated the underlying genetic architecture of these AI/ML-derived aging clocks, uncovering strong genetic correlations and causal relationships that tie each MetBAG to specific cardiometabolic diseases. Furthermore, integrating cross-organ and cross-omics features significantly enhanced the predictive power for 14 systemic disease categories and mortality, underscoring their potential in clinical applications.

Biological age prediction based on plasma metabolites demonstrated comparable performance to previous models using neuroimaging (MAE=4.92–7.95 years with multi-modal brain MRI features^2^), plasma proteomics (MAE=4.33–10.19 years across 11 organ systems), and more broadly-defined phenotype-based aging clocks^11^ (MAE∼5 years). Neuroimaging captures structural and functional brain alterations tied closely to neural aging, making it particularly suited for evaluating central nervous system-related aging processes, such as the brain BAG^24^. Plasma proteomics offers a window into the dynamic protein landscape and post-translational modifications, reflecting organ-specific aging and systemic responses such as inflammation and metabolic dysregulation^25^. In contrast, plasma metabolites represent small-molecule intermediates and end products of metabolic pathways, offering maybe a more direct link to cellular metabolism and cross-system integration^26^, supported by the higher cross-organ correlations (Fig. 3a) than our previously proposed PhenoBAG^3,2^ and ProtBAGs. The ability of plasma metabolites to predict biological age suggests their potential to serve as accessible and cost-effective biomarkers while complementing proteomics and neuroimaging by providing molecular-level insights into systemic metabolic health and disease processes. Integrating these modalities could yield a more comprehensive understanding of aging, accounting for system-specific and systemic processes.

This study provides a comprehensive characterization of the genetic architecture of the 5 MetBAGs through three key genetic parameters. The moderate SNP-based heritability (0.09<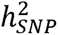 <0.18) estimates indicate that genetic factors significantly contribute to metabolome-based biological aging variation. This underscores the genetic basis of metabolic aging and highlights the influence of inherited genomic variation on age-related metabolic profiles. Notably, we used SBayesS, which relies solely on GWAS summary statistics and an LD reference panel to estimate heritability. Previous studies have demonstrated that this method produces lower heritability estimates than approaches using individual-level data, such as GCTA^27^ ; these estimates from different methods showed high correlations^28^. Strong negative selection signatures indicate that genetic variants influencing MetBAGs are under evolutionary pressure to be less deleterious, consistent with previous estimates^29^ in various complex human traits. These signatures highlight the evolutionary importance of maintaining stable metabolic-related processes to ensure survival and reproduction, suggesting that maladaptive variants are likely to be eliminated over generations. This also implies that genetic variants associated with unfavorable metabolic aging profiles may contribute to disease susceptibility, particularly in conditions where metabolic dysregulation is a key driver (e.g., cardiometabolic diseases^30^). Finally, the observed high polygenicity suggests that many genetic variants with small effects collectively shape the metabolic aging process. This supports the complex, multi-genic nature of metabolic regulation and aging and aligns with the involvement of diverse metabolic pathways, reflecting the systemic integration of organ-specific aging processes influenced by genetics^13,26,31,32^. A previous study^33^ analyzes 325 NMR biomarkers in 250,341 UKBB participants, identifying 54 aging-related biomarkers linked to all-cause mortality. Using GWAS and Mendelian randomization, the researchers uncover 439 potential causal biomarker-disease relationships. They develop a metabolomic aging/mortality score that improves mortality risk prediction and identify candidate pro-aging and anti-aging biomarkers, highlighting the potential of metabolomics for personalized aging monitoring and early disease intervention.

We further depicted the genetic relationships between the 5 MetBAGs, 20 other omics-based BAGs, and 525 DEs. Consistent with expectations, we identified strong and concordant associations between the 5 MetBAGs and DEs linked to cardiometabolic conditions. This alignment was evident across genetic correlation (Fig. 3d) and Mendelian randomization (Fig. 4) analyses, underscoring the shared genetic and causal pathways. The observed genetic correlations highlight overlapping polygenic architectures, where common genetic variants simultaneously influence metabolomic profiles^13,26,31,32^ and cardiometabolic diseases^30,34,35^. Mendelian randomization further supports this link by providing evidence of causality, showing that metabolomic changes directly lead to disease risk and likely vice versa. These findings are likely rooted in the central role of plasma metabolites in systemic physiological processes, such as lipid metabolism, glucose regulation, and inflammation, which are foundational to cardiometabolic health. Interestingly, the strong associations between metabolites and cardiometabolic conditions were also observed in black individuals, where integrative omics approaches harnessed genomic diversity to uncover novel insights into the complexities of cardiometabolic diseases^36^.

Finally, we showcased the clinical potential of the 5 MetBAGs through two prediction tasks, highlighting the value of a holistic, integrative approach that combines multi-omics and multi-organ features to predict disease diagnoses, prognoses, and mortality risks. While classification performance across 14 systemic disease categories was modest, reflecting the heterogeneity of disease groups and data, the genitourinary disease category emerged with the highest predictive accuracy. This outcome may stem from the central role of the genitourinary system, including the kidneys and urinary tract, in metabolite filtration, excretion, and homeostasis. Metabolomics provides a direct readout of systemic metabolites interacting with these processes, making it particularly sensitive to genitourinary health and aging^37^. Furthermore, genitourinary diseases frequently involve metabolic dysregulation, such as in diabetes-related nephropathy or urinary tract dysfunction, which aligns closely with plasma metabolomics^38,39^. The metabolome’s dynamic nature, reflecting both physiological and pathological states, further enhances its capacity to capture relevant biomarkers of genitourinary conditions, including renal function indicators like creatinine, urea, and urinary metabolites. A notable finding is the unexpected negative association between the hepatic MetBAG and all-cause mortality risk (Fig. 5b). The protective association between hepatic MetBAG and mortality may stem from several factors. First, the hepatic MetBAG captures not only liver aging but also adaptive metabolic responses that may enhance resilience and longevity. Supporting this, a recent metabolomics aging study^40^ has shown that several hepatic-specific metabolites defined in our study to derive the hepatic MetBAG, such as DHA and ApoA1 (https://labs-laboratory.com/medicine/metabolite_organ_annotation), are negatively associated with mortality. Second, selection bias in the UK Biobank could contribute, as individuals with severe liver dysfunction may be underrepresented due to survival bias. Third, hepatic MetBAG likely reflects broader systemic metabolic profiles, where some aging-related shifts do not necessarily increase mortality risk. Similarly, findings from a recent proteome-based aging clock study^41^ indicate that some younger biological age profiles in specific organs (e.g., artery) can paradoxically correlate with increased mortality. Collectively, these findings underscore the value of MetBAGs as integrative tools for linking molecular markers of aging to the intricate mechanisms underlying complex diseases and offer AI/ML-derived biomarkers with potential applications in disease monitoring and therapeutic development.

More broadly speaking, the 5 MetBAGs expand our previously proposed aging clcoks based on clinical phenotypes^3^, neuroimaging^2^, and plasma proteomics^17^ data. Recently, Ikram^42^ suggested that biological aging clocks are flawed concepts in understanding disease. The author challenges the assumption that aging itself is a direct cause of disease and instead suggests that aging (whether measured by calendar age or biological aging) is better understood as a summary measure of accumulated changes over time (i.e., a snapshot within the passage of time). While the author emphasizes that biological aging clocks should be used as a propensity score like calendar age as an additional confounder, but itself not as a causal factor. Subsequently, Ferrucci et al.^43^ argues that biological aging is not merely a redundant or supplementary measure to calendar age but provides unique insights into aging and disease risk. While Ikram suggests that biological age biomarkers should only be used as covariates alongside chronological age – implying that they do not capture fundamentally distinct information – Ferrucci counters this by asserting that aging is more than just the passage of time. Here, we directly provided support evidence to demonstrate the additional value of these multi-organ and multi-omics aging clocks, on top of chronological age, for predicting mortality and systemic diseases (**Extended Data Figure 5**). As shown, each individual BAG among the 23 multi-organ and multi-omics aging clocks contributed only marginally to mortality prediction. The only significant association after Bonferroni correction was observed for the Immune PhenoBAG. However, when all 23 BAGs were combined, they explained an additional 1.52% variance in all-cause mortality. This is expected, as other factors such as genetics, environmental influences, and lifestyle play major roles in mortality. For systemic disease categories, combining all 23 aging clocks accounted for an additional 6.45% variance, beyond chronological age and sex (9.85%). These findings support the use of AI/ML-derived biological aging clocks as clinically relevant biomarkers to enhance our understanding of human aging and disease, with potential implications for developing therapies targeting aging-related conditions.

### Limitation & Outlook

Multi-organ biological aging clocks derived from plasma metabolomes provide a dynamic perspective on molecular aging, offering an enhancement to current biological age models^3,2,17^. These biomarkers have the potential to accelerate clinical applications, extend health span and lifespan, and guide strategies to promote healthy aging. First, we believe MetBAGs could be potentially used in clinical settings for risk stratification of age-related diseases, personalized intervention strategies, and optimizing clinical trial designs. Second, we emphasize the importance of investigating the dynamic shifts in the metabolome throughout the lifespan and integrating these insights into longitudinal models, particularly through multi-omics data, to better capture both programmed and stochastic aging influences. Furthermore, while we acknowledge the limitations of relying on UKBB data, we agree that validating MetBAGs in diverse populations is essential to assess their broader applicability, though platform consistency challenges may arise. Finally, further methodological advancements are needed to enhance the organ specificity of plasma metabolomics data, as these datasets, including plasma proteomics, inherently exhibit pleiotropic effects on traits associated with multiple organ systems.

## Methods

### Method 1: The MULTI consortium

The MULTI consortium is an ongoing initiative to integrate and consolidate multi-organ, such as brain MRI, and multi-omics data, including imaging, genetics, proteomics, and metabolomics. Building on existing consortia and studies, MULTI aims to curate and harmonize the data to model human aging and disease across the lifespan. This study used individual-level NMR metabolomics data from UKBB to derive the 5 MetBAGs. Summary-level GWAS summary data from FinnGen and PGC were used for downstream genetic analysis. **Supplementary eTable1** details the sample characteristics.

### UK Biobank

UKBB^44^ is a population-based research initiative comprising around 500,000 individuals from the United Kingdom between 2006 and 2010. Ethical approval for the UKBB study has been secured, and information about the ethics committee can be found here: https://www.ukbiobank.ac.uk/learn-more-about-uk-biobank/governance/ethics-advisory-committee. This study used plasma NMR metabolic biomarkers from Nightingale Health for approximately 280,000 participants in the UK Biobank. Data were collected in Phase 1 (June 2019–April 2020) and Phase 2 (April 2020–June 2022), with measurements performed on EDTA plasma samples using Nightingale’s high-throughput metabolic profiling platform. The biomarkers encompass a wide range of metabolic pathways, including lipoprotein lipids across 14 subclasses, fatty acids, and their compositions, as well as low-molecular-weight metabolites such as amino acids, ketone bodies, and glycolysis-related metabolites, all quantified in molar concentration units. In addition, imputed genotype data covering the populations of the 5 MetBAGs were used for all genetic analyses (see **Method 3** for details of our quality check for the genetic data).

### FinnGen

The FinnGen^20^ study is a large-scale genomics initiative that has analyzed over 500,000 Finnish biobank samples and correlated genetic variation with health data to understand disease mechanisms and predispositions. The project is a collaboration between research organizations and biobanks within Finland and international industry partners. For the benefit of research, FinnGen generously made their GWAS findings accessible to the wider scientific community (https://www.finngen.fi/en/access_results). This research utilized the publicly released GWAS summary statistics (version R9), which became available on May 11, 2022, after harmonization by the consortium. No individual data were used in the current study.

FinnGen published the R9 version of GWAS summary statistics via REGENIE software (v2.2.4)^45^, covering 2272 DEs, including 2269 binary traits and 3 quantitative traits. The GWAS model encompassed covariates like age, sex, the initial 10 genetic principal components, and the genotyping batch. Genotype imputation was referenced on the population-specific SISu v4.0 panel. We included GWAS summary statistics for 521 FinnGen DEs in our analyses.

### Psychiatric Genomics Consortium

PGC^21^ is an international collaboration of researchers studying the genetic basis of psychiatric disorders. PGC aims to identify and understand the genetic factors contributing to various psychiatric disorders such as schizophrenia, bipolar disorder, major depressive disorder, and others. The GWAS summary statistics were acquired from the PGC website (https://pgc.unc.edu/for-researchers/download-results/), underwent quality checks, and were harmonized to ensure seamless integration into our analysis. No individual data were used from PGC. Each study detailed its specific GWAS models and methodologies, and the consortium consolidated the release of GWAS summary statistics derived from individual studies. In the current study, we included summary data for 4 brain diseases for which allele frequencies were present.

### Method 2: Metabolomics analyses to derive the 5 MetBAGs

#### Additional quality checks

We downloaded the original data (Category ID: 220), which were analyzed and made available to the community by Nightingale Health Plc. The original data were i) calibrated absolute concentrations (or ratios) and not raw NMR spectra, and ii) before release, had already been subject to quality control (QC) procedures by Nightingale Health Plc^46^. Following the additional procedures described in Ritchie et al.^47^, we performed additional quality check steps to remove a range of unwanted technical variations, including shipping batch, 96-well plate, well position, aliquoting robot, and aliquot tip. We focused our analysis on the first instance of the metabolomics data ("instance"=0). This resulted in 327 metabolites (we used only the 107 non-derived metabolites detailed in **Supplementary eTable 2** for our main analyses) in 274,247 participants.

#### (b) Organ-specific profiles of the 107 non-derived metabolites

Plasma NMR metabolites circulate throughout the human body and predominantly influence cardiometabolic diseases. To identify the organ-specific profile of each metabolite, we annotated 107 non-derived metabolites with 2,448 Olink plasma proteins and RNA expression data from the Human Protein Atlas (HPA) project (https://www.proteinatlas.org). The 107 non-derived metabolites were selected because composite metabolites, derived by combining two or more from the 107 non-derived biomarkers, often led to model overfitting in our age prediction models, which included all 327 metabolites (**Supplementary eFigure 1**). The annotation process involved two steps. First, a linear regression model was used to link the 107 metabolites to plasma proteins, assigning the most strong associated protein to the metabolite of interest. Next, the protein’s RNA expression profile was queried in the HPA. The consensus data – combining HPA RNA-seq data and RNA-seq data from the GTEx project – was used to identify the organ or tissue with the highest normalized transcript per million (nTPM) values as the target organ (e.g., S_HDL_P→LCAT→liver: https://www.proteinatlas.org/ENSG00000213398-LCAT/tissue). This workflow resulted in a comprehensive annotation linking plasma metabolites to proteins and their associated organs.

Certain considerations warrant careful attention. Plasma metabolites, by nature, are less organ-specific compared to phenotypic data like brain MRI. As a result, a single metabolite may strongly associate with multiple proteins highly expressed across different tissues. We opted to annotate each metabolite to a single protein and organ, aiming to balance the number of features in each age prediction model and enhance organ specificity across various systems. In our annotation, the 107 metabolites were mapped to 25 unique proteins and 6 organ systems (endocrine, hepatic, immune, digestive, CNS, and heart). An interactive network visualization demonstrating this mapping is accessible at https://labs-laboratory.com/medicine/metabolite_organ_annotation. Notably, metabolites specific to the CNS and heart were excluded due to only one organ-specific metabolite for each; in addition, all 107 metabolites were retained for calculating the metabolic MetBAG. This resulted in organ-specific metabolites for a total of 5 organ systems, as shown in Fig. 1a. Furthermore, the circulative nature of plasma metabolites also echos strong correlations between the 5 MetBAGs and the underlying metabolites (Fig. 3a**-b**).

#### (d) Two AI/ML models

Our previous study systematically evaluated age prediction performance across various AI/ML models using multi-modal brain MRI features^2^ and proteomics data. Applying the same framework, we assessed the performance of models in deriving the 5 MetBAGs using one linear approach (Lasso regression and SVR) and one non-linear method (neural networks). For linear models, hyperparameter tuning was performed through nested, repeated hold-out cross-validation^48^ with 50 repetitions (80% training/validation and 20% testing). However, nested cross-validation was not applied to the neural network due to the impracticality of exhaustively testing all hyperparameter combinations. Instead, a 5-fold cross-validation was employed. An independent test dataset was held out for both models. Support vector regressor was not included here because the model does not scale with a large sample size (N>200,000).

#### (e) Population splits

To rigorously train the AI/ML models, we have split the CN (without any pathologies based on ICD code and inpatient history) data (*N*=34,354, varying across the 5 organs) into the following datasets (**Supplementary eTable 1**):

- **CN independent test dataset**: 5000 participants were randomly drawn from the CN population;
- **CN training/validation dataset**: 80% of the remaining 29,354 CN were used for the inner loop 10-fold CV for hyperparameter selection;
- **CN cross-validation test dataset**: 20% of the remaining 29,354 CN were used for the outer loop 50 repetitions;
- **PT dataset**: 239,893 patients that have at least one ICD-10-based diagnosis. Model evaluation metrics included mean absolute error (MAE) and Pearson’s *r*.

Importantly, consistent with our prior studies, only healthy control (CN) participants were included in the training/validation dataset. At the same time, individuals with any disease diagnosis were reserved for the independent PT dataset.

The BAG was determined by subtracting the participant’s chronological age from the AI/ML-predicted age, and age bias correction was applied using the approach outlined by Beheshti et al.^23^.

### Method 3: Genetic analyses

We used the imputed genotype data from UKBB for all genetic analyses. Our quality check pipeline focused on European ancestry in UKBB (6,477,810 SNPs passing quality checks), and the quality-checked genetic data were merged with respective organ-specific populations for GWAS. We summarize our genetic quality check steps. First, we skipped the step for family relationship inference^49^ because the linear mixed model via fastGWA^50^ inherently addresses population stratification, encompassing additional cryptic population stratification factors. We then removed duplicated variants from all 22 autosomal chromosomes. Individuals whose genetically identified sex did not match their self-acknowledged sex were removed. Other excluding criteria were: *i*) individuals with more than 3% of missing genotypes; *ii*) variants with minor allele frequency (MAF; dosage mode) of less than 1%; *iii*) variants with larger than 3% missing genotyping rate; *iv*) variants that failed the Hardy-Weinberg test at 1×10^-10^. To further adjust for population stratification,^51^ we derived the first 40 genetic principle components using the FlashPCA software^52^. Details of the genetic quality check protocol are described elsewhere^53,2,3,54,55^.

#### (a) GWAS

We applied a linear mixed model regression to the European ancestry populations using fastGWA^50^ implemented in GCTA^56^. We used fastGWA to perform the 5 MetBAG GWASs, adjusting age, dataset status (training/validation/test or independent test), age-squared, sex, interactions of age with sex, systolic/diastolic blood pressure, BMI, waist circumstance, standing height, weight, and the first 40 genetic principal components. We applied a genome-wide significance threshold (5□×□10□□/5) to annotate the significant independent genomic loci. Additionally, we conducted GWAS for the 327 NMR metabolites and used the resulting summary statistics for our genetic correlation analyses.

#### Annotation of genomic loci

For all GWASs, genomic loci were annotated using FUMA^57^. For genomic loci annotation, FUMA initially identified lead SNPs (correlation *r^2^*≤ 0.1, distance < 250 kilobases) and assigned them to non-overlapping genomic loci. The lead SNP with the lowest P-value (i.e., the top lead SNP) represented the genomic locus. Further details on the definitions of top lead SNP, lead SNP, independent significant SNP, and candidate SNP can be found in **Supplementary eNote 3**. For visualization purposes in Fig. 3, we have mapped the top lead SNP of each locus to the cytogenetic regions based on the GRCh37 cytoband.

#### (b) Three key genetic parameters

We used SBayesS^29^ to estimate three key genetic parameters that characterize the genetic architecture of the 5 MetBAGs. SBayesS is an expanded approach capable of estimating three essential parameters characterizing the genetic architecture of complex traits through a Bayesian mixed linear model^58^. This method only requires GWAS summary statistics of the SNPs and LD information from a reference sample. These parameters include SNP-based heritability (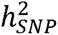), polygenicity (*Pi*), and the relationship between minor allele frequency (MAF) and effect size (*S*) as nature selection signatures. We used the software pre-computed sparse LD correlation matrix derived from the European ancestry by Zeng et al.^29^. More mathematical details can be found in the original paper from Zeng et al.^29^. We ran the *gctb* command^58^ using the argument *--sbayes S*, and left all other arguments by default.

#### (c) Genetic correlation

We estimated the genetic correlation (*g_c_*) using the LDSC^18^ software. We employed precomputed LD scores from the 1000 Genomes of European ancestry, maintaining default settings for other parameters in LDSC. It’s worth noting that LDSC corrects for sample overlap, ensuring an unbiased genetic correlation estimate^59^. We also computed the pairwise Pearson’s *r* correlation coefficient to understand whether the genetic correlation largely mirrors the phenotypic correlation (*p_c_*). Statistical significance was determined using Bonferroni correction (0.05/*N*). Three sets of analyses were performed: *i*) pair-wise *g_c_* and *p_c_* among the 5 MetBAGs (Fig. 3a), *ii*) the genetic correlation between the 5 MetBAGs and 107 non-derived raw metabolites (Fig. 3b), and *iii*) the genetic correlation between the 5 MetBAGs, and 9 PhenoBAGs^3^, 11 ProtBAGs, and 525 DEs (Fig. 3d).

#### (d) Two-sample bidirectional Mendelian randomization

We constructed a causal network linking the 5 MetBAGs, and 525 DEs using a bi-directional Mendelian randomization approach. In total, 2 bi-directional causal networks were established: i) *MetBAG2DE* and ii) *DE2MetBAG*. These networks used summary statistics from our MetBAG GWAS in the UKBB, the FinnGen^20^, and the PGC^21^ study for the 525 DEs. The systematic quality-checking procedures to ensure unbiased exposure/outcome variable and instrumental variable (IVs) selection are detailed below.

We used a two-sample Mendelian randomization approach implemented in the *TwoSampleMR* package^60^ to infer the causal relationships within these networks. We employed five distinct Mendelian randomization methods, presenting the results of the inverse variance weighted (IVW) method in the main text and the outcomes of the other four methods (Egger, weighted median, simple mode, and weighted mode estimators) in the supplement. The STROBE-MR Statement^61^ guided our analyses to increase transparency and reproducibility, encompassing the selection of exposure and outcome variables, reporting statistics, and implementing sensitivity checks to identify potential violations of underlying assumptions. First, we performed an unbiased quality check on the GWAS summary statistics. Notably, the absence of population overlapping bias^62^ was confirmed, given that FinnGen and UKBB participants largely represent populations of European ancestry without explicit overlap with UKBB. PGC GWAS summary data were ensured to exclude UKBB participants. Furthermore, all consortia’s GWAS summary statistics were based on or lifted to GRCh37. Subsequently, we selected the effective exposure variables by assessing the statistical power of the exposure GWAS summary statistics in terms of instrumental variables (IVs), ensuring that the number of IVs exceeded 7 before harmonizing the data. Crucially, the function "*clump_data*" was applied to the exposure GWAS data, considering LD. The function "*harmonise_data*" was then used to harmonize the GWAS summary statistics of the exposure and outcome variables. Bonferroni correction was applied to all tested traits based on the number of effective DEs.

Finally, we conducted multiple sensitivity analyses. First, we conducted a heterogeneity test to scrutinize potential violations in the IV’s assumptions. To assess horizontal pleiotropy, which indicates the IV’s exclusivity assumption^63^, we utilized a funnel plot, single-SNP Mendelian randomization methods, and the Egger estimator. Furthermore, we performed a leave-one-out analysis, systematically excluding one instrument (SNP/IV) at a time, to gauge the sensitivity of the results to individual SNPs.

#### (e) PRS calculation

PRS was computed using split-sample GWASs (split1 and split2) for the 5 MetBAG GWASs. The PRS weights were established using split1/discovery GWAS data as the base/training set, while the split2/replication GWAS summary statistics served as the target/testing data. Both base and target data underwent rigorous quality control procedures involving several steps: *i*) excluding duplicated and ambiguous SNPs in the base data; *ii*) excluding high heterozygosity samples in the target data; and *v*) eliminating duplicated, mismatching, and ambiguous SNPs in the target data.

After completing the QC procedures, PRS for the split2 and split1 groups was calculated using the PRS-CS^64^ method. PRC-CS applies a continuous shrinkage prior, which adjusts the SNP effect sizes based on their LD structure. SNPs with weaker evidence are "shrunk" toward zero, while those with stronger evidence retain larger effect sizes. This avoids overfitting and improves prediction performance. The shrinkage parameter was not set, and the algorithm learned it via a fully Bayesian approach. The predictive power of the PRS for the MetBAG was evaluated using its incremental *R^2^* (**Supplementary eTable 11**).

### Method 4: Prediction analyses for 14 systemic disease categories, the risk of mortality, and single disease endopoints

We investigated the clinical promise of the 5 MetBAGs and their PRSs (MetBAG-PRS) in two sets of prediction analyses: *i*) classification tasks for predicting 14 systemic disease categories based on the ICD-10 code, *ii*) survival analysis for the risk of all-cause mortality, and *iii*) ICD-based single disease endpoints.

#### (a) Neural networks to classify patients of disease categories vs. controls

We applied a neural network with MetBAG and their PRS. 5-fold cross-validation was used for the training/test dataset (*N* varies across disease categories and the MetBAG), and the model was then applied to an independent test dataset (*N*=10,000) with the same distribution of PT/CN ratios. Patients for each disease category were defined by the ICD-10 code (Field-ID: 41270). Figure 5a reports the balanced accuracy (BA) obtained from the independent test data. **Supplementary eTable 8** provides detailed metrics, including balanced accuracy, sensitivity, specificity, negative predictive value, positive predictive value, and sample sizes for the training/test datasets.

#### (b) Survival analysis for mortality risk

we employed a Cox proportional hazard model while adjusting for covariates(i.e., age and sex) to test the associations of the 5 MetBAGs and 5 MetBAG-PRSs with all-cause mortality. The covariates were included as additional right-side variables in the model. The hazard ratio (HR), exp(β*_R_*), was calculated and reported as the effect size measure that indicates the influence of each biomarker on the risk of mortality. To train the model, the "time" variable was determined by calculating the difference between the date of death (Field ID: 40000) for cases (or the censoring date for non-cases) and the date attending the assessment center (Field ID: 53). Participants who passed away after enrolling in the study were classified as cases.

#### (c) Survival analysis for ICD-based single disease endpoint

we employed a Cox proportional hazard model while adjusting for covariates(i.e., age and sex) to test the associations of the 5 MetBAGs with the incidence of ICD-based single disease entity. The covariates were included as additional right-side variables in the model. To train the model, the "time" variable was determined by calculating the difference between the date of death (Field ID: 40000) for cases (or the censoring date for non-cases) and the date attending the assessment center (Field ID: 53). Participants who were diagnosed for a specific disease of interest after enrolling in the study were classified as cases.

## Data Availability

The GWAS summary statistics and pre-trained AI models from this study are publicly accessible via the MEDICINE Knowledge Portal (https://labs-laboratory.com/medicine/) and Synapse (https://www.synapse.org/Synapse:syn64923248/wiki/630992^65^). Our study used data generated by the human protein atlas (HPA: https://www.proteinatlas.org). GWAS summary data for the DEs were downloaded from the official websites of FinnGen (R9: https://www.finngen.fi/en/access_results) and PGC (https://pgc.unc.edu/for-researchers/download-results/). Individual data from UKBB can be requested with proper registration at https://www.ukbiobank.ac.uk/. Certain sensitive data (e.g., allele frequency information) supporting the findings are also available from the author upon request.

## Code Availability

The software and resources used in this study are all publicly available:

- MLNI^66^: https://github.com/anbai106/mlni, MetBAG generation and classification for disease categories;
- ukbnmf: https://github.com/sritchie73/ukbnmr, Quality checks for the raw metabolites;
- FUMA: https://fuma.ctglab.nl/, Gene mapping, genomic locus annotation;
- GCTA: https://yanglab.westlake.edu.cn/software/gcta/#Overview, fastGWA;
- LDSC: https://github.com/bulik/ldsc, genetic correlation
- TwoSampleMR: https://mrcieu.github.io/TwoSampleMR/index.html, Mendelian randomization;
- PRScs: https://github.com/getian107/PRScs, PRS calculation;
- Lifelines: https://lifelines.readthedocs.io/en/latest/, Survival analysis;
- GCTB: https://cnsgenomics.com/software/gctb/; SBayesS.

## Authors’ contributions

Dr. Wen has full access to all the study data and is responsible for its integrity and accuracy.

*Study concept and design*: W.J.

*Drafting of the manuscript*: W.J.

*Critical revision of the manuscript for important intellectual content*: All authors

*Statistical analysis*: W.J.

## Acknowledgments

The MULTI consortium (J.W; UK Biobank Application Number: 647044) aims to integrate multi-organ imaging and multi-omics data to advance our understanding of human aging and disease mechanisms. This study used the UK Biobank resource under Application Number 35148. We want to express our sincere gratitude to the UK Biobank team for their invaluable contribution to advancing clinical research in our field (https://www.ukbiobank.ac.uk/). We want to acknowledge the participants and investigators of the FinnGen study and the PGC consortium, and we thank FinnGen (https://www.finngen.fi/en) and PGC (https://pgc.unc.edu/) for their generosity in sharing the GWAS summary statistics with the scientific community.

## Extended Data Figures

**Extended Data Figure 1:**
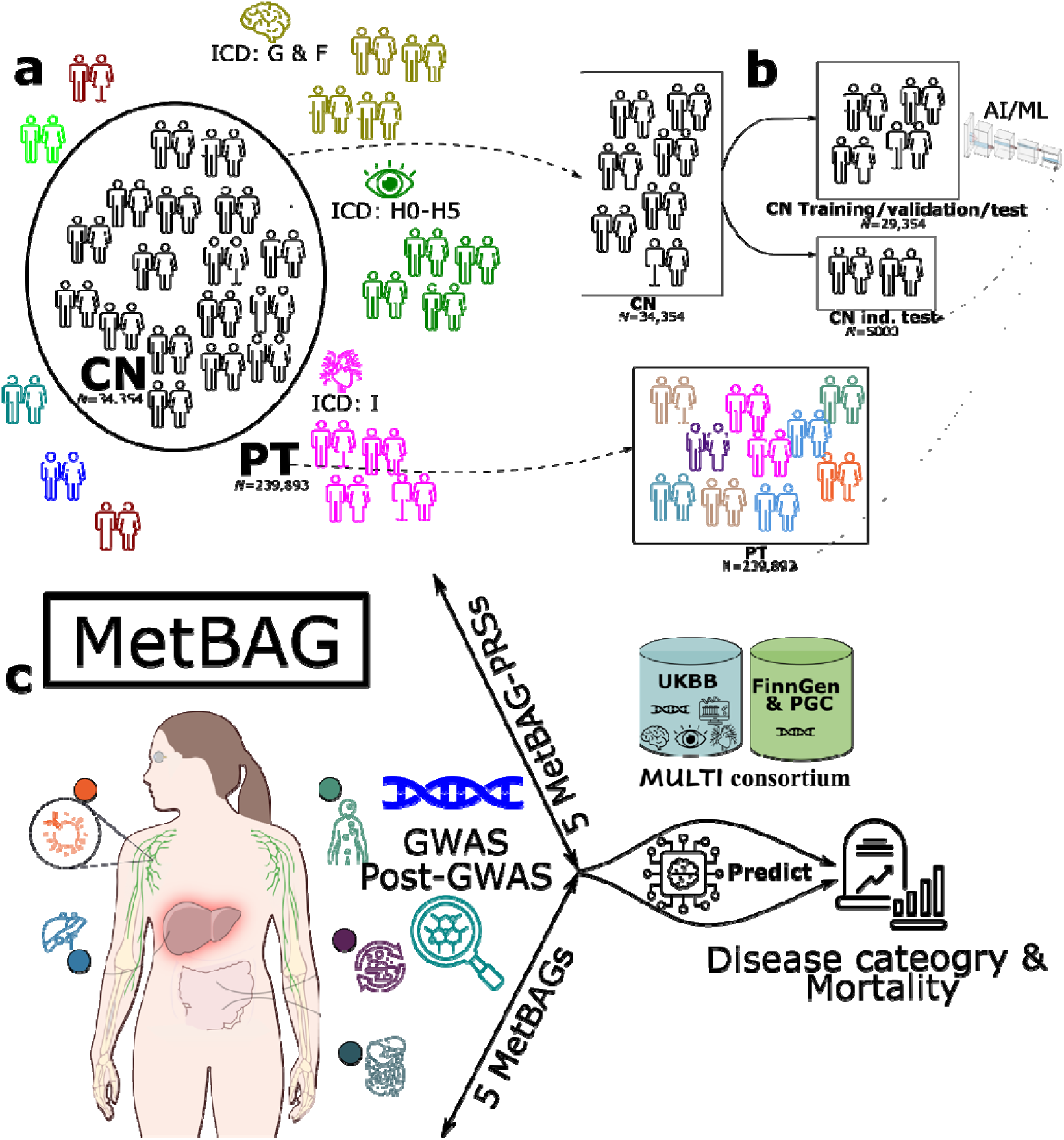
Schematic diagram of the definition of populations to derive MetBAG and overall analytic workflow of the study. **a)** We split the UK Biobank metabolomics population into 34,354 healthy controls (CN) and 239,893 patients (PT) based on ICD-10 codes and clinical history. **b)** To develop the 5 MetBAGs, we trained two AI/ML models exclusively on the CN training/validation/test population (N=29,354) using a (nested) cross-validation procedure to identify the optimal model. Independent test datasets included the CN independent test set (N=5000) and the PT population (N=239,893). c) The study’s analytical workflow encompassed deriving the 5 MetBAGs and their PRSs, performing GWAS and post-GWAS analyses, and assessing the predictive performance of the MetBAGs and MetBAG-PRSs across 14 systemic disease categories and mortality outcomes.

**Extended Data Figure 2:**
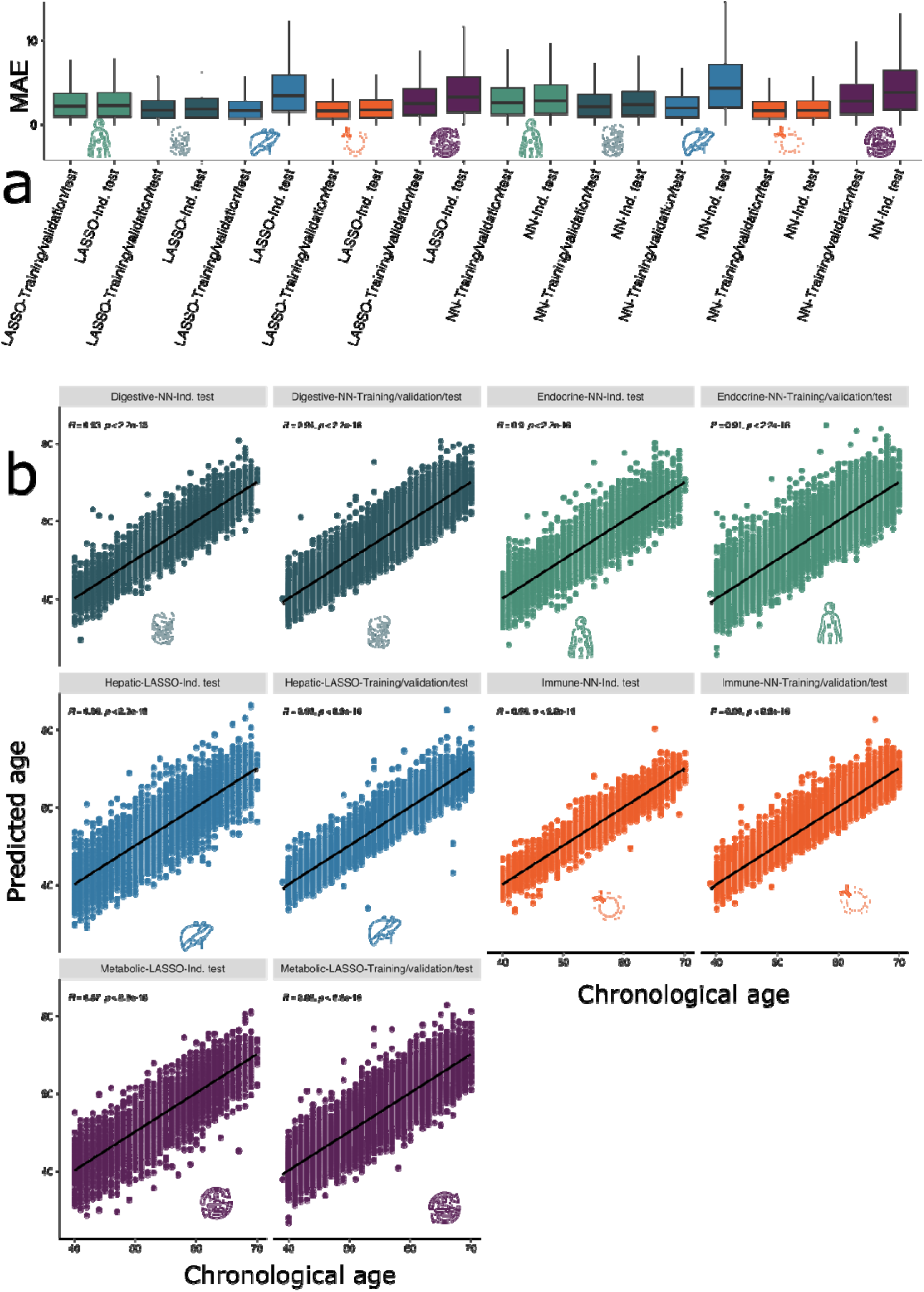
The model generalizability and scatter plot between AI/ML-predicted biological age and chronological age after age bias correction. **a)** MAE for each AI/ML model in the CN training/validation/test dataset and independent test dataset after age bias correction. **b)** The scatter plot between the AI/ML-derived biological age and chronological age after the age bias correction is applied.

**Extended Data Figure 3:**
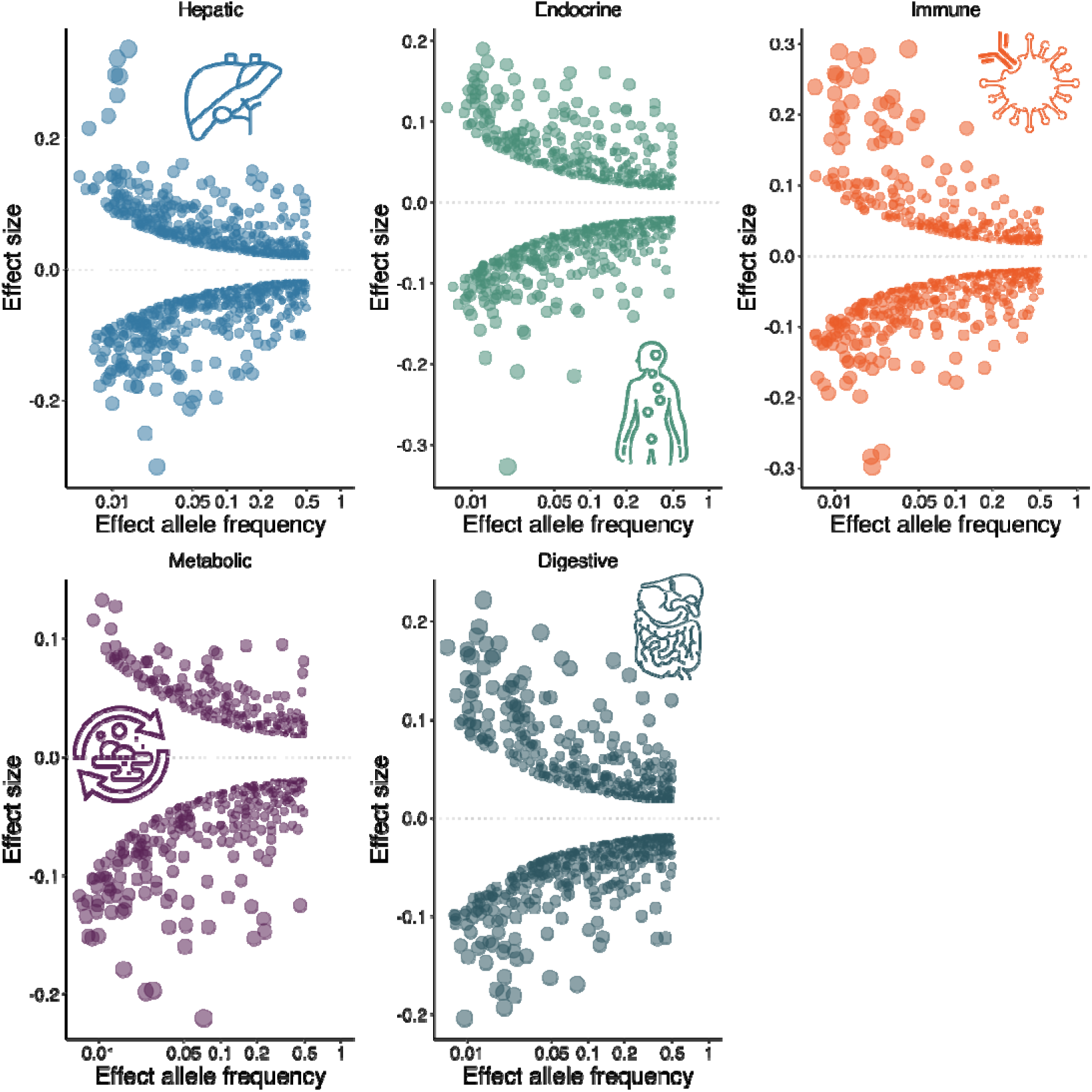
Trumpet plots of the effect allele frequency vs. the *β* coefficient of the 5 MetBAG GWASs. The trumpet plots display the inverse relationship between the alternative (effect) allele frequency and the effect size (*β* coefficient) for the 5 MetBAGs. We present the independent significant SNPs defined in FUMA. The dot size corresponds to the effect size, while the transparency of the dot is proportional to its statistical significance.

**Extended Data Figure 4:**
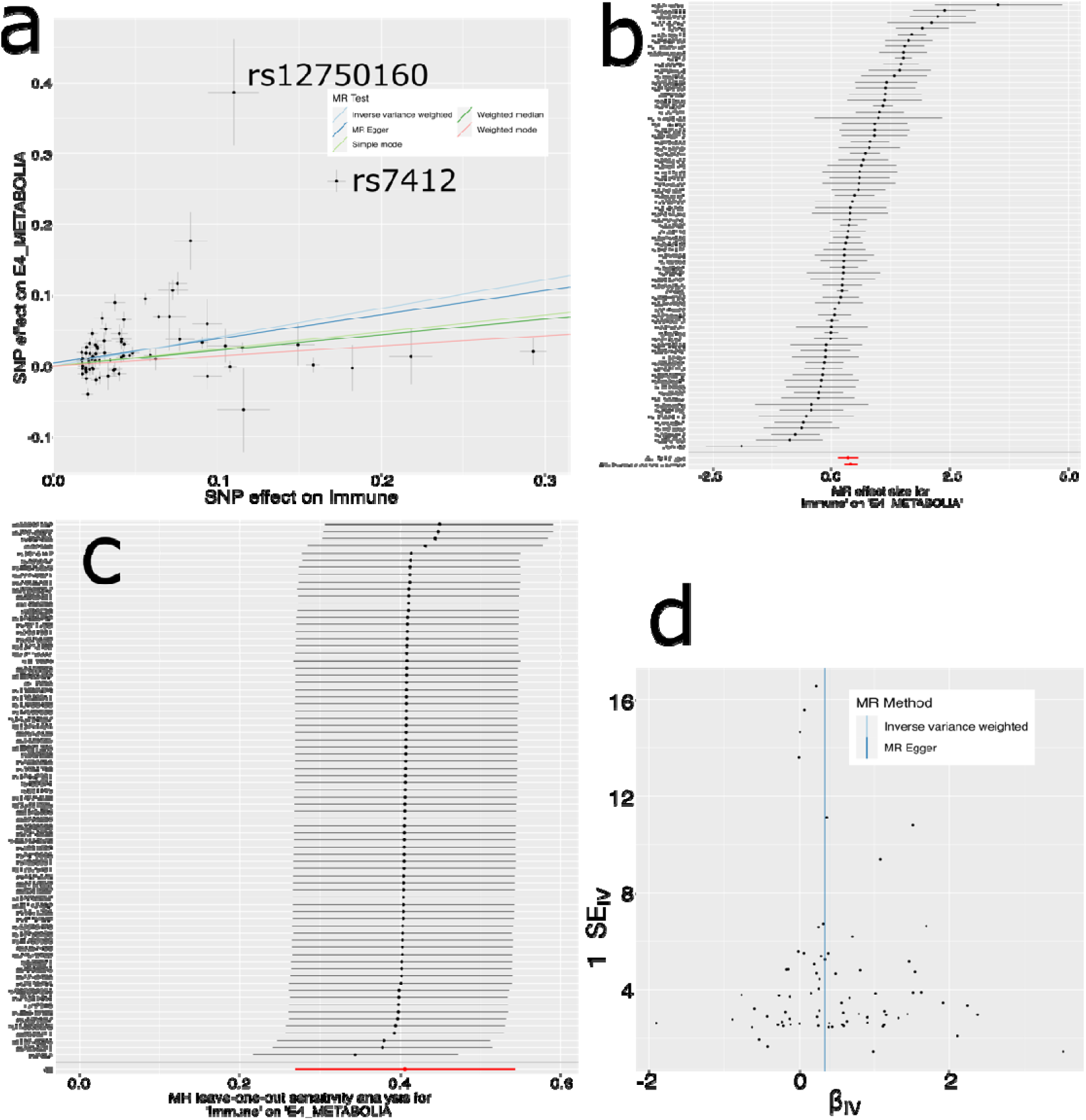
Sensitivity check analyses for the causal pathway of “Immune MetBAG→E4_METABOLIA”. **a**) Scatter plot for the MR effect sizes of the SNP-Immune MetBAG association (*x*-axis, log OR) and the SNP-E4_METABOLIA associations (*y*-axis, log OR) with standard error bars. The slopes of the five lines correspond to the causal effect sizes estimated by the five MR estimators, respectively. Two outlier IVs are annotated with their SNP rs number; we re-performed the MR by excluding the two outliers as IVs. **b**) Forest plot for the single-SNP MR results. Each dot represents the MR effect (log OR)), and the error bar displays the 95% CI for the immune MetBAG on E4_METABOLIA using only one SNP; the red line shows the MR effect using all SNPs together for IVW and MR Egger estimators. **c**) Leave-one-SNP-out analysis of the immune MetBAG on E4_METABOLIA. Each dot represents the MR effect (log OR), and the error bar displays the 95% CI by excluding that SNP from the analysis. The red line depicts the IVW estimator using all SNPs. **d**) Funnel plot for the relationship between the causal effect of the immune MetBAG on E4_METABOLIA. Each dot represents MR effect sizes estimated using each SNP as a separate instrument against the inverse of the standard error of the causal estimate.

**Extended Data Figure 5:**
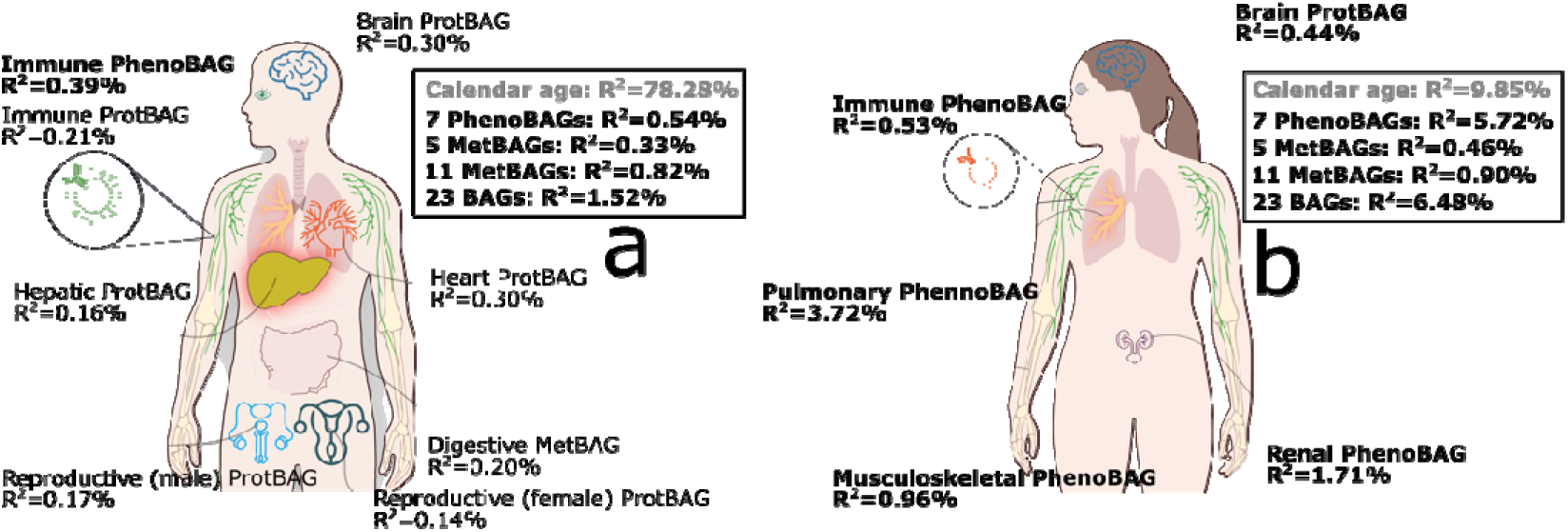
23 multi-organ, multi-omics BAGs provide incremental *R^2^* on top of calendar/chronological age for prediction of all-cause mortality and respiratory disease categories. Using our previously developed 7 PhenoBAGs^3^ (excluding brain and eye due to sample size constraints after merging BAG populations), the 5 MetBAGs from the current study, and 11 ProtBAGs^67^, we evaluated their additional predictive power – quantified as incremental *R^2^* – beyond calendar age and sex. This analysis focused on predicting **a**) all-cause mortality (*N*=658 participants) and **b**) the respiratory disease category (ICD=J) in 866 healthy controls (without any ICD diagnosis) and 1590 patients, which yielded the most significant results among 14 ICD-based systemic disease categories tested. The null model included only calendar age, sex, and genetic principal components as predictors (grey-colored R^2^ values), while the alternative model incorporated additional BAGs. Significant results (P-value < 0.05/23) are highlighted in bold. The same survival analysis model was applied as described in **Method 4b** and **4c**.

