## Supplementary materials for "Multi-organ metabolome biological age implicates cardiometabolic conditions and mortality risk"

**Online Supplementary Materials**

**eNote 1:** **Potential factors leading to overfitting and poor generalization to independent test datasets**

**eNote 2:** **Sex-stratified analyses for the metabolic MetBAG**

**eNote 3:** **The definition of genomic loci, independent significant SNP, lead SNP, candidate SNP**

**eNote 4:** **The SNP-based heritability estimates for the 5 MetBAGs, 11 ProtBAGs, and 9 PhenoBAGs using SBayesS**

**eNote 5:** **Sensitivity check analyses for an example of the MR results (Immune MetBAG→E4_METABOLIA)**

**eFigure 1: Incorporating composite metabolites led to overfitting**

**eFigure 2: Sex-stratified results for the metabolic MetBAG**

**eFigure 3: Comparisons of the SNP-based heritability across different omics-based BAGs**

**eFigure 4: The systemic disease category classification performance with age and sex as additional features**

**eFigure 5: The training and test loss of the neural network**

**eFigure 6: Using 5 MetBAGs to predict the incidence of ICD-based single disease entities in the UK Biobank**

**eTable 1: The characteristics of the MULTI consortium**

**eTable 2: The organ-specific metabolite annotations**

**eTable 3: Biological age prediction performance before and after age bias correction**

**eTable 4: Genomic loci of the 5 MetBAG GWASs**

**eTable 5: The detailed statistics of the three key genetic parameters of SBayesS**

**eTable 6: The genetic correlation results**

**eTable 7: The Mendelian randomization results**

**eTable 8: The classification results to predict the 14 systemic disease categories**

**eTable 9: The survival analysis to predict mortality**

**eTable 10: The survival analysis to predict ICD-based disease incidence**

**eTable 11: The incremental R2 of the 5 MetBAG-PRS to predict the MetBAG**

**eFolder 1: Sensitivity analyses for the MR analyses**

**eNote 1:** **Potential factors leading to overfitting and poor generalization to independent test datasets**

We identified two potential sources of overfitting in certain models of the 5 MetBAGs and conducted additional analyses to mitigate them.

**Overly complex neural networks lead to overfitting**

For instance, lasso outperformed NN for the hepatic MetBAG, showing minimal overfitting, with the MAE increasing only slightly from 6.14 in the independent test dataset to 6.58 in the training/validation/test dataset (Cohen’s D=0.108). In contrast, the NN exhibited substantial overfitting, with the MAE rising significantly from 6.18 in the independent test dataset to 11.7 in the training/test dataset (Cohen’s D=1.160).

One possible reason for this overfitting phenomenon was potential domain shift in demographics and pathologies between training/validation and independent test datasets (NN was not performed with nested CV as it is too liberal regarding the choices of possible NN architectures). However, this was ruled out since the Lasso model, which used the same (nested) cross-validation (CV) splits, did not exhibit overfitting.

We then hypothesized that the 3-layer NN was too complex for the hepatic MetBAG dataset, which contained only 22 hepatic-specific metabolites, with 28142 CN participants as the training dataset. To test this, we reduced the NN complexity by decreasing the number of nodes per layer. This reduction decreased the number of trainable parameters from 11,265 to 291. The overfitting issue disappeared when evaluating the trained model on the independent test set, where the mean absolute error (MAE) remained stable (MAE-CV-training = 6.28 years; MAE-ind.-test = 6.47 years). Additionally, training loss monitoring confirmed that the light-version NN did not exhibit significant overfitting from CV-training to CV-validation datasets (unlike the original NN).

To summarize, training a neural network (NN) is essential for identifying potential overfitting, especially since implementing nested cross-validation is challenging due to computational complexity. However, the independent test dataset provides an additional safeguard to ensure that overfitted models are not used in downstream analyses.

**High collinearity from composite metabolite results in overfitting**

Another source of overfitting was the integration of composite metabolite features (i.e., sums or ratios of the 107 original metabolites). These composite metabolites inherently introduced high collinearity into the feature set, leading to substantial overfitting. As shown in **eFigure 1**, models incorporating composite features exhibited poor generalizability, further supporting this conclusion.

By addressing these two overfitting sources – reducing model complexity and avoiding redundant composite metabolites – we improved the robustness and generalizability of the hepatic MetBAG model.

**eNote 2:** **Sex-stratified analyses for the metabolic MetBAG**

Recent studies have identified sex differences in aging clocks across multiple omics levels. For instance, Moguilner et al.^1^ examined brain aging clocks derived from EEG data in aging and dementia across geographically diverse populations. Their analysis included 5,306 participants from 15 countries, spanning both Latin American and Caribbean (LAC) and non-LAC regions. The cohort comprised healthy controls and individuals diagnosed with mild cognitive impairment (MCI), Alzheimer’s disease (AD), and behavioral variant frontotemporal dementia (bvFTD). Regarding sex differences, they found that in LAC regions, females exhibited larger brain-age gaps than males within both the control and AD groups when the model was trained on combined-sex data (**Fig. 4** in their paper).

In another study, Argentieri et al.^2^ investigated the influence of sex on proteome-based aging clocks using UK Biobank data. Their findings revealed similar age prediction performance between sex-specific models (i.e., training the data using data exclusively from one sex) and a combined-sex model (i.e., training the model using data from both sexes), with predicted ages from both approaches showing near-perfect correlation. Notably, the female-only model achieved a slightly better fit (MAE = 2.25) compared to the male-only model (MAE = 2.45). Both models incorporated over 2,000 proteins and were not organ specific.

In a previous study^3^, I explored sex differences in proteomics across 11 multi-organ ProtBAGs, providing further evidence of sex biases in proteome-derived aging clocks. Notably, sex differences were evident in the brain ProtBAG, where male-specific models exhibited greater susceptibility to overfitting compared to female-specific models. While predicted ages from sex-specific and combined-sex models were highly consistent (i.e., strongly correlated), it is essential to address challenges such as model overfitting and domain shift rather than relying solely on Pearson’s correlation coefficient for evaluation.

Here, we assessed whether such age biases existed in the proteome-based aging clocks. Using the metabolic MetBAG as an example, we found that the metabolic MetBAG was evidence between the males (MAE=6.5; BAG=2.63) and females (MAE=5.7; BAG=0.417) regarding both MAE and BAG in the ind. test dataset (**eFigure 2a**). Additionally, despite differences in data availability between the two training approaches (sex-specific vs. sex-combined data), the metabolic MetBAG in males (Pearson’s *r*=0.92) and females (Pearson’s *r*=0.84) remains highly correlated across both methods (**eFigure 2b-c**). Finally, we examined sex differences in GWAS signals and found substantial overlap in the genetics of male- and female-specific MetBAG. However, we also identified distinct signals, such as those on chromosome 2.

In summary, sex differences are evident in the metabolic MetBAG, with males exhibiting higher MAE and BAG than females. Although predicted ages from sex-specific models and those trained on combined data are highly correlated, it is essential to address challenges such as model overfitting and domain shift rather than relying solely on Pearson’s *r* for evaluation. Additionally, future studies on sex-specific GWAS are critical for further insights.

**eNote 3:** **The definition of genomic loci, independent significant SNP, lead SNP, candidate SNP**

FUMA defined the significant independent SNPs, lead SNPs, candidate SNPs, and genomic risk loci as follows ([https://fuma.ctglab.nl/tutorial#snp2gene](https://fuma.ctglab.nl/tutorial%23snp2gene)):

*Independent significant SNPs*

They are defined as SNPs with *P*≤5×10^-8^ that are independent of each other at the user-defined *r^2^* (set to 0.6 in the current study). We further describe *candidate SNPs* as those in linkage disequilibrium (LD) with independent significant SNPs. FUMA then queries each candidate SNP in the GWAS Catalog to check whether any clinical traits have been reported to be associated with previous GWAS studies.

*Lead SNPs*

Lead SNPs are defined as independent significant SNPs that are also independent of each other at *r^2^*<0.1. If multiple independent significant SNPs are correlated at *r^2^*≥0.1, then the one with the lowest individual *P*-value becomes the lead SNP. If *r^2^* threshold is set to 0.1 for the independent significant SNPs, then they would constitute the identical set as the lead SNPs. FUMA thus advises setting *r^2^* to be 0.6 or higher.

*Genomic risk loci*

FUMA defines genomic risk loci to include all independent signals physically close or overlapping in a single locus. First, independent significant SNPs dependent on each other at *r^2^*≥0.1 are assigned to the same genomic risk locus. Then, independent significant SNPs with less than the user-defined distance (250 kilobases by default) away from one another are merged into the same genomic risk locus – the distance between two LD blocks of two independent significant SNPs is the distance between the closest points from each LD block. Each locus is represented by the SNP within the locus with the lowest *P*-value.

**eNote 4:** **The SNP-based heritability estimates for the 5 MetBAGs, 11 ProtBAGs, and 9 PhenoBAGs using SBayesS**

Our study, along with others, has demonstrated that SNP-based heritability estimates can vary depending on the approach used, both at the data level (e.g., GWAS summary statistics vs. raw individual genotype data) and the methodological level (e.g., LDSC vs. GCTA-GREML). In general, methods utilizing raw individual-level data tend to yield higher heritability estimates than those relying on summary statistics, such as the commonly used LDSC method. However, our previous research found that despite differences in magnitude, heritability estimates across these methods were highly correlated^4^.

Here, we compared the SNP-based heritability estimates of the 5 MetBAGs to those of our previously developed 11 ProtBAGs and nine PhenoBAGs using the SBayesS method, ensuring an "apple-to-apple" comparison (**eFigure 3**). It is important to note that while the genetic data underwent the same quality control procedures, the populations and sample sizes differed across MetBAGs, PhenoBAGs, and ProtBAGs. Theoretically, this discrepancy should affect only the standard error (SE) of the estimates rather than the absolute magnitude of SNP-based heritability, as SBayesS derives its estimates from GWAS summary statistics. The heritability estimate primarily depends on SNP effect sizes rather than directly on sample size.

Several key observations emerged from our analysis. First, the highest SNP-based heritability estimate was observed for the brain PhenoBAG (0.27 ± 0.01), followed by the skin ProtBAG (0.20 ± 0.01) and the pulmonary PhenoBAG (0.199 ± 0.003). Second, when comparing across different omics data types, the mean heritability estimates for the 9 PhenoBAGs (0.155) was slightly higher than that of the 5 MetBAGs (0.139) and 11 ProtBAGs (0.137). PhenoBAG traits likely show higher heritability due to stronger genetic influence, better measurement precision, and improved LD tagging in GWAS. In contrast, metabolomic and proteomic traits may be more affected by complex regulation and environmental factors, leading to lower estimates. However, this interpretation requires cautious validation and further investigation.

**eNote 5:** **Sensitivity check analyses for an example of the MR results (Immune MetBAG→E4_METABOLIA)**

As Mendelian randomization is sensitive to underlying IV assumptions, we performed sensitivity analyses to investigate the potential violation, exemplified by the three-layer causal pathway of Immune MetBAG→E4_METABOLIA.

For the causal relationship from the immune MetBAG→E4_METABOLIA, we observed two potential outlier instrumental variables (IV; i.e., independent SNPs: rs12750160 and rs7412) for the effect sizes on the exposure and outcome variables (**Extended Fig. 4 a**). We excluded those two outliers to rerun the analysis; the causal effect persisted, and the *beta* value slightly decreased (from 0.40 to 0.34).

We then showed the forest plot for the causal effect sizes at the individual SNP level, indicating that most IVs exert positive effects (**Extended Fig. 4 b**). As evidenced by Cochran’s Q test (P = 0.03), the IV estimates from each genetic variant were slightly heterogeneous. We then performed a leave-one-IV-out analysis and found that no single SNPs largely dominated the causal effect (**Extended Fig. 4 c**). Finally, we showed a symmetry funnel plot that indicates no potential “directional pleiotropy” (**Extended Fig. 4 d**). To further scrutinize this bias, we applied MR-Egger regression with MAF-corrected weights to the summarized data, yielding an intercept estimate of 0.0040 with an associated P-value of 0.44. This further supports the absence of directional pleiotropy. In summary, there is no evidence that directional pleiotropy is to bias the conclusion.

**eFigure 1: Incorporating composite metabolites led to overfitting**

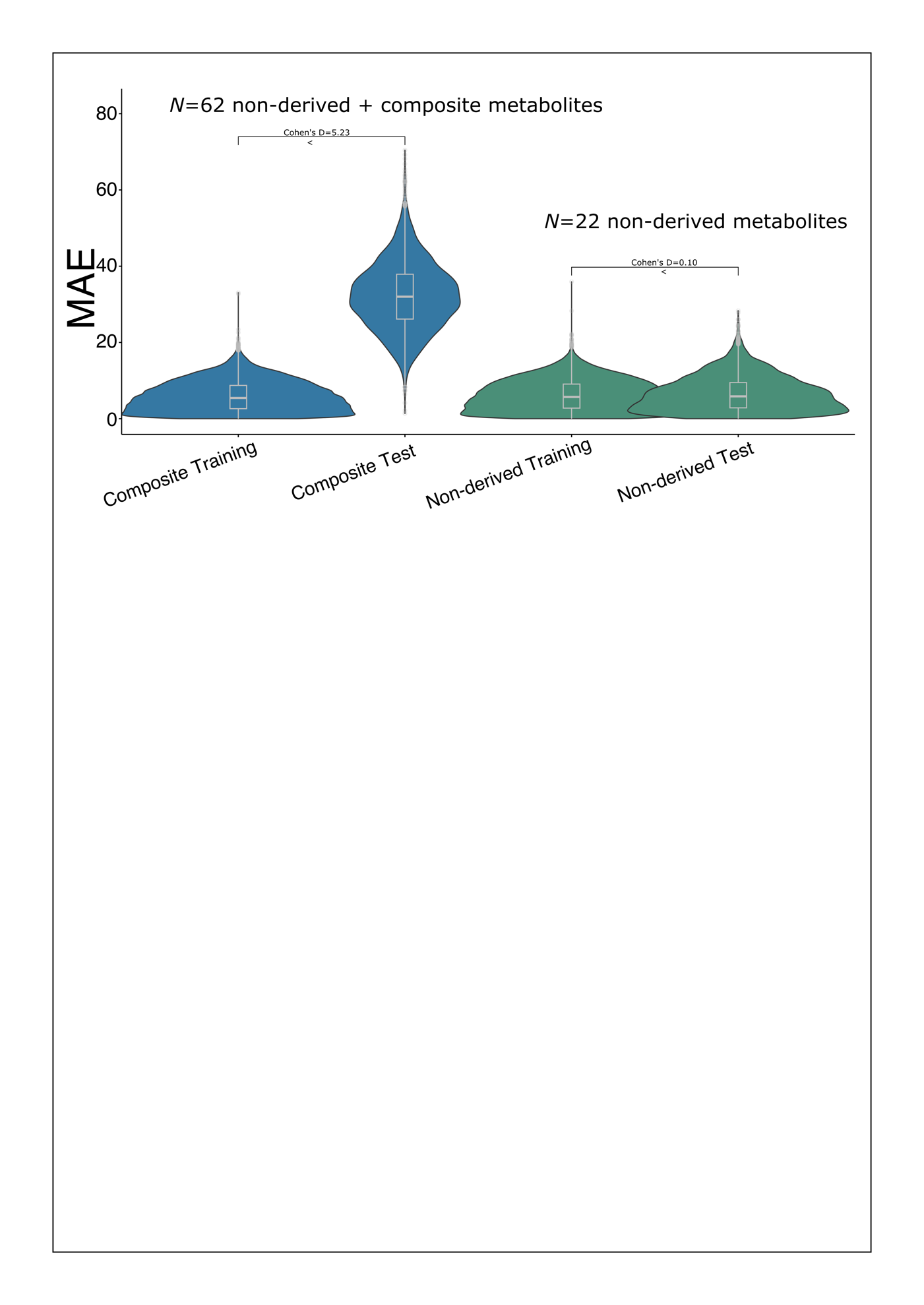

Including composite metabolites in the training features, as demonstrated with the hepatic MetBAG using Lasso regression, resulted in significant overfitting and poor generalizability to the independent dataset.

**eFigure 2: Sex-stratified results for the metabolic MetBAG**

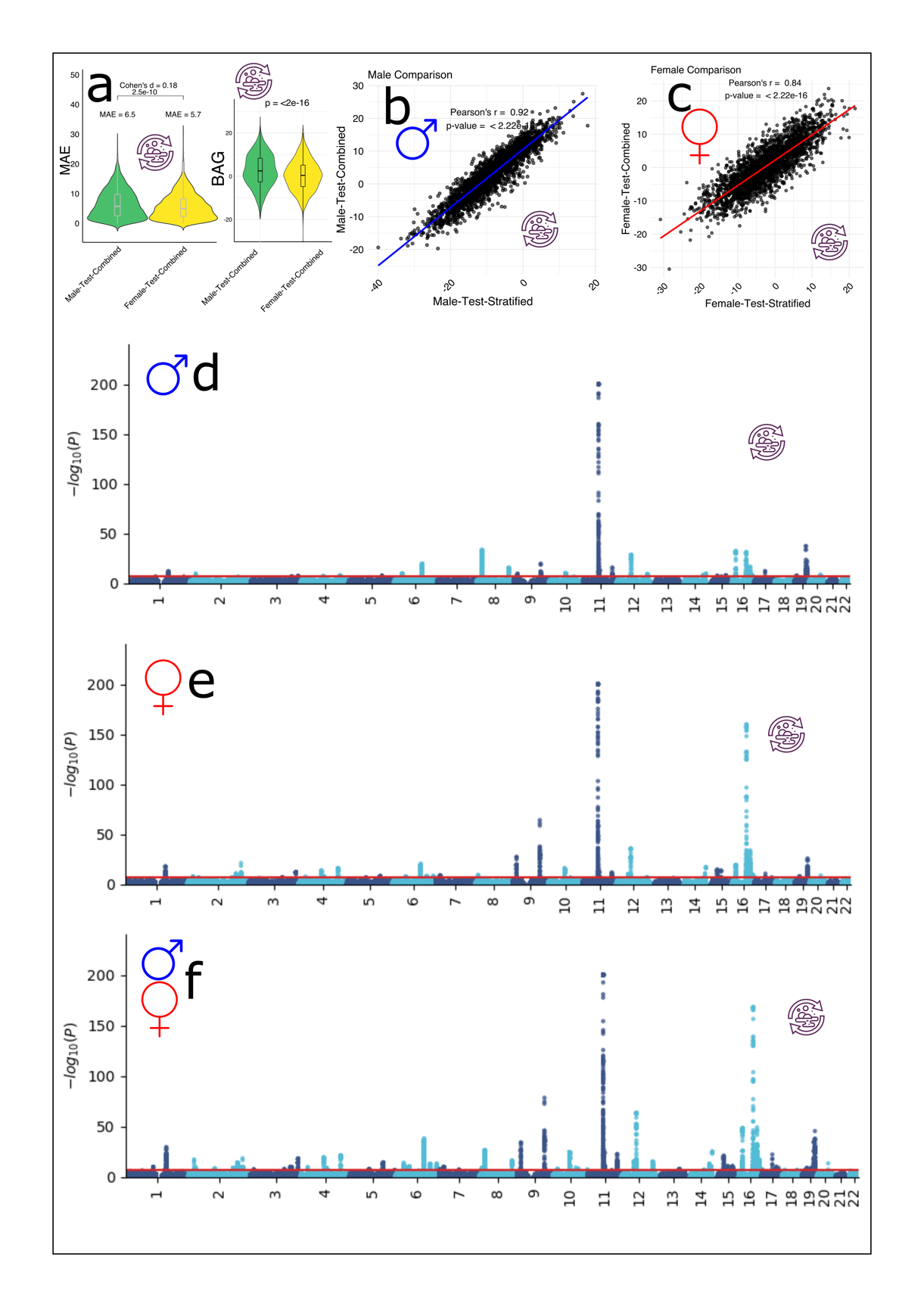

**a**) Male-only and female-only results from ind. test datasets using the Lasso model via the metabolic MetBAG. The distribution for MAE and BAG between the two sexes are presented. **b**) We conducted a sex-stratified analysis using Lasso to derive brain ProtBAG separately for males and females. This approach differs from Figure **a**), where the ML model was trained on data from both sexes combined. Following the method of Argentieri et al.³, we created a scatter plot comparing two approaches: i) a model trained exclusively on male data and ii) a model trained on data from both sexes combined. **c**) The same approach as in **b**) but for female-only data. **d-f**). The Manhattan plot for the metabolic MetBAG from three different approaches: i) male-only data, ii) female-only data, and iii) data from both sexes.

**eFigure 3: Comparisons of the SNP-based heritability across different omics-based BAGs**

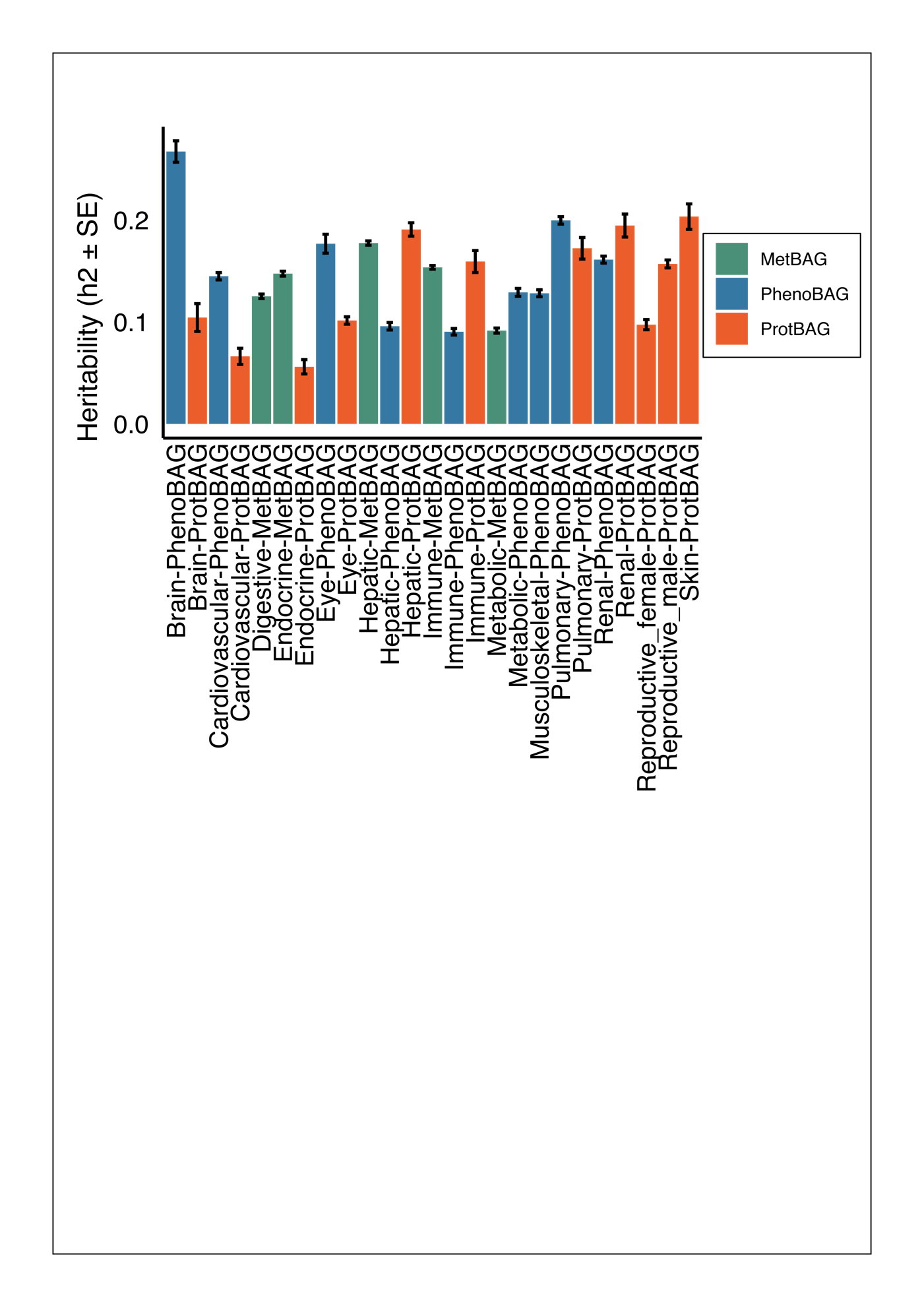

The SNP-based heritability of the 5 MetBAGs, 11 ProtBAGs, and 9 PhenoBAGs using the SBayesS method.

**eFigure 4: The systemic disease category classification performance with age and sex as additional features**

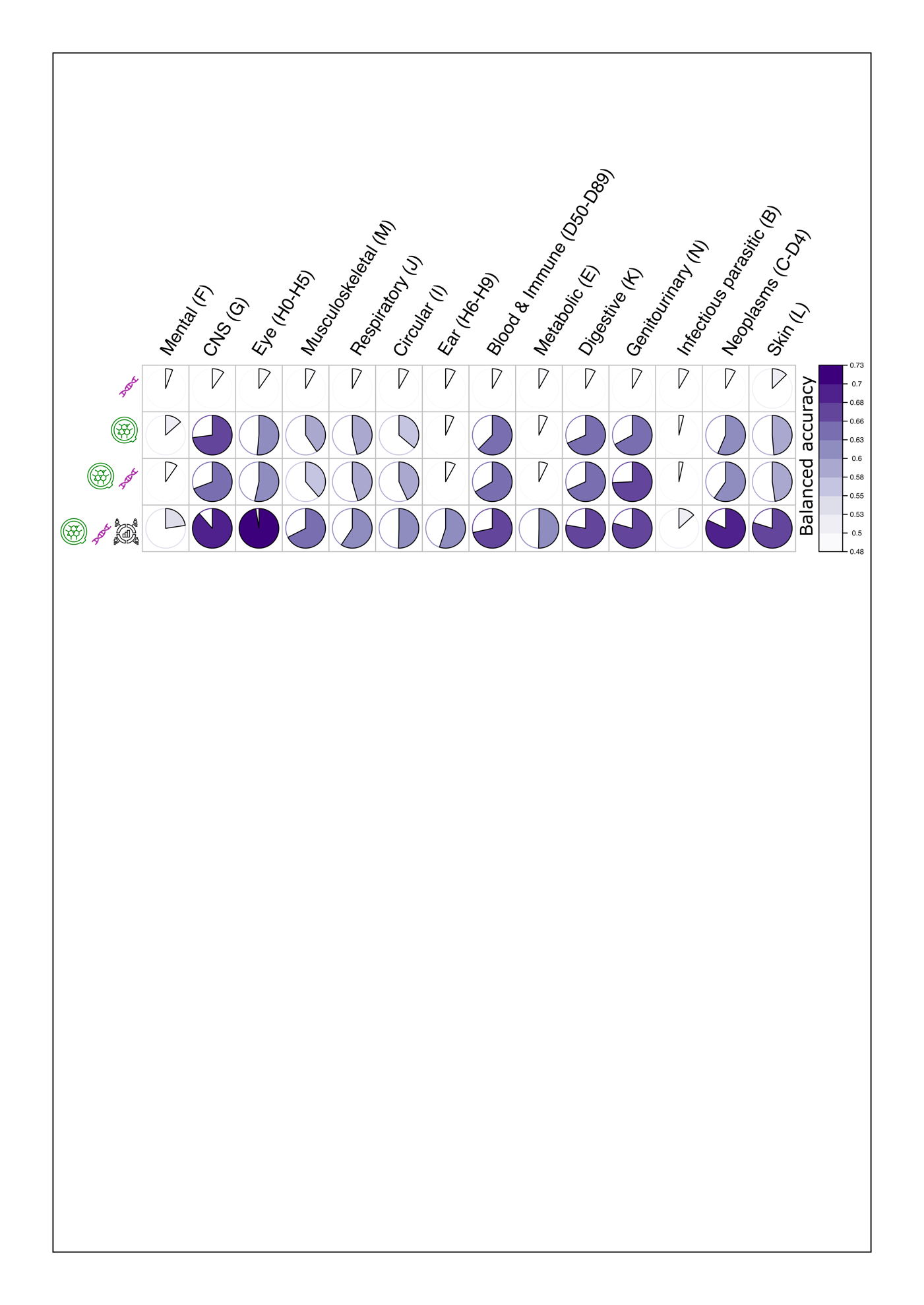

Classification results (balanced accuracy) by adding age and sex as additional features on top of the 5 MetBAGs and MetBAG-PRSs.

**eFigure 5: The training and test loss of the neural network**

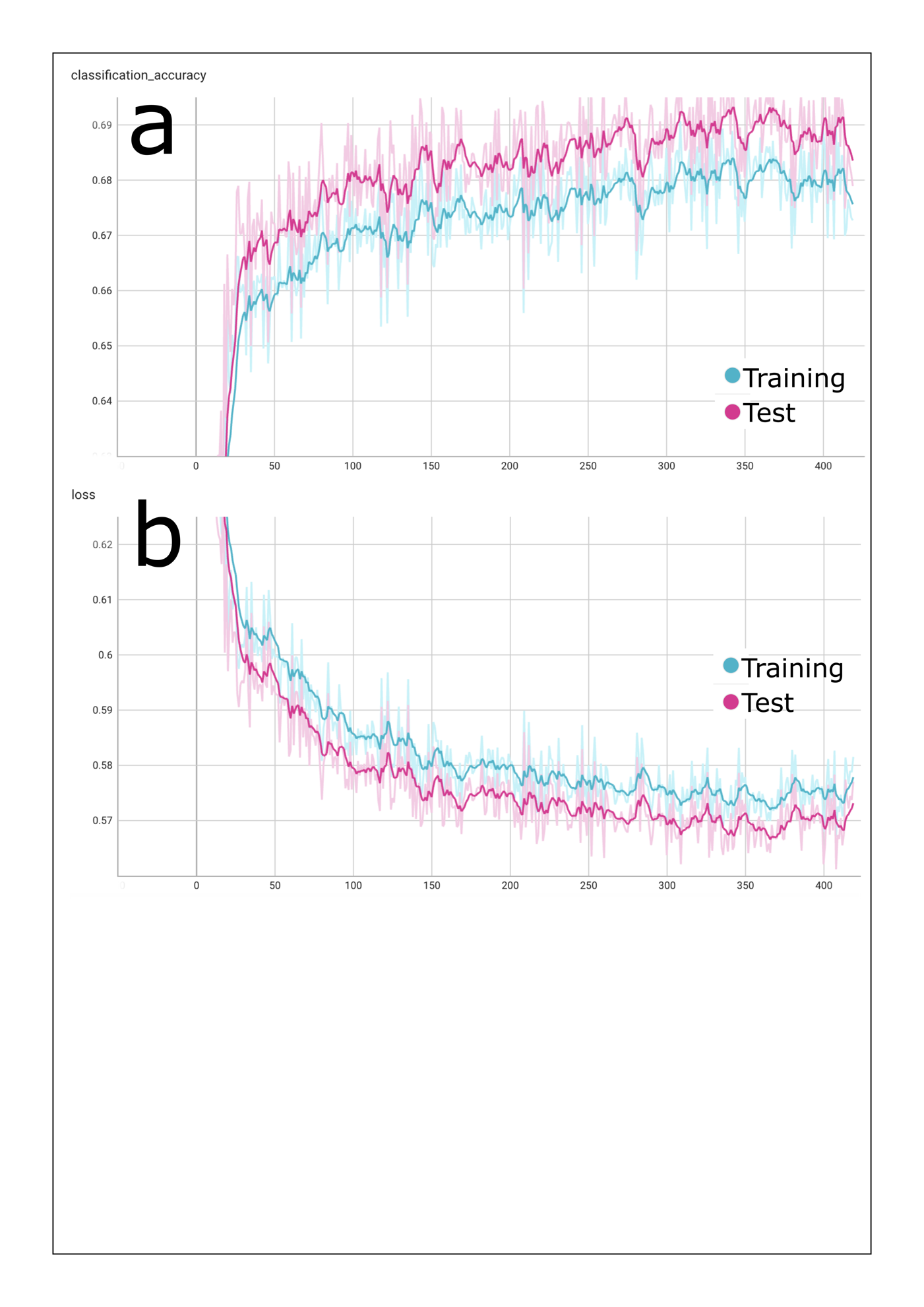

**a**) The classification accuracy (balanced accuracy) during the training procedure (5-fold cross-validation) for the training dataset and test dataset. **b**) The loss of the training and test datasets. Overall, we did not observe overfitting. The slightly lower test loss compared to training loss is likely not an issue and may be attributed to the use of dropout and early stopping criteria.

**eFigure 6: Using 5 MetBAGs to predict the incidence of ICD-based single disease entities in the UK Biobank**

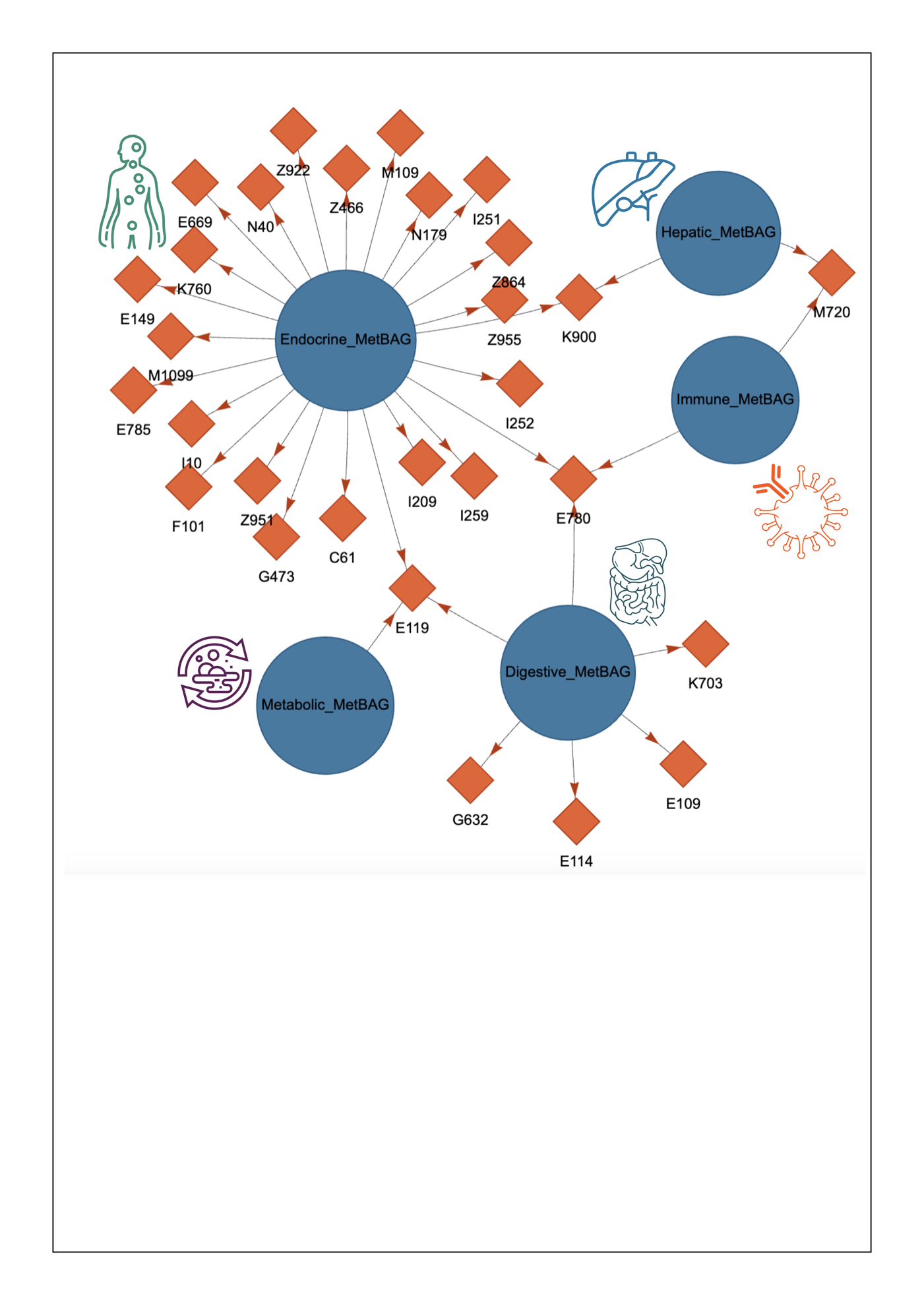

We defined the incidence of ICD-based disease entities using the ICD-10 code. Control participants were those without the disease diagnosis or any other disease diagnoses, while cases were individuals diagnosed with the disease after enrollment in the UK Biobank. We restricted our analyses for diseases that have at least 100 cases; significant results are presented after correcting for multiple comparisons using the Bonferroni correction (P-value < 0.05/304 disease endpoints). A Cox proportional hazards model was applied to derive all statistics, including P-values and hazard ratios, as shown in **eTable 10**. Additionally, we developed an interactive web link to display the results at: <https://labs-laboratory.com/medicine/metbag_de_sa>. Overall, we found that the endocrine MetBAG showed the most prominent prediction power with various diseases of the metabolic system (e.g., E669 for obesity), circulatory system (e.g., I209 for Angina pectoris). E119 (non-insulin-dependent diabetes mellitus) was linked to the metabolic, the digestive, and the endocrine BAGs. The disease entity was coded by ICD-10 in the UK Biobank: <https://biobank.ndph.ox.ac.uk/ukb/field.cgi?id=41270>.

**eTable 1: The characteristics of the MULTI consortium**

| Data type | Data type | Study | *N* | Age  [year (mean/std)] | | | | Sex (female) | | |
| --- | --- | --- | --- | --- | --- | --- | --- | --- | --- | --- |
|  |  |  |  | CN^c^ training/validation/test (*N*=29,354) | CN ind. test  (*N*=5000) | | PT (*N*=239,893) | CN training/validation/test (*N*=29,354) | CN ind. test (*N*=5000) | PT (*N*=239,893) |
| Individual-level | Plasma metabolomics/ Genetics | UKBB | 274,247 | 53.67±7.70^b^ | 53.76±7.69 | | 56.98±8.05 | 14,548 /51% | 2575/52% | 127,301/54% |
|  |  |  |  | 56.56±8.08 | | | | 147,994/54% | | |
| Summary-level | GWAS summary statistics | FinnGen | 521^a^ | NA | | NA | NA | NA | NA | NA |
|  |  | PGC | 4 ^a^ | NA | | NA | NA | NA | NA | NA |

^a^We only used the publicly available GWAS summary statistics from FinnGen and PGC for our Mendelian randomization analyses. Detailed information regarding each studies are presented online at <https://finngen.gitbook.io/documentation/data-download> for FinnGen, and <https://pgc.unc.edu/for-researchers/download-results/> for PGC.

^b^The exact training sample size for each organ may vary; we showed the number for the digestive MetBAG.

^c^CN was defined as participants with no recorded disease diagnoses following their enrollment in the UK Biobank study.

**eTable 2: The organ-specific metabolite annotations**

| **Metabolite** | **Protein** | **Organ** | **Description** | **Units** | **Group** | **Sub-group** | **Type** | **UKB Field ID** | **QC Flag Field ID** |
| --- | --- | --- | --- | --- | --- | --- | --- | --- | --- |
| bOHbutyrate | ANGPTL4 | Endocrine | 3-Hydroxybutyrate | mmol/L | Ketone bodies | NAN | Non-derived | 23474 | 23774 |
| Acetate | FGF21 | Hepatic | Acetate | mmol/L | Ketone bodies | NAN | Non-derived | 23475 | 23775 |
| Acetoacetate | ANGPTL4 | Endocrine | Acetoacetate | mmol/L | Ketone bodies | NAN | Non-derived | 23476 | 23776 |
| Acetone | ANGPTL4 | Endocrine | Acetone | mmol/L | Ketone bodies | NAN | Non-derived | 23477 | 23777 |
| Ala | IGFBP1 | Hepatic | Alanine | mmol/L | Amino acids | NAN | Non-derived | 23460 | 23760 |
| Albumin | CSF1 | Endocrine | Albumin | g/l | Fluid balance | NAN | Non-derived | 23479 | 23779 |
| ApoA1 | APOA1 | Hepatic | Apolipoprotein A1 | g/l | Apolipoproteins | NAN | Non-derived | 23440 | 23740 |
| ApoB | PLA2G7 | Immune | Apolipoprotein B | g/l | Apolipoproteins | NAN | Non-derived | 23439 | 23739 |
| HDL_size | PLTP | Endocrine | Average Diameter for HDL Particles | nm | Lipoprotein particle sizes | NAN | Non-derived | 23433 | 23733 |
| LDL_size | APOF | Hepatic | Average Diameter for LDL Particles | nm | Lipoprotein particle sizes | NAN | Non-derived | 23432 | 23732 |
| VLDL_size | LDLR | Endocrine | Average Diameter for VLDL Particles | nm | Lipoprotein particle sizes | NAN | Non-derived | 23431 | 23731 |
| XXL_VLDL_CE | LDLR | Endocrine | Cholesteryl Esters in Chylomicrons and Extremely Large VLDL | mmol/L | Lipoprotein subclasses | Chylomicrons and extremely large VLDL (particle diameters from 75 nm upwards) | Non-derived | 23485 | 23785 |
| IDL_CE | FGFBP1 | Digestive | Cholesteryl Esters in IDL | mmol/L | Lipoprotein subclasses | IDL (average diameter 28.6 nm) | Non-derived | 23527 | 23827 |
| L_HDL_CE | PLTP | Endocrine | Cholesteryl Esters in Large HDL | mmol/L | Lipoprotein subclasses | Large HDL (average diameter 12.1 nm) | Non-derived | 23562 | 23862 |
| L_LDL_CE | PLA2G7 | Immune | Cholesteryl Esters in Large LDL | mmol/L | Lipoprotein subclasses | Large LDL (average diameter 25.5 nm) | Non-derived | 23534 | 23834 |
| L_VLDL_CE | LDLR | Endocrine | Cholesteryl Esters in Large VLDL | mmol/L | Lipoprotein subclasses | Large VLDL (average diameter 53.6 nm) | Non-derived | 23499 | 23799 |
| M_HDL_CE | APOA1 | Hepatic | Cholesteryl Esters in Medium HDL | mmol/L | Lipoprotein subclasses | Medium HDL (average diameter 10.9 nm) | Non-derived | 23569 | 23869 |
| M_LDL_CE | PLA2G7 | Immune | Cholesteryl Esters in Medium LDL | mmol/L | Lipoprotein subclasses | Medium LDL (average diameter 23 nm) | Non-derived | 23541 | 23841 |
| M_VLDL_CE | FGFBP1 | Digestive | Cholesteryl Esters in Medium VLDL | mmol/L | Lipoprotein subclasses | Medium VLDL (average diameter 44.5 nm) | Non-derived | 23506 | 23806 |
| S_HDL_CE | APOM | Hepatic | Cholesteryl Esters in Small HDL | mmol/L | Lipoprotein subclasses | Small HDL (average diameter 8.7 nm) | Non-derived | 23576 | 23876 |
| S_LDL_CE | PLA2G7 | Immune | Cholesteryl Esters in Small LDL | mmol/L | Lipoprotein subclasses | Small LDL (average diameter 18.7 nm) | Non-derived | 23548 | 23848 |
| S_VLDL_CE | LDLR | Endocrine | Cholesteryl Esters in Small VLDL | mmol/L | Lipoprotein subclasses | Small VLDL (average diameter 36.8 nm) | Non-derived | 23513 | 23813 |
| XL_HDL_CE | ADIPOQ | Endocrine | Cholesteryl Esters in Very Large HDL | mmol/L | Lipoprotein subclasses | Very large HDL (average diameter 14.3 nm) | Non-derived | 23555 | 23855 |
| XL_VLDL_CE | LDLR | Endocrine | Cholesteryl Esters in Very Large VLDL | mmol/L | Lipoprotein subclasses | Very large VLDL (average diameter 64 nm) | Non-derived | 23492 | 23792 |
| XS_VLDL_CE | FGFBP1 | Digestive | Cholesteryl Esters in Very Small VLDL | mmol/L | Lipoprotein subclasses | Very small VLDL (average diameter 31.3 nm) | Non-derived | 23520 | 23820 |
| Citrate | ANGPTL4 | Endocrine | Citrate | mmol/L | Glycolysis related metabolites | NAN | Non-derived | 23473 | 23773 |
| Clinical_LDL_C | PLA2G7 | Immune | Clinical LDL Cholesterol | mmol/L | Cholesterol | NAN | Non-derived | 23404 | 23704 |
| XXL_VLDL_P | LDLR | Endocrine | Concentration of Chylomicrons and Extremely Large VLDL Particles | mmol/L | Lipoprotein subclasses | Chylomicrons and extremely large VLDL (particle diameters from 75 nm upwards) | Non-derived | 23481 | 23781 |
| IDL_P | PLA2G7 | Immune | Concentration of IDL Particles | mmol/L | Lipoprotein subclasses | IDL (average diameter 28.6 nm) | Non-derived | 23523 | 23823 |
| L_HDL_P | PLTP | Endocrine | Concentration of Large HDL Particles | mmol/L | Lipoprotein subclasses | Large HDL (average diameter 12.1 nm) | Non-derived | 23558 | 23858 |
| L_LDL_P | PLA2G7 | Immune | Concentration of Large LDL Particles | mmol/L | Lipoprotein subclasses | Large LDL (average diameter 25.5 nm) | Non-derived | 23530 | 23830 |
| L_VLDL_P | LDLR | Endocrine | Concentration of Large VLDL Particles | mmol/L | Lipoprotein subclasses | Large VLDL (average diameter 53.6 nm) | Non-derived | 23495 | 23795 |
| M_HDL_P | APOA1 | Hepatic | Concentration of Medium HDL Particles | mmol/L | Lipoprotein subclasses | Medium HDL (average diameter 10.9 nm) | Non-derived | 23565 | 23865 |
| M_LDL_P | PLA2G7 | Immune | Concentration of Medium LDL Particles | mmol/L | Lipoprotein subclasses | Medium LDL (average diameter 23 nm) | Non-derived | 23537 | 23837 |
| M_VLDL_P | LDLR | Endocrine | Concentration of Medium VLDL Particles | mmol/L | Lipoprotein subclasses | Medium VLDL (average diameter 44.5 nm) | Non-derived | 23502 | 23802 |
| S_HDL_P | LCAT | Hepatic | Concentration of Small HDL Particles | mmol/L | Lipoprotein subclasses | Small HDL (average diameter 8.7 nm) | Non-derived | 23572 | 23872 |
| S_LDL_P | PLA2G7 | Immune | Concentration of Small LDL Particles | mmol/L | Lipoprotein subclasses | Small LDL (average diameter 18.7 nm) | Non-derived | 23544 | 23844 |
| S_VLDL_P | LDLR | Endocrine | Concentration of Small VLDL Particles | mmol/L | Lipoprotein subclasses | Small VLDL (average diameter 36.8 nm) | Non-derived | 23509 | 23809 |
| XL_HDL_P | PLTP | Endocrine | Concentration of Very Large HDL Particles | mmol/L | Lipoprotein subclasses | Very large HDL (average diameter 14.3 nm) | Non-derived | 23551 | 23851 |
| XL_VLDL_P | LDLR | Endocrine | Concentration of Very Large VLDL Particles | mmol/L | Lipoprotein subclasses | Very large VLDL (average diameter 64 nm) | Non-derived | 23488 | 23788 |
| XS_VLDL_P | LDLR | Endocrine | Concentration of Very Small VLDL Particles | mmol/L | Lipoprotein subclasses | Very small VLDL (average diameter 31.3 nm) | Non-derived | 23516 | 23816 |
| Creatinine | IGFBP6 | CNS | Creatinine | mmol/L | Fluid balance | NAN | Non-derived | 23478 | 23778 |
| Unsaturation | PRAP1 | Digestive | Degree of Unsaturation | degree | Fatty acids | NAN | Non-derived | 23443 | 23743 |
| DHA | APOC1 | Hepatic | Docosahexaenoic Acid | mmol/L | Fatty acids | NAN | Non-derived | 23450 | 23750 |
| XXL_VLDL_FC | LDLR | Endocrine | Free Cholesterol in Chylomicrons and Extremely Large VLDL | mmol/L | Lipoprotein subclasses | Chylomicrons and extremely large VLDL (particle diameters from 75 nm upwards) | Non-derived | 23486 | 23786 |
| IDL_FC | FGFBP1 | Digestive | Free Cholesterol in IDL | mmol/L | Lipoprotein subclasses | IDL (average diameter 28.6 nm) | Non-derived | 23528 | 23828 |
| L_HDL_FC | PLTP | Endocrine | Free Cholesterol in Large HDL | mmol/L | Lipoprotein subclasses | Large HDL (average diameter 12.1 nm) | Non-derived | 23563 | 23863 |
| L_LDL_FC | FGFBP1 | Digestive | Free Cholesterol in Large LDL | mmol/L | Lipoprotein subclasses | Large LDL (average diameter 25.5 nm) | Non-derived | 23535 | 23835 |
| L_VLDL_FC | LDLR | Endocrine | Free Cholesterol in Large VLDL | mmol/L | Lipoprotein subclasses | Large VLDL (average diameter 53.6 nm) | Non-derived | 23500 | 23800 |
| M_HDL_FC | APOA1 | Hepatic | Free Cholesterol in Medium HDL | mmol/L | Lipoprotein subclasses | Medium HDL (average diameter 10.9 nm) | Non-derived | 23570 | 23870 |
| M_LDL_FC | PLA2G7 | Immune | Free Cholesterol in Medium LDL | mmol/L | Lipoprotein subclasses | Medium LDL (average diameter 23 nm) | Non-derived | 23542 | 23842 |
| M_VLDL_FC | PLA2G7 | Immune | Free Cholesterol in Medium VLDL | mmol/L | Lipoprotein subclasses | Medium VLDL (average diameter 44.5 nm) | Non-derived | 23507 | 23807 |
| S_HDL_FC | APOM | Hepatic | Free Cholesterol in Small HDL | mmol/L | Lipoprotein subclasses | Small HDL (average diameter 8.7 nm) | Non-derived | 23577 | 23877 |
| S_LDL_FC | PLA2G7 | Immune | Free Cholesterol in Small LDL | mmol/L | Lipoprotein subclasses | Small LDL (average diameter 18.7 nm) | Non-derived | 23549 | 23849 |
| S_VLDL_FC | PLA2G7 | Immune | Free Cholesterol in Small VLDL | mmol/L | Lipoprotein subclasses | Small VLDL (average diameter 36.8 nm) | Non-derived | 23514 | 23814 |
| XL_HDL_FC | ADIPOQ | Endocrine | Free Cholesterol in Very Large HDL | mmol/L | Lipoprotein subclasses | Very large HDL (average diameter 14.3 nm) | Non-derived | 23556 | 23856 |
| XL_VLDL_FC | LDLR | Endocrine | Free Cholesterol in Very Large VLDL | mmol/L | Lipoprotein subclasses | Very large VLDL (average diameter 64 nm) | Non-derived | 23493 | 23793 |
| XS_VLDL_FC | PLA2G7 | Immune | Free Cholesterol in Very Small VLDL | mmol/L | Lipoprotein subclasses | Very small VLDL (average diameter 31.3 nm) | Non-derived | 23521 | 23821 |
| Glucose | PLXNB2 | Digestive | Glucose | mmol/L | Glycolysis related metabolites | NAN | Non-derived | 23470 | 23770 |
| Gln | IGFBP2 | Digestive | Glutamine | mmol/L | Amino acids | NAN | Non-derived | 23461 | 23761 |
| Gly | LPL | Endocrine | Glycine | mmol/L | Amino acids | NAN | Non-derived | 23462 | 23762 |
| GlycA | RARRES2 | Endocrine | Glycoprotein Acetyls | mmol/L | Inflammation | NAN | Non-derived | 23480 | 23780 |
| His | FGL1 | Hepatic | Histidine | mmol/L | Amino acids | NAN | Non-derived | 23463 | 23763 |
| Ile | GCG | Digestive | Isoleucine | mmol/L | Amino acids | Branched-chain amino acids | Non-derived | 23465 | 23765 |
| Lactate | TNFSF14 | Hepatic | Lactate | mmol/L | Glycolysis related metabolites | NAN | Non-derived | 23471 | 23771 |
| Leu | GCG | Digestive | Leucine | mmol/L | Amino acids | Branched-chain amino acids | Non-derived | 23466 | 23766 |
| LA | APOM | Hepatic | Linoleic Acid | mmol/L | Fatty acids | NAN | Non-derived | 23449 | 23749 |
| MUFA | LDLR | Endocrine | Monounsaturated Fatty Acids | mmol/L | Fatty acids | NAN | Non-derived | 23447 | 23747 |
| Omega_3 | LDLR | Endocrine | Omega-3 Fatty Acids | mmol/L | Fatty acids | NAN | Non-derived | 23444 | 23744 |
| Omega_6 | APOC1 | Hepatic | Omega-6 Fatty Acids | mmol/L | Fatty acids | NAN | Non-derived | 23445 | 23745 |
| Phe | GCG | Digestive | Phenylalanine | mmol/L | Amino acids | Aromatic amino acids | Non-derived | 23468 | 23768 |
| Phosphatidylc | APOC1 | Hepatic | Phosphatidylcholines | mmol/L | Other lipids | NAN | Non-derived | 23437 | 23737 |
| Phosphoglyc | APOC1 | Hepatic | Phosphoglycerides | mmol/L | Other lipids | NAN | Non-derived | 23434 | 23734 |
| XXL_VLDL_PL | LDLR | Endocrine | Phospholipids in Chylomicrons and Extremely Large VLDL | mmol/L | Lipoprotein subclasses | Chylomicrons and extremely large VLDL (particle diameters from 75 nm upwards) | Non-derived | 23483 | 23783 |
| IDL_PL | FGFBP1 | Digestive | Phospholipids in IDL | mmol/L | Lipoprotein subclasses | IDL (average diameter 28.6 nm) | Non-derived | 23525 | 23825 |
| L_HDL_PL | PLTP | Endocrine | Phospholipids in Large HDL | mmol/L | Lipoprotein subclasses | Large HDL (average diameter 12.1 nm) | Non-derived | 23560 | 23860 |
| L_LDL_PL | PLA2G7 | Immune | Phospholipids in Large LDL | mmol/L | Lipoprotein subclasses | Large LDL (average diameter 25.5 nm) | Non-derived | 23532 | 23832 |
| L_VLDL_PL | LDLR | Endocrine | Phospholipids in Large VLDL | mmol/L | Lipoprotein subclasses | Large VLDL (average diameter 53.6 nm) | Non-derived | 23497 | 23797 |
| M_HDL_PL | APOA1 | Hepatic | Phospholipids in Medium HDL | mmol/L | Lipoprotein subclasses | Medium HDL (average diameter 10.9 nm) | Non-derived | 23567 | 23867 |
| M_LDL_PL | PLA2G7 | Immune | Phospholipids in Medium LDL | mmol/L | Lipoprotein subclasses | Medium LDL (average diameter 23 nm) | Non-derived | 23539 | 23839 |
| M_VLDL_PL | LDLR | Endocrine | Phospholipids in Medium VLDL | mmol/L | Lipoprotein subclasses | Medium VLDL (average diameter 44.5 nm) | Non-derived | 23504 | 23804 |
| S_HDL_PL | MAMDC4 | Hepatic | Phospholipids in Small HDL | mmol/L | Lipoprotein subclasses | Small HDL (average diameter 8.7 nm) | Non-derived | 23574 | 23874 |
| S_LDL_PL | PLA2G7 | Immune | Phospholipids in Small LDL | mmol/L | Lipoprotein subclasses | Small LDL (average diameter 18.7 nm) | Non-derived | 23546 | 23846 |
| S_VLDL_PL | LDLR | Endocrine | Phospholipids in Small VLDL | mmol/L | Lipoprotein subclasses | Small VLDL (average diameter 36.8 nm) | Non-derived | 23511 | 23811 |
| XL_HDL_PL | ADIPOQ | Endocrine | Phospholipids in Very Large HDL | mmol/L | Lipoprotein subclasses | Very large HDL (average diameter 14.3 nm) | Non-derived | 23553 | 23853 |
| XL_VLDL_PL | LDLR | Endocrine | Phospholipids in Very Large VLDL | mmol/L | Lipoprotein subclasses | Very large VLDL (average diameter 64 nm) | Non-derived | 23490 | 23790 |
| XS_VLDL_PL | LDLR | Endocrine | Phospholipids in Very Small VLDL | mmol/L | Lipoprotein subclasses | Very small VLDL (average diameter 31.3 nm) | Non-derived | 23518 | 23818 |
| Pyruvate | TNFSF14 | Hepatic | Pyruvate | mmol/L | Glycolysis related metabolites | NAN | Non-derived | 23472 | 23772 |
| SFA | LDLR | Endocrine | Saturated Fatty Acids | mmol/L | Fatty acids | NAN | Non-derived | 23448 | 23748 |
| Sphingomyelins | APOM | Hepatic | Sphingomyelins | mmol/L | Other lipids | NAN | Non-derived | 23438 | 23738 |
| Cholines | APOC1 | Hepatic | Total Cholines | mmol/L | Other lipids | NAN | Non-derived | 23436 | 23736 |
| XXL_VLDL_TG | LDLR | Endocrine | Triglycerides in Chylomicrons and Extremely Large VLDL | mmol/L | Lipoprotein subclasses | Chylomicrons and extremely large VLDL (particle diameters from 75 nm upwards) | Non-derived | 23487 | 23787 |
| IDL_TG | LDLR | Endocrine | Triglycerides in IDL | mmol/L | Lipoprotein subclasses | IDL (average diameter 28.6 nm) | Non-derived | 23529 | 23829 |
| L_HDL_TG | MFGE8 | Heart | Triglycerides in Large HDL | mmol/L | Lipoprotein subclasses | Large HDL (average diameter 12.1 nm) | Non-derived | 23564 | 23864 |
| L_LDL_TG | LDLR | Endocrine | Triglycerides in Large LDL | mmol/L | Lipoprotein subclasses | Large LDL (average diameter 25.5 nm) | Non-derived | 23536 | 23836 |
| L_VLDL_TG | LDLR | Endocrine | Triglycerides in Large VLDL | mmol/L | Lipoprotein subclasses | Large VLDL (average diameter 53.6 nm) | Non-derived | 23501 | 23801 |
| M_HDL_TG | LDLR | Endocrine | Triglycerides in Medium HDL | mmol/L | Lipoprotein subclasses | Medium HDL (average diameter 10.9 nm) | Non-derived | 23571 | 23871 |
| M_LDL_TG | LDLR | Endocrine | Triglycerides in Medium LDL | mmol/L | Lipoprotein subclasses | Medium LDL (average diameter 23 nm) | Non-derived | 23543 | 23843 |
| M_VLDL_TG | LDLR | Endocrine | Triglycerides in Medium VLDL | mmol/L | Lipoprotein subclasses | Medium VLDL (average diameter 44.5 nm) | Non-derived | 23508 | 23808 |
| S_HDL_TG | LDLR | Endocrine | Triglycerides in Small HDL | mmol/L | Lipoprotein subclasses | Small HDL (average diameter 8.7 nm) | Non-derived | 23578 | 23878 |
| S_LDL_TG | LDLR | Endocrine | Triglycerides in Small LDL | mmol/L | Lipoprotein subclasses | Small LDL (average diameter 18.7 nm) | Non-derived | 23550 | 23850 |
| S_VLDL_TG | LDLR | Endocrine | Triglycerides in Small VLDL | mmol/L | Lipoprotein subclasses | Small VLDL (average diameter 36.8 nm) | Non-derived | 23515 | 23815 |
| XL_HDL_TG | LDLR | Endocrine | Triglycerides in Very Large HDL | mmol/L | Lipoprotein subclasses | Very large HDL (average diameter 14.3 nm) | Non-derived | 23557 | 23857 |
| XL_VLDL_TG | LDLR | Endocrine | Triglycerides in Very Large VLDL | mmol/L | Lipoprotein subclasses | Very large VLDL (average diameter 64 nm) | Non-derived | 23494 | 23794 |
| XS_VLDL_TG | LDLR | Endocrine | Triglycerides in Very Small VLDL | mmol/L | Lipoprotein subclasses | Very small VLDL (average diameter 31.3 nm) | Non-derived | 23522 | 23822 |
| Tyr | GCG | Digestive | Tyrosine | mmol/L | Amino acids | Aromatic amino acids | Non-derived | 23469 | 23769 |
| Val | GCG | Digestive | Valine | mmol/L | Amino acids | Branched-chain amino acids | Non-derived | 23467 | 23767 |

**eTable 3: Biological age prediction performance before and after age bias correction**

A) Before age bias correction (**bolded results** are used for down-stream genetic analyses and prediction analyses):

| **BAG** | **MAE** | ***r*** | **Dataset** | **ML** |
| --- | --- | --- | --- | --- |
| Endocrine | 5.76822853 | 0.4345279 | Training/validation/test | LASSO |
| Endocrine | 11.1361923 | 0.3874523 | Ind. test | LASSO |
| Digestive | 6.15951989 | 0.3027713 | Training/validation/test | LASSO |
| Digestive | 6.55314784 | 0.30249991 | Ind. test | LASSO |
| Hepatic | **6.13720861** | 0.3099576 | Training/validation/test | LASSO |
| Hepatic | **6.57808892** | 0.25613969 | Ind. test | LASSO |
| Immune | 6.18216096 | 0.29126101 | Training/validation/test | LASSO |
| Immune | 8.73888796 | 0.28528541 | Ind. test | LASSO |
| Metabolic | **5.48790561** | 0.50486541 | Training/validation/test | LASSO |
| Metabolic | **6.09364695** | 0.35733733 | Ind. test | LASSO |
| Endocrine | **5.79719047** | 0.41483156 | Training/validation | NN |
| Endocrine | **7.15260623** | 0.42018615 | Ind. test | NN |
| Digestive | **6.1116544** | 0.31273247 | Training/validation | NN |
| Digestive | **6.48912378** | 0.29448793 | Ind. test | NN |
| Hepatic | 6.18080543 | 0.27668762 | Training/validation | NN |
| Hepatic | 11.7005039 | 0.23945256 | Ind. test | NN |
| Immune | **6.2644537** | 0.2454384 | Training/validation | NN |
| Immune | **6.5868877** | 0.24823601 | Ind. test | NN |
| Metabolic | 5.49739664 | 0.4925019 | Training/validation | NN |
| Metabolic | 7.67826195 | 0.42878154 | Ind. test | NN |

B) After age bias correction:

| **BAG** | **MAE** | ***r*** | **Dataset** | **ML** |
| --- | --- | --- | --- | --- |
| Endocrine | 2.27333859 | 0.93545843 | Training/validation/test | LASSO |
| Endocrine | 2.36709351 | 0.92743513 | Ind. test | LASSO |
| Digestive | 1.7362041 | 0.96155521 | Training/validation/test | LASSO |
| Digestive | 1.92129559 | 0.95248095 | Ind. test | LASSO |
| Hepatic | 1.70730603 | 0.96145802 | Training/validation/test | LASSO |
| Hepatic | 3.58234894 | 0.86215561 | Ind. test | LASSO |
| Immune | 1.64198089 | 0.9642867 | Training/validation/test | LASSO |
| Immune | 1.82053834 | 0.95719061 | Ind. test | LASSO |
| Metabolic | 2.60134806 | 0.91863681 | Training/validation/test | LASSO |
| Metabolic | 3.41933343 | 0.86883836 | Ind. test | LASSO |
| Endocrine | 2.68836515 | 0.91299422 | Training/validation | NN |
| Endocrine | 2.92294245 | 0.89631842 | Ind. test | NN |
| Digestive | 2.18673886 | 0.94084957 | Training/validation | NN |
| Digestive | 2.45744283 | 0.92545238 | Ind. test | NN |
| Hepatic | 2.02642324 | 0.94679575 | Training/validation | NN |
| Hepatic | 4.46778082 | 0.80936998 | Ind. test | NN |
| Immune | 1.67396695 | 0.96308709 | Training/validation | NN |
| Immune | 1.76094423 | 0.95902129 | Ind. test | NN |
| Metabolic | 2.91046033 | 0.90158848 | Training/validation | NN |
| Metabolic | 3.99673387 | 0.83362258 | Ind. test | NN |

C) **Before age bias correction for the PT group:**

| **BAG** | **MAE** | ***r*** | **Dataset** | **ML** |
| --- | --- | --- | --- | --- |
| Endocrine | 16.9924811 | 0.36627929 | PT | LASSO |
| Digestive | 7.36681321 | 0.21522594 | PT | LASSO |
| Hepatic | 8.48245909 | 0.18390403 | PT | LASSO |
| Immune | 15.6766426 | 0.21023532 | PT | LASSO |
| Metabolic | 22.6202452 | 0.41893355 | PT | LASSO |
| Endocrine | 11.7906443 | 0.34458636 | PT | NN |
| Digestive | 8.07142565 | 0.23386993 | PT | NN |
| Hepatic | 7.66370614 | 0.19371896 | PT | NN |
| Immune | 9.41591855 | 0.14578841 | PT | NN |
| Metabolic | 14.6478198 | 0.42748563 | PT | NN |

**eTable 4: Genomic loci of the 5 MetBAG GWASs**

| **TopLeadSNP** | **Chromosome** | **Position** | **P-value** | **Phenotype** | **Cytogenetic_region** |
| --- | --- | --- | --- | --- | --- |
| rs79598313 | 1 | 27284913 | 7.10E-11 | Endocrine | 1p36.11 |
| rs79439217 | 1 | 63060037 | 1.36E-112 | Endocrine | 1p31.3 |
| rs607518 | 1 | 150954671 | 1.78E-09 | Endocrine | 1q21.3 |
| rs11589479 | 1 | 155033308 | 3.05E-15 | Endocrine | 1q22 |
| rs1801274 | 1 | 161479745 | 1.51E-14 | Endocrine | 1q23.3 |
| rs61830291 | 1 | 221001142 | 2.57E-10 | Endocrine | 1q41 |
| rs11899121 | 2 | 20367973 | 5.82E-17 | Endocrine | 2p24.1 |
| 2:21398970_AT_A | 2 | 21398970 | 3.30E-38 | Endocrine | 2p24.1 |
| rs1260326 | 2 | 27730940 | 3.10E-182 | Endocrine | 2p23.3 |
| rs72926946 | 2 | 203477868 | 5.37E-09 | Endocrine | 2q33.2 |
| rs1047891 | 2 | 211540507 | 1.06E-68 | Endocrine | 2q34 |
| rs531842900 | 2 | 219460127 | 4.71E-14 | Endocrine | 2q35 |
| rs6775191 | 3 | 12272199 | 1.83E-10 | Endocrine | 3p25.2 |
| rs115744844 | 3 | 48632756 | 1.15E-15 | Endocrine | 3p21.31 |
| rs78946096 | 3 | 132188163 | 2.01E-10 | Endocrine | 3q22.1 |
| rs2857984 | 4 | 3490072 | 1.77E-14 | Endocrine | 4p16.3 |
| rs77849807 | 4 | 26152727 | 8.08E-18 | Endocrine | 4p15.2 |
| rs9884390 | 4 | 69373407 | 5.56E-21 | Endocrine | 4q13.2 |
| rs55772354 | 4 | 74276150 | 2.05E-11 | Endocrine | 4q13.3 |
| rs2035403 | 4 | 88018991 | 5.73E-09 | Endocrine | 4q22.1 |
| rs3811741 | 4 | 128803159 | 8.65E-10 | Endocrine | 4q28.2 |
| rs2921604 | 5 | 14867948 | 5.24E-13 | Endocrine | 5p15.2 |
| rs28650790 | 5 | 55861464 | 5.38E-10 | Endocrine | 5q11.2 |
| rs72801474 | 5 | 132444128 | 3.51E-09 | Endocrine | 5q31.1 |
| rs6882076 | 5 | 156390297 | 8.89E-27 | Endocrine | 5q33.3 |
| rs200977 | 6 | 27854301 | 1.31E-15 | Endocrine | 6p22.1 |
| rs114863007 | 6 | 34729158 | 2.16E-09 | Endocrine | 6p21.31 |
| rs6905288 | 6 | 43758873 | 4.88E-10 | Endocrine | 6p21.1 |
| rs2068408 | 6 | 127227414 | 5.39E-10 | Endocrine | 6q22.33 |
| rs7775698 | 6 | 135418635 | 8.97E-15 | Endocrine | 6q23.3 |
| rs199607859 | 6 | 139835418 | 3.74E-10 | Endocrine | 6q24.1 |
| rs140570886 | 6 | 161013013 | 5.42E-26 | Endocrine | 6q26 |
| rs11764937 | 7 | 1092074 | 2.42E-10 | Endocrine | 7p22.3 |
| rs62442540 | 7 | 2352693 | 7.50E-10 | Endocrine | 7p22.3 |
| rs4722551 | 7 | 25991826 | 4.77E-14 | Endocrine | 7p15.2 |
| rs17145750 | 7 | 73026378 | 2.57E-88 | Endocrine | 7q11.23 |
| rs4841132 | 8 | 9183596 | 3.27E-21 | Endocrine | 8p23.1 |
| rs17810889 | 8 | 11623236 | 4.84E-15 | Endocrine | 8p23.1 |
| rs146812806 | 8 | 18272503 | 8.09E-26 | Endocrine | 8p22 |
| rs117199990 | 8 | 19820916 | 2.80E-50 | Endocrine | 8p21.3 |
| rs2737265 | 8 | 116667634 | 7.20E-14 | Endocrine | 8q23.3 |
| rs2954021 | 8 | 126482077 | 1.02E-109 | Endocrine | 8q24.13 |
| rs820503 | 9 | 6667928 | 1.53E-10 | Endocrine | 9p24.1 |
| rs686030 | 9 | 15304782 | 3.77E-21 | Endocrine | 9p22.3 |
| rs2740488 | 9 | 107661742 | 6.63E-41 | Endocrine | 9q31.1 |
| 9:136138765_GCGCCCACCACTA_G | 9 | 136138765 | 1.37E-10 | Endocrine | 9q34.2 |
| 10:5246753_CACAGAT_C | 10 | 5246753 | 4.38E-16 | Endocrine | 10p15.1 |
| rs56278466 | 10 | 17875857 | 9.03E-12 | Endocrine | 10p12.33 |
| rs34370527 | 10 | 93614675 | 4.61E-11 | Endocrine | 10q23.32 |
| rs2068888 | 10 | 94839642 | 1.07E-13 | Endocrine | 10q23.33 |
| rs174582 | 11 | 61607168 | 7.55897e-310 | Endocrine | 11q12.2 |
| rs2229738 | 11 | 68562328 | 9.55E-28 | Endocrine | 11q13.3 |
| rs499974 | 11 | 75455021 | 2.33E-37 | Endocrine | 11q13.5 |
| rs964184 | 11 | 116648917 | 9.54E-263 | Endocrine | 11q23.3 |
| rs4764939 | 12 | 103522952 | 9.93E-13 | Endocrine | 12q23.2 |
| rs7979473 | 12 | 121420260 | 4.09E-23 | Endocrine | 12q24.31 |
| rs7140110 | 13 | 114544024 | 3.78E-15 | Endocrine | 13q34 |
| rs28929474 | 14 | 94844947 | 5.14E-185 | Endocrine | 14q32.13 |
| rs261334 | 15 | 58726744 | 5.04751e-314 | Endocrine | 15q21.3 |
| rs10444863 | 15 | 78358837 | 9.61E-11 | Endocrine | 15q25.1 |
| rs8025505 | 15 | 102067841 | 1.67E-09 | Endocrine | 15q26.3 |
| rs2269558 | 16 | 682250 | 3.05E-10 | Endocrine | 16p13.3 |
| rs11644601 | 16 | 15172118 | 9.58E-49 | Endocrine | 16p13.11 |
| rs183130 | 16 | 56991363 | 1.76E-87 | Endocrine | 16q13 |
| rs4986970 | 16 | 67976320 | 2.02E-09 | Endocrine | 16q22.1 |
| rs75448233 | 17 | 6611798 | 1.07E-17 | Endocrine | 17p13.1 |
| rs72829446 | 17 | 7552123 | 4.16E-11 | Endocrine | 17p13.1 |
| rs854784 | 17 | 18040690 | 5.71E-11 | Endocrine | 17p11.2 |
| rs62074055 | 17 | 45771933 | 3.43E-12 | Endocrine | 17q21.32 |
| rs537926683 | 17 | 66003541 | 2.84E-10 | Endocrine | 17q24.2 |
| rs77542162 | 17 | 67081278 | 6.03E-14 | Endocrine | 17q24.2 |
| rs75056343 | 17 | 73758204 | 9.74E-10 | Endocrine | 17q25.1 |
| 18:9712723_GTGCGTGCACTTAAAAAC_G | 18 | 9712723 | 1.40E-10 | Endocrine | 18p11.22 |
| rs77960347 | 18 | 47109955 | 7.08E-38 | Endocrine | 18q21.1 |
| rs116843064 | 19 | 8429323 | 5.15E-24 | Endocrine | 19p13.2 |
| rs17699030 | 19 | 11330942 | 2.65E-13 | Endocrine | 19p13.2 |
| rs58542926 | 19 | 19379549 | 3.75E-109 | Endocrine | 19p13.11 |
| rs584007 | 19 | 45416478 | 5.43E-37 | Endocrine | 19q13.32 |
| rs113886122 | 19 | 50044741 | 2.15E-47 | Endocrine | 19q13.33 |
| rs7261820 | 20 | 34160840 | 2.25E-10 | Endocrine | 20q11.22 |
| rs1883711 | 20 | 39179822 | 7.16E-09 | Endocrine | 20q12 |
| rs6129767 | 20 | 39822332 | 1.90E-10 | Endocrine | 20q12 |
| rs1800961 | 20 | 43042364 | 4.59E-15 | Endocrine | 20q13.12 |
| rs6073958 | 20 | 44551855 | 2.30E-57 | Endocrine | 20q13.12 |
| rs1206540 | 22 | 19235363 | 4.71E-11 | Endocrine | 22q11.21 |
| rs4821124 | 22 | 21979289 | 2.00E-09 | Endocrine | 22q11.21 |
| 22:24250072_AG_A | 22 | 24250072 | 3.62E-11 | Endocrine | 22q11.23 |
| rs117563943 | 22 | 50114083 | 5.33E-09 | Endocrine | 22q13.33 |
| rs11591147 | 1 | 55505647 | 4.11E-49 | Digestive | 1p32.3 |
| rs6682423 | 1 | 63171063 | 5.06E-29 | Digestive | 1p31.3 |
| rs1730850 | 1 | 107597988 | 1.38E-12 | Digestive | 1p13.3 |
| rs12740374 | 1 | 109817590 | 4.10E-51 | Digestive | 1p13.3 |
| rs10923358 | 1 | 118164794 | 7.80E-09 | Digestive | 1p12 |
| rs79687284 | 1 | 214150821 | 2.14E-18 | Digestive | 1q32.3 |
| rs2642438 | 1 | 220970028 | 1.45E-15 | Digestive | 1q41 |
| rs4846922 | 1 | 230307182 | 3.10E-13 | Digestive | 1q42.13 |
| rs907866 | 2 | 20371380 | 4.59E-21 | Digestive | 2p24.1 |
| rs312944 | 2 | 21325188 | 1.10E-57 | Digestive | 2p24.1 |
| rs1260326 | 2 | 27730940 | 1.33E-55 | Digestive | 2p23.3 |
| rs4299376 | 2 | 44072576 | 2.62E-23 | Digestive | 2p21 |
| rs2215870 | 2 | 63132342 | 3.44E-10 | Digestive | 2p15 |
| rs6759692 | 2 | 101739211 | 7.39E-09 | Digestive | 2q11.2 |
| rs10198390 | 2 | 191706712 | 1.24E-17 | Digestive | 2q32.2 |
| rs62189001 | 2 | 226905094 | 4.88E-11 | Digestive | 2q36.3 |
| rs11708067 | 3 | 123065778 | 2.19E-25 | Digestive | 3q21.1 |
| rs9840812 | 3 | 135843162 | 7.25E-14 | Digestive | 3q22.3 |
| rs11706810 | 3 | 160159921 | 1.15E-19 | Digestive | 3q25.33 |
| rs35662434 | 4 | 69339933 | 6.89E-22 | Digestive | 4q13.2 |
| rs116564150 | 4 | 89199397 | 8.24E-37 | Digestive | 4q22.1 |
| rs2602836 | 4 | 100014805 | 8.29E-36 | Digestive | 4q23 |
| rs13107325 | 4 | 103188709 | 1.46E-12 | Digestive | 4q24 |
| rs12916 | 5 | 74656539 | 5.35E-26 | Digestive | 5q13.3 |
| rs55776147 | 5 | 156400808 | 5.54E-10 | Digestive | 5q33.3 |
| rs12206654 | 6 | 111556834 | 1.11E-118 | Digestive | 6q21 |
| rs12662901 | 6 | 116305637 | 2.94E-10 | Digestive | 6q22.1 |
| rs10455872 | 6 | 161010118 | 9.67E-22 | Digestive | 6q26 |
| rs9769088 | 7 | 1029585 | 2.67E-36 | Digestive | 7p22.3 |
| rs73091233 | 7 | 28210687 | 1.09E-14 | Digestive | 7p15.1 |
| rs217358 | 7 | 44622286 | 4.45E-09 | Digestive | 7p13 |
| rs35332062 | 7 | 73012042 | 1.32E-19 | Digestive | 7q11.23 |
| rs10953301 | 7 | 100264893 | 1.49E-12 | Digestive | 7q22.1 |
| rs35614264 | 8 | 9185533 | 2.90E-13 | Digestive | 8p23.1 |
| rs2720587 | 8 | 17378468 | 2.39E-10 | Digestive | 8p22 |
| rs80302977 | 8 | 19934339 | 4.51E-10 | Digestive | 8p21.3 |
| rs4876611 | 8 | 116671848 | 1.95E-16 | Digestive | 8q23.3 |
| rs11558471 | 8 | 118185733 | 4.06E-16 | Digestive | 8q24.11 |
| rs2954021 | 8 | 126482077 | 9.22E-72 | Digestive | 8q24.13 |
| rs385893 | 9 | 4763176 | 1.07E-09 | Digestive | 9p24.1 |
| rs10811661 | 9 | 22134094 | 4.82E-11 | Digestive | 9p21.3 |
| rs1883025 | 9 | 107664301 | 2.62E-16 | Digestive | 9q31.1 |
| 9:136138765_GCGCCCACCACTA_G | 9 | 136138765 | 9.20E-49 | Digestive | 9q34.2 |
| 10:65226769_TA_T | 10 | 65226769 | 2.72E-63 | Digestive | 10q21.3 |
| rs17096421 | 10 | 88820592 | 8.88E-09 | Digestive | 10q23.2 |
| rs2297991 | 10 | 113913222 | 8.74E-13 | Digestive | 10q25.2 |
| rs10766473 | 11 | 18401531 | 3.52E-11 | Digestive | 11p15.1 |
| rs174592 | 11 | 61618608 | 2.84352e-317 | Digestive | 11q12.2 |
| rs2229738 | 11 | 68562328 | 1.78E-16 | Digestive | 11q13.3 |
| rs499974 | 11 | 75455021 | 2.06E-15 | Digestive | 11q13.5 |
| rs1871395 | 12 | 21352315 | 3.39E-15 | Digestive | 12p12.1 |
| rs11047639 | 12 | 24951488 | 2.62E-09 | Digestive | 12p12.1 |
| rs79295634 | 12 | 47180008 | 1.80E-34 | Digestive | 12q13.11 |
| rs2657878 | 12 | 56866962 | 4.64E-14 | Digestive | 12q13.3 |
| rs17122673 | 12 | 59990767 | 1.88E-12 | Digestive | 12q14.1 |
| rs991817 | 12 | 111410537 | 9.90E-10 | Digestive | 12q24.11 |
| rs2393775 | 12 | 121424574 | 2.16E-108 | Digestive | 12q24.31 |
| rs372273603 | 12 | 122289655 | 1.41E-12 | Digestive | 12q24.31 |
| rs112368163 | 12 | 133171120 | 3.19E-09 | Digestive | 12q24.33 |
| rs150641790 | 13 | 41687844 | 7.13E-14 | Digestive | 13q14.11 |
| rs34160920 | 14 | 69264564 | 7.35E-11 | Digestive | 14q24.1 |
| 14:102703016_ACAC_A | 14 | 102703016 | 4.47E-13 | Digestive | 14q32.31 |
| rs28442086 | 15 | 58705036 | 5e-324 | Digestive | 15q21.3 |
| rs340005 | 15 | 60878030 | 9.06E-09 | Digestive | 15q22.2 |
| rs72789541 | 16 | 15127534 | 1.67E-34 | Digestive | 16p13.11 |
| rs150851429 | 16 | 71625831 | 1.35E-60 | Digestive | 16q22.2 |
| rs4575545 | 16 | 79755446 | 1.79E-10 | Digestive | 16q23.2 |
| rs149822599 | 17 | 7169299 | 3.38E-10 | Digestive | 17p13.1 |
| rs77697917 | 17 | 41840849 | 3.39E-15 | Digestive | 17q21.31 |
| 17:44204299_AC_A | 17 | 44204299 | 4.64E-14 | Digestive | 17q21.31 |
| 17:44204299_AC_A | 17 | 44204299 | 4.64E-14 | Digestive | 17q21.31 |
| rs4264433 | 17 | 45737275 | 1.43E-18 | Digestive | 17q21.32 |
| rs1801689 | 17 | 64210580 | 1.04E-27 | Digestive | 17q24.2 |
| rs77542162 | 17 | 67081278 | 4.98E-38 | Digestive | 17q24.2 |
| rs77960347 | 18 | 47109955 | 1.95E-30 | Digestive | 18q21.1 |
| rs61194703 | 19 | 11192193 | 5.22E-108 | Digestive | 19p13.2 |
| rs1065853 | 19 | 45413233 | 9.96E-193 | Digestive | 19q13.32 |
| rs545587 | 19 | 49319664 | 3.48E-21 | Digestive | 19q13.33 |
| rs4806498 | 19 | 54674742 | 1.34E-11 | Digestive | 19q13.42 |
| rs6060491 | 20 | 34175100 | 1.41E-12 | Digestive | 20q11.22 |
| rs1883711 | 20 | 39179822 | 3.68E-20 | Digestive | 20q12 |
| rs2866372 | 20 | 39841091 | 4.63E-10 | Digestive | 20q12 |
| rs1800961 | 20 | 43042364 | 7.49E-14 | Digestive | 20q13.12 |
| rs111602331 | 20 | 44557474 | 4.96E-64 | Digestive | 20q13.12 |
| rs3747207 | 22 | 44324855 | 2.78E-21 | Digestive | 22q13.31 |
| rs66731853 | 1 | 20916238 | 4.13E-09 | Hepatic | 1p36.12 |
| rs11206189 | 1 | 39964974 | 7.37E-12 | Hepatic | 1p34.3 |
| rs11591147 | 1 | 55505647 | 6.26E-14 | Hepatic | 1p32.3 |
| 1:62909949_AAAG_A | 1 | 62909949 | 3.09E-116 | Hepatic | 1p31.3 |
| rs660240 | 1 | 109817838 | 4.15E-15 | Hepatic | 1p13.3 |
| rs198325 | 1 | 150937405 | 4.75E-23 | Hepatic | 1q21.3 |
| rs9426830 | 1 | 154594690 | 5.35E-10 | Hepatic | 1q21.3 |
| rs6671847 | 1 | 161478810 | 1.05E-09 | Hepatic | 1q23.3 |
| rs4846918 | 1 | 230300586 | 5.20E-25 | Hepatic | 1q42.13 |
| rs558971 | 1 | 234853406 | 2.36E-29 | Hepatic | 1q42.3 |
| rs907868 | 2 | 20371167 | 2.91E-24 | Hepatic | 2p24.1 |
| rs544450 | 2 | 21384358 | 8.18E-37 | Hepatic | 2p24.1 |
| rs11691698 | 2 | 26642190 | 4.73E-12 | Hepatic | 2p23.3 |
| rs11691698 | 2 | 26642190 | 4.73E-12 | Hepatic | 2p23.3 |
| rs1260326 | 2 | 27730940 | 1.70E-136 | Hepatic | 2p23.3 |
| rs1861399 | 2 | 64906787 | 2.68E-12 | Hepatic | 2p14 |
| rs11693150 | 2 | 203496575 | 8.50E-09 | Hepatic | 2q33.2 |
| rs17036170 | 3 | 12330411 | 1.19E-11 | Hepatic | 3p25.2 |
| rs76061303 | 3 | 48609041 | 5.38E-13 | Hepatic | 3p21.31 |
| rs2461824 | 3 | 119530027 | 9.73E-09 | Hepatic | 3q13.33 |
| rs80080062 | 3 | 185812169 | 1.08E-10 | Hepatic | 3q27.2 |
| rs139750425 | 4 | 3470174 | 5.03E-13 | Hepatic | 4p16.3 |
| rs9884390 | 4 | 69373407 | 3.13E-29 | Hepatic | 4q13.2 |
| rs13107325 | 4 | 103188709 | 1.59E-21 | Hepatic | 4q24 |
| rs76967117 | 6 | 34603691 | 1.93E-13 | Hepatic | 6p21.31 |
| rs9462860 | 6 | 42947013 | 1.29E-10 | Hepatic | 6p21.1 |
| rs12662901 | 6 | 116305637 | 4.04E-09 | Hepatic | 6q22.1 |
| rs10282584 | 7 | 1081442 | 2.53E-22 | Hepatic | 7p22.3 |
| rs836545 | 7 | 6479410 | 1.24E-15 | Hepatic | 7p22.1 |
| rs876039 | 7 | 50308811 | 3.83E-09 | Hepatic | 7p12.2 |
| rs13234805 | 7 | 73049182 | 1.17E-27 | Hepatic | 7q11.23 |
| rs141370588 | 7 | 155043252 | 7.96E-11 | Hepatic | 7q36.2 |
| rs9987289 | 8 | 9183358 | 1.74E-65 | Hepatic | 8p23.1 |
| rs804267 | 8 | 11629241 | 1.59E-16 | Hepatic | 8p23.1 |
| rs35246381 | 8 | 18272535 | 1.07E-10 | Hepatic | 8p22 |
| rs15285 | 8 | 19824667 | 2.49E-31 | Hepatic | 8p21.3 |
| rs2245221 | 8 | 116624879 | 8.70E-17 | Hepatic | 8q23.3 |
| rs2954021 | 8 | 126482077 | 4.71E-63 | Hepatic | 8q24.13 |
| rs7460860 | 8 | 144304941 | 2.97E-17 | Hepatic | 8q24.3 |
| rs12685293 | 9 | 6667421 | 6.58E-11 | Hepatic | 9p24.1 |
| rs1215112 | 9 | 15303583 | 2.60E-31 | Hepatic | 9p22.3 |
| rs2740488 | 9 | 107661742 | 1.45E-117 | Hepatic | 9q31.1 |
| 9:136151579_TGGTGCAGGCGCAGGAAAAAATTGTGGCAATTCCTCA_T | 9 | 136151579 | 1.31E-29 | Hepatic | 9q34.2 |
| rs34538474 | 10 | 3139540 | 3.40E-13 | Hepatic | 10p15.2 |
| 10:5246753_CACAGAT_C | 10 | 5246753 | 2.34E-22 | Hepatic | 10p15.1 |
| rs56278466 | 10 | 17875857 | 1.97E-10 | Hepatic | 10p12.33 |
| rs61854123 | 10 | 46065451 | 4.88E-16 | Hepatic | 10q11.21 |
| 10:65265705_CA_C | 10 | 65265705 | 9.74E-25 | Hepatic | 10q21.3 |
| rs200811093 | 10 | 88238881 | 2.02E-09 | Hepatic | 10q23.2 |
| rs112468457 | 10 | 102036074 | 2.08E-10 | Hepatic | 10q24.31 |
| rs10787429 | 10 | 113949664 | 6.06E-10 | Hepatic | 10q25.2 |
| rs80235628 | 10 | 122859270 | 5.51E-11 | Hepatic | 10q26.12 |
| rs2930196 | 11 | 48048802 | 3.98E-13 | Hepatic | 11p11.2 |
| rs174462 | 11 | 61657666 | 7.60E-308 | Hepatic | 11q12.2 |
| rs2229738 | 11 | 68562328 | 6.65E-40 | Hepatic | 11q13.3 |
| rs600518 | 11 | 75455373 | 5.88E-72 | Hepatic | 11q13.5 |
| rs12721030 | 11 | 116705278 | 7.18E-121 | Hepatic | 11q23.3 |
| rs752780106 | 12 | 21323338 | 6.90E-11 | Hepatic | 12p12.1 |
| rs10783367 | 12 | 50915436 | 2.78E-09 | Hepatic | 12q13.12 |
| rs2058804 | 12 | 109909011 | 9.08E-11 | Hepatic | 12q24.11 |
| rs10744774 | 12 | 112090022 | 1.54E-09 | Hepatic | 12q24.12 |
| rs7979473 | 12 | 121420260 | 3.83E-19 | Hepatic | 12q24.31 |
| rs4759377 | 12 | 123796849 | 4.33E-12 | Hepatic | 12q24.31 |
| rs10773112 | 12 | 125338529 | 2.46E-14 | Hepatic | 12q24.31 |
| rs10483863 | 14 | 75322327 | 3.37E-09 | Hepatic | 14q24.3 |
| rs28929474 | 14 | 94844947 | 2.58E-14 | Hepatic | 14q32.13 |
| rs11848805 | 14 | 105255658 | 2.79E-11 | Hepatic | 14q32.33 |
| rs397923 | 15 | 58692118 | 4.07994e-319 | Hepatic | 15q21.3 |
| rs11075253 | 16 | 15148646 | 3.54E-112 | Hepatic | 16p13.11 |
| rs118146573 | 16 | 57000938 | 6.80E-258 | Hepatic | 16q13 |
| rs4986970 | 16 | 67976320 | 1.72E-15 | Hepatic | 16q22.1 |
| rs11078597 | 17 | 1618363 | 3.98E-09 | Hepatic | 17p13.3 |
| rs1794287 | 17 | 7578837 | 3.65E-09 | Hepatic | 17p13.1 |
| rs2168345 | 17 | 17419338 | 1.34E-09 | Hepatic | 17p11.2 |
| rs241780 | 17 | 26628747 | 3.28E-14 | Hepatic | 17q11.2 |
| rs11078931 | 17 | 38147294 | 1.66E-15 | Hepatic | 17q21.1 |
| rs72836561 | 17 | 41926126 | 4.12E-18 | Hepatic | 17q21.31 |
| rs62074055 | 17 | 45771933 | 1.76E-17 | Hepatic | 17q21.32 |
| rs34931250 | 17 | 66879927 | 1.36E-11 | Hepatic | 17q24.2 |
| rs4129767 | 17 | 76403984 | 8.20E-12 | Hepatic | 17q25.3 |
| rs3934427 | 18 | 2979828 | 3.97E-09 | Hepatic | 18p11.31 |
| rs34250533 | 18 | 19565847 | 4.23E-12 | Hepatic | 18q11.2 |
| rs77960347 | 18 | 47109955 | 2.09E-138 | Hepatic | 18q21.1 |
| rs12976739 | 19 | 8461663 | 8.61E-12 | Hepatic | 19p13.2 |
| rs17699030 | 19 | 11330942 | 2.89E-40 | Hepatic | 19p13.2 |
| rs58542926 | 19 | 19379549 | 4.69E-50 | Hepatic | 19p13.11 |
| rs5167 | 19 | 45448465 | 1.18E-44 | Hepatic | 19q13.32 |
| rs34514836 | 19 | 46385438 | 2.36E-11 | Hepatic | 19q13.32 |
| rs34010237 | 19 | 50012574 | 2.10E-13 | Hepatic | 19q13.33 |
| rs11435987 | 19 | 54799692 | 3.09E-17 | Hepatic | 19q13.42 |
| rs8115257 | 20 | 25207931 | 6.66E-10 | Hepatic | 20p11.21 |
| rs738703 | 20 | 34190699 | 6.11E-09 | Hepatic | 20q11.22 |
| rs2092203 | 20 | 39858545 | 2.18E-11 | Hepatic | 20q12 |
| rs1800961 | 20 | 43042364 | 8.28E-64 | Hepatic | 20q13.12 |
| rs6065904 | 20 | 44534651 | 5.10E-12 | Hepatic | 20q13.12 |
| rs6018652 | 20 | 46340974 | 1.19E-09 | Hepatic | 20q13.12 |
| rs1151624 | 20 | 62369895 | 2.02E-11 | Hepatic | 20q13.33 |
| rs2298428 | 22 | 21982892 | 7.06E-10 | Hepatic | 22q11.21 |
| rs3747207 | 22 | 44324855 | 9.01E-25 | Hepatic | 22q13.31 |
| rs11591147 | 1 | 55505647 | 2.14E-39 | Immune | 1p32.3 |
| rs7531579 | 1 | 63147040 | 9.02E-48 | Immune | 1p31.3 |
| rs12740374 | 1 | 109817590 | 3.49E-51 | Immune | 1p13.3 |
| rs2811290 | 1 | 178572978 | 1.54E-10 | Immune | 1q25.2 |
| rs2642438 | 1 | 220970028 | 1.18E-34 | Immune | 1q41 |
| rs4846916 | 1 | 230296492 | 2.09E-18 | Immune | 1q42.13 |
| rs558971 | 1 | 234853406 | 1.06E-17 | Immune | 1q42.3 |
| rs907868 | 2 | 20371167 | 1.66E-19 | Immune | 2p24.1 |
| rs312970 | 2 | 21369815 | 3.66E-120 | Immune | 2p24.1 |
| rs1260326 | 2 | 27730940 | 2.49E-76 | Immune | 2p23.3 |
| rs75331444 | 2 | 44069772 | 2.38E-09 | Immune | 2p21 |
| rs2422283 | 2 | 64901725 | 5.63E-10 | Immune | 2p14 |
| rs1145106 | 3 | 136111917 | 1.02E-09 | Immune | 3q22.3 |
| rs12486792 | 3 | 170677123 | 1.10E-15 | Immune | 3q26.2 |
| rs9884390 | 4 | 69373407 | 4.47E-31 | Immune | 4q13.2 |
| rs1458038 | 4 | 81164723 | 4.82E-09 | Immune | 4q21.21 |
| 4:100486558_CTAT_C | 4 | 100486558 | 5.39E-12 | Immune | 4q23 |
| rs12916 | 5 | 74656539 | 3.42E-14 | Immune | 5q13.3 |
| rs2270924 | 5 | 156482101 | 1.22E-11 | Immune | 5q33.3 |
| rs3800461 | 6 | 34616322 | 5.82E-09 | Immune | 6p21.31 |
| rs9367490 | 6 | 52459940 | 6.13E-10 | Immune | 6p12.2 |
| rs55994889 | 6 | 127187921 | 1.19E-09 | Immune | 6q22.33 |
| rs118039278 | 6 | 160985526 | 7.16E-40 | Immune | 6q25.3 |
| rs10233430 | 7 | 1051664 | 4.36E-50 | Immune | 7p22.3 |
| rs17685 | 7 | 75616105 | 9.84E-09 | Immune | 7q11.23 |
| rs11461696 | 8 | 6571241 | 6.54E-09 | Immune | 8p23.1 |
| rs55686478 | 8 | 18250375 | 3.79E-09 | Immune | 8p22 |
| rs765547 | 8 | 19866274 | 2.83E-09 | Immune | 8p21.3 |
| rs2081687 | 8 | 59388565 | 2.79E-15 | Immune | 8q12.1 |
| rs2737252 | 8 | 116663898 | 8.48E-11 | Immune | 8q23.3 |
| rs2223056 | 8 | 117010468 | 3.64E-09 | Immune | 8q23.3 |
| rs2954021 | 8 | 126482077 | 4.60E-96 | Immune | 8q24.13 |
| rs686030 | 9 | 15304782 | 1.95E-19 | Immune | 9p22.3 |
| rs2740488 | 9 | 107661742 | 3.21E-32 | Immune | 9q31.1 |
| 9:136151579_TGGTGCAGGCGCAGGAAAAAATTGTGGCAATTCCTCA_T | 9 | 136151579 | 9.93E-34 | Immune | 9q34.2 |
| rs79618354 | 10 | 5320950 | 1.55E-14 | Immune | 10p15.1 |
| rs10881987 | 10 | 93641869 | 3.43E-10 | Immune | 10q23.32 |
| rs11190478 | 10 | 102102132 | 1.57E-12 | Immune | 10q24.31 |
| rs10787429 | 10 | 113949664 | 1.76E-23 | Immune | 10q25.2 |
| rs99780 | 11 | 61596633 | 6.59E-278 | Immune | 11q12.2 |
| rs499974 | 11 | 75455021 | 2.07E-09 | Immune | 11q13.5 |
| rs12225230 | 11 | 116728630 | 2.10E-32 | Immune | 11q23.3 |
| 12:109888158_GGCC_G | 12 | 109888158 | 2.92E-11 | Immune | 12q24.11 |
| rs7970695 | 12 | 121423376 | 8.59E-42 | Immune | 12q24.31 |
| rs10773112 | 12 | 125338529 | 1.31E-12 | Immune | 12q24.31 |
| rs11147492 | 13 | 32960363 | 1.35E-10 | Immune | 13q13.1 |
| 14:75228486_CT_C | 14 | 75228486 | 3.52E-09 | Immune | 14q24.3 |
| rs139974673 | 15 | 44027885 | 1.17E-12 | Immune | 15q15.3 |
| rs55966152 | 15 | 58561006 | 5e-324 | Immune | 15q21.3 |
| rs780583791 | 15 | 75382239 | 9.59E-10 | Immune | 15q24.2 |
| rs11075253 | 16 | 15148646 | 1.63E-27 | Immune | 16p13.11 |
| rs3816117 | 16 | 56996158 | 4.04E-17 | Immune | 16q13 |
| rs11648003 | 16 | 72052348 | 1.64E-17 | Immune | 16q22.2 |
| rs79843305 | 17 | 40395211 | 4.72E-10 | Immune | 17q21.2 |
| rs72836561 | 17 | 41926126 | 1.20E-26 | Immune | 17q21.31 |
| rs4399567 | 17 | 45749172 | 6.13E-18 | Immune | 17q21.32 |
| rs1801689 | 17 | 64210580 | 5.54E-25 | Immune | 17q24.2 |
| rs77542162 | 17 | 67081278 | 2.36E-16 | Immune | 17q24.2 |
| rs77960347 | 18 | 47109955 | 2.89E-60 | Immune | 18q21.1 |
| rs1736194 | 19 | 2786397 | 1.15E-09 | Immune | 19p13.3 |
| rs73015021 | 19 | 11192915 | 1.55E-72 | Immune | 19p13.2 |
| rs739846 | 19 | 19419071 | 7.11E-16 | Immune | 19p13.11 |
| rs7412 | 19 | 45412079 | 2.26E-205 | Immune | 19q13.32 |
| rs8112972 | 19 | 46390605 | 4.40E-11 | Immune | 19q13.32 |
| 19:49213853_AT_A | 19 | 49213853 | 1.44E-22 | Immune | 19q13.33 |
| rs771193934 | 20 | 17844518 | 9.24E-09 | Immune | 20p12.1 |
| rs6087723 | 20 | 34175783 | 3.25E-12 | Immune | 20q11.22 |
| rs1883711 | 20 | 39179822 | 1.31E-15 | Immune | 20q12 |
| rs2866372 | 20 | 39841091 | 1.05E-09 | Immune | 20q12 |
| rs1800961 | 20 | 43042364 | 3.75E-18 | Immune | 20q13.12 |
| rs7528419 | 1 | 109817192 | 3.65E-11 | Metabolic | 1p13.3 |
| rs5082 | 1 | 161193683 | 2.33E-30 | Metabolic | 1q23.3 |
| rs533617 | 2 | 21233972 | 2.10E-18 | Metabolic | 2p24.1 |
| rs1260326 | 2 | 27730940 | 6.56E-11 | Metabolic | 2p23.3 |
| rs4610076 | 2 | 102115331 | 1.22E-09 | Metabolic | 2q11.2 |
| rs6745833 | 2 | 174798495 | 3.88E-10 | Metabolic | 2q31.1 |
| rs9973507 | 2 | 191725648 | 1.71E-15 | Metabolic | 2q32.2 |
| rs1047891 | 2 | 211540507 | 1.27E-19 | Metabolic | 2q34 |
| rs13322435 | 3 | 156795468 | 5.39E-11 | Metabolic | 3q25.31 |
| rs6791056 | 3 | 160025238 | 1.83E-10 | Metabolic | 3q25.33 |
| rs4917 | 3 | 186337713 | 2.67E-19 | Metabolic | 3q27.3 |
| rs6847390 | 4 | 26177356 | 7.73E-11 | Metabolic | 4p15.2 |
| rs1440580 | 4 | 89226747 | 9.75E-21 | Metabolic | 4q22.1 |
| 4:100039422_GAAACA_G | 4 | 100039422 | 4.30E-10 | Metabolic | 4q23 |
| rs752736590 | 4 | 155490649 | 2.08E-22 | Metabolic | 4q31.3 |
| rs7718024 | 5 | 131785879 | 1.69E-15 | Metabolic | 5q31.1 |
| rs9395819 | 6 | 52643987 | 1.40E-11 | Metabolic | 6p12.2 |
| 6:111558032_CT_C | 6 | 111558032 | 2.85E-39 | Metabolic | 6q21 |
| rs9402686 | 6 | 135427817 | 6.34E-14 | Metabolic | 6q23.3 |
| rs140570886 | 6 | 161013013 | 2.08E-14 | Metabolic | 6q26 |
| rs74453229 | 7 | 2481061 | 4.58E-09 | Metabolic | 7p22.3 |
| rs4544985 | 7 | 17983807 | 2.59E-11 | Metabolic | 7p21.1 |
| rs35555151 | 8 | 9219463 | 3.48E-10 | Metabolic | 8p23.1 |
| rs12679834 | 8 | 19820433 | 1.74E-27 | Metabolic | 8p21.3 |
| rs2980888 | 8 | 126507308 | 1.78E-17 | Metabolic | 8q24.13 |
| rs686030 | 9 | 15304782 | 3.27E-35 | Metabolic | 9p22.3 |
| rs2740488 | 9 | 107661742 | 2.80E-79 | Metabolic | 9q31.1 |
| rs10909680 | 10 | 17889190 | 3.70E-09 | Metabolic | 10p12.33 |
| rs2393966 | 10 | 65134814 | 1.48E-25 | Metabolic | 10q21.3 |
| rs17096421 | 10 | 88820592 | 2.09E-10 | Metabolic | 10q23.2 |
| rs7078003 | 10 | 99359412 | 1.24E-09 | Metabolic | 10q24.2 |
| rs6537612 | 10 | 135336717 | 9.26E-09 | Metabolic | 10q26.3 |
| rs174595 | 11 | 61619893 | 6.97e-322 | Metabolic | 11q12.2 |
| rs2229738 | 11 | 68562328 | 4.88E-25 | Metabolic | 11q13.3 |
| rs3741298 | 11 | 116657561 | 9.36E-24 | Metabolic | 11q23.3 |
| rs79295634 | 12 | 47180008 | 1.91E-16 | Metabolic | 12q13.11 |
| rs2638315 | 12 | 56865056 | 1.35E-64 | Metabolic | 12q13.3 |
| rs2393791 | 12 | 121423956 | 3.25E-16 | Metabolic | 12q24.31 |
| rs11839648 | 13 | 91996303 | 6.91E-09 | Metabolic | 13q31.3 |
| rs1303 | 14 | 94844843 | 3.16E-14 | Metabolic | 14q32.13 |
| rs12883091 | 14 | 102669022 | 1.49E-11 | Metabolic | 14q32.31 |
| rs8009444 | 14 | 106194461 | 2.67E-26 | Metabolic | 14q32.33 |
| rs150844304 | 15 | 43726625 | 3.72E-22 | Metabolic | 15q15.3 |
| rs261334 | 15 | 58726744 | 1.31E-11 | Metabolic | 15q21.3 |
| rs340029 | 15 | 60894965 | 1.40E-11 | Metabolic | 15q22.2 |
| rs10444863 | 15 | 78358837 | 1.83E-11 | Metabolic | 15q25.1 |
| rs12928099 | 16 | 15150505 | 1.97E-49 | Metabolic | 16p13.11 |
| rs247617 | 16 | 56990716 | 5.44E-169 | Metabolic | 16q13 |
| rs76495028 | 16 | 68177036 | 8.55E-11 | Metabolic | 16q22.1 |
| rs10530053 | 16 | 69686676 | 8.15E-13 | Metabolic | 16q22.1 |
| rs10530053 | 16 | 69686676 | 8.15E-13 | Metabolic | 16q22.1 |
| rs12598251 | 16 | 70658252 | 1.13E-09 | Metabolic | 16q22.1 |
| rs763665 | 16 | 72078043 | 3.77E-50 | Metabolic | 16q22.2 |
| rs57652769 | 16 | 79753976 | 3.22E-18 | Metabolic | 16q23.2 |
| rs7196553 | 16 | 88541097 | 2.27E-09 | Metabolic | 16q24.2 |
| rs72836561 | 17 | 41926126 | 2.82E-23 | Metabolic | 17q21.31 |
| rs1292045 | 17 | 57947212 | 2.80E-11 | Metabolic | 17q23.1 |
| rs77542162 | 17 | 67081278 | 6.35E-10 | Metabolic | 17q24.2 |
| rs28534973 | 19 | 18535607 | 4.17E-10 | Metabolic | 19p13.11 |
| rs58895965 | 19 | 35551428 | 6.30E-10 | Metabolic | 19q13.12 |
| rs1065853 | 19 | 45413233 | 2.05E-35 | Metabolic | 19q13.32 |
| rs67546213 | 19 | 50082587 | 8.70E-39 | Metabolic | 19q13.33 |
| rs1800961 | 20 | 43042364 | 1.47E-14 | Metabolic | 20q13.12 |

**eTable 5: The detailed statistics of the three key genetic parameters of SbayesS**

| **BAG** | **h2_mean** | **h2_se** | **S** | **S_se** | **Pi** | **Pi_se** |
| --- | --- | --- | --- | --- | --- | --- |
| Endocrine | 0.147486 | 0.002398 | -0.928022 | 0.063385 | 0.003089 | 0.000185 |
| Digestive | 0.125212 | 0.00223 | -0.871973 | 0.077194 | 0.001922 | 0.000142 |
| Hepatic | 0.177534 | 0.002119 | -0.94177 | 0.096908 | 0.001651 | 0.000109 |
| Immune | 0.153633 | 0.001836 | -0.769576 | 0.093281 | 0.001194 | 8.90E-05 |
| Metabolic | 0.0916 | 0.002549 | -0.832472 | 0.077095 | 0.002039 | 0.000195 |

**eTable 6: The genetic correlation results**

A) Genetic and Phenotypic correlations among the 5 MetBAGs

Phenotypic correlation:

|  | Endocrine | Digestive | Hepatic | Immune | Metabolic |
| --- | --- | --- | --- | --- | --- |
| Endocrine | 1 | 0.34216727 | 0.51794626 | 0.49714094 | 0.51712057 |
| Digestive | 0.34216727 | 1 | 0.47661427 | 0.64996356 | 0.29317315 |
| Hepatic | 0.51794626 | 0.47661427 | 1 | 0.66915195 | 0.22778409 |
| Immune | 0.49714094 | 0.64996356 | 0.66915195 | 1 | 0.14387914 |
| Metabolic | 0.51712057 | 0.29317315 | 0.22778409 | 0.14387914 | 1 |

Genetic correlation:

|  | Endocrine | Digestive | Hepatic | Immune | Metabolic |
| --- | --- | --- | --- | --- | --- |
| Endocrine | 1 | 0.2142 | 0.6613 | 0.6073 | 0.4031 |
| Digestive | 0.2142 | 1 | 0.3791 | 0.5618 | 0.5613 |
| Hepatic | 0.6613 | 0.3791 | 1 | 0.7797 | 0.198 |
| Immune | 0.6073 | 0.5618 | 0.7797 | 1 | 0.1733 |
| Metabolic | 0.4031 | 0.5613 | 0.198 | 0.1733 | 1 |

B) Genetic correlations between the 5 MetBAGs and 327 metabolites

Due to the large-sized table, the results are presented in **Supplementary eFile 1**.

C) Genetic correlation between the 5 MetBAGs and 545 traits

Due to the large-sized table, the results are presented in **Supplementary eFile 2**.

**eTable 7: The Mendelian randomization results**

Due to the large table, the results are presented in **Supplementary eFile 3**. We present only the results that pass the Bonferroni correction for each network based on the number of DEs.

**eTable 8: The classification results to predict the 14 systemic disease categories**

A) **Training/test dataset**

| **Disease_category** | **Feature_combination** | **accuracy** | **balanced_accuracy** | **sensitivity** | **specificity** | **ppv** | **npv** | **train_N** |
| --- | --- | --- | --- | --- | --- | --- | --- | --- |
| infectious_parasitic_disease_diagnosis | bagprs | 0.52024933 | 0.50781826 | 0.7963306 | 0.21930593 | 0.52648968 | 0.49693569 | 54866 |
| neoplasms_diagnosis | bagprs | 0.70884138 | 0.5000233 | 0.99929995 | 0.00074664 | 0.70913024 | 0.30434783 | 96693 |
| blood_and_immune_system_diagnosis | bagprs | 0.67521098 | 0.5 | 1 | 0 | 0.67521098 | 0 | 85554 |
| endocrine_nutritional_metabolic_disease_diagnosis | bagprs | 0.71119434 | 0.5 | 1 | 0 | 0.71119434 | 0 | 97460 |
| mental_behavioural_disorder_diagnosis | bagprs | 0.47528144 | 0.50269065 | 0.19290255 | 0.81247876 | 0.55124729 | 0.45741016 | 58094 |
| nerve_system_diagnosis | bagprs | 0.54701716 | 0.50140773 | 0.92424242 | 0.07857304 | 0.55468665 | 0.45510026 | 59574 |
| eye_diagnosis | bagprs | 0.55418644 | 0.50360725 | 0.79094104 | 0.21627345 | 0.59023272 | 0.42022813 | 65330 |
| ear_diagnosis | bagprs | 0.7281729 | 0.5 | 0 | 1 | 0 | 0.7281729 | 32620 |
| circular_system_diagnosis | bagprs | 0.77908741 | 0.5 | 1 | 0 | 0.77908741 | 0 | 130486 |
| respiratory_system_diagnosis | bagprs | 0.65504365 | 0.5002023 | 0.99826607 | 0.00213853 | 0.65553484 | 0.39333333 | 80071 |
| digestive_system_diagnosis | bagprs | 0.80086527 | 0.5 | 1 | 0 | 0.80086527 | 0 | 145851 |
| skin_system_diagnosis | bagprs | 0.51126502 | 0.5046216 | 0.57285076 | 0.43639243 | 0.55270954 | 0.45662209 | 58766 |
| musculoskeletal_system_diagnosis | bagprs | 0.75714201 | 0.5 | 1 | 0 | 0.75714201 | 0 | 117789 |
| genitourinary_system_diagnosis | bagprs | 0.72611385 | 0.5 | 1 | 0 | 0.72611385 | 0 | 103313 |
| infectious_parasitic_disease_diagnosis | bag | 0.69436445 | 0.68829254 | 0.82921545 | 0.54736962 | 0.66632968 | 0.74620897 | 54866 |
| neoplasms_diagnosis | bag | 0.77800875 | 0.67235893 | 0.9249639 | 0.41975396 | 0.79533997 | 0.69647808 | 96693 |
| blood_and_immune_system_diagnosis | bag | 0.75766183 | 0.68347487 | 0.89518237 | 0.47176737 | 0.77891249 | 0.684043 | 85554 |
| endocrine_nutritional_metabolic_disease_diagnosis | bag | 0.77505643 | 0.65833384 | 0.93467315 | 0.38199453 | 0.78833049 | 0.70366492 | 97460 |
| mental_behavioural_disorder_diagnosis | bag | 0.70542569 | 0.69000011 | 0.86434513 | 0.5156551 | 0.68061367 | 0.76095196 | 58094 |
| nerve_system_diagnosis | bag | 0.70762413 | 0.69187168 | 0.83790909 | 0.54583427 | 0.69614804 | 0.73058326 | 59574 |
| eye_diagnosis | bag | 0.71772539 | 0.68458823 | 0.87283613 | 0.49634033 | 0.71210126 | 0.73224074 | 65330 |
| ear_diagnosis | bag | 0.74258124 | 0.5651635 | 0.17638435 | 0.95394266 | 0.58841234 | 0.75625793 | 32620 |
| circular_system_diagnosis | bag | 0.81819506 | 0.62070002 | 0.97452292 | 0.26687712 | 0.82418908 | 0.74812798 | 130486 |
| respiratory_system_diagnosis | bag | 0.74531353 | 0.65713094 | 0.9407797 | 0.37348218 | 0.74069518 | 0.76826722 | 80071 |
| digestive_system_diagnosis | bag | 0.83308308 | 0.6030387 | 0.98534334 | 0.22073406 | 0.83566885 | 0.78924043 | 145851 |
| skin_system_diagnosis | bag | 0.70801144 | 0.69389712 | 0.83885374 | 0.5489405 | 0.6933429 | 0.73697798 | 58766 |
| musculoskeletal_system_diagnosis | bag | 0.80343665 | 0.62531373 | 0.971665 | 0.27896246 | 0.80774035 | 0.75949367 | 117789 |
| genitourinary_system_diagnosis | bag | 0.78805184 | 0.65824881 | 0.94527907 | 0.37121855 | 0.7994228 | 0.71900883 | 103313 |
| infectious_parasitic_disease_diagnosis | bag_bagprs | 0.70165494 | 0.69598053 | 0.82767779 | 0.56428327 | 0.67433517 | 0.75025324 | 54866 |
| neoplasms_diagnosis | bag_bagprs | 0.77663326 | 0.64976899 | 0.95309697 | 0.34644102 | 0.78046887 | 0.75185185 | 96693 |
| blood_and_immune_system_diagnosis | bag_bagprs | 0.75913458 | 0.66375914 | 0.93593228 | 0.39158599 | 0.76179338 | 0.74619394 | 85554 |
| endocrine_nutritional_metabolic_disease_diagnosis | bag_bagprs | 0.77768315 | 0.66242773 | 0.93529352 | 0.38956194 | 0.79048896 | 0.70970874 | 97460 |
| mental_behavioural_disorder_diagnosis | bag_bagprs | 0.71081351 | 0.69703147 | 0.85280071 | 0.54126223 | 0.6894321 | 0.7548591 | 58094 |
| nerve_system_diagnosis | bag_bagprs | 0.71255917 | 0.69442102 | 0.86257576 | 0.52626628 | 0.69335509 | 0.75512959 | 59574 |
| eye_diagnosis | bag_bagprs | 0.72211848 | 0.7008032 | 0.82189249 | 0.57971391 | 0.73622479 | 0.69516596 | 65330 |
| ear_diagnosis | bag_bagprs | 0.7281729 | 0.5 | 0 | 1 | 0 | 0.7281729 | 32620 |
| circular_system_diagnosis | bag_bagprs | 0.81931395 | 0.62930931 | 0.96971277 | 0.28890585 | 0.82786218 | 0.73007802 | 130486 |
| respiratory_system_diagnosis | bag_bagprs | 0.74977208 | 0.66729723 | 0.93258641 | 0.40200805 | 0.7478989 | 0.75815162 | 80071 |
| digestive_system_diagnosis | bag_bagprs | 0.83116331 | 0.59909799 | 0.98476119 | 0.21343479 | 0.83430283 | 0.7769144 | 145851 |
| skin_system_diagnosis | bag_bagprs | 0.71544771 | 0.70484543 | 0.81373279 | 0.59595807 | 0.71001786 | 0.72464698 | 58766 |
| musculoskeletal_system_diagnosis | bag_bagprs | 0.80555909 | 0.62942225 | 0.97191169 | 0.28693281 | 0.80949979 | 0.76617194 | 117789 |
| genitourinary_system_diagnosis | bag_bagprs | 0.78379294 | 0.64006295 | 0.95788955 | 0.32223636 | 0.78933609 | 0.74268958 | 103313 |
| infectious_parasitic_disease_diagnosis | bag_bagprs_cov | 0.74233587 | 0.73956792 | 0.80380919 | 0.67532665 | 0.72963456 | 0.75948933 | 54866 |
| neoplasms_diagnosis | bag_bagprs_cov | 0.80254 | 0.70625365 | 0.9364709 | 0.47603641 | 0.81333283 | 0.7545224 | 96693 |
| blood_and_immune_system_diagnosis | bag_bagprs_cov | 0.78260514 | 0.73163292 | 0.87709246 | 0.58617339 | 0.81502751 | 0.69642552 | 85554 |
| endocrine_nutritional_metabolic_disease_diagnosis | bag_bagprs_cov | 0.79648061 | 0.69958162 | 0.92898879 | 0.47017444 | 0.81195148 | 0.72890505 | 97460 |
| mental_behavioural_disorder_diagnosis | bag_bagprs_cov | 0.73807966 | 0.73148481 | 0.80602208 | 0.65694754 | 0.73723494 | 0.73932078 | 58094 |
| nerve_system_diagnosis | bag_bagprs_cov | 0.74060832 | 0.7287743 | 0.83848485 | 0.61906375 | 0.73214616 | 0.75529131 | 59574 |
| eye_diagnosis | bag_bagprs_cov | 0.78556559 | 0.76881789 | 0.86395939 | 0.67367639 | 0.79074145 | 0.77626509 | 65330 |
| ear_diagnosis | bag_bagprs_cov | 0.7969344 | 0.71214166 | 0.5263336 | 0.89794973 | 0.65815823 | 0.83548122 | 32620 |
| circular_system_diagnosis | bag_bagprs_cov | 0.83687139 | 0.68601073 | 0.95628566 | 0.41573579 | 0.85233828 | 0.72948624 | 130486 |
| respiratory_system_diagnosis | bag_bagprs_cov | 0.77147781 | 0.72811655 | 0.8675927 | 0.5886404 | 0.8004817 | 0.70033205 | 80071 |
| digestive_system_diagnosis | bag_bagprs_cov | 0.83908235 | 0.61554098 | 0.98703845 | 0.24404352 | 0.84002798 | 0.82399442 | 145851 |
| skin_system_diagnosis | bag_bagprs_cov | 0.74619678 | 0.73500058 | 0.84998759 | 0.62001357 | 0.73114579 | 0.77270805 | 58766 |
| musculoskeletal_system_diagnosis | bag_bagprs_cov | 0.81651937 | 0.64588503 | 0.97767512 | 0.31409495 | 0.81630513 | 0.81860423 | 117789 |
| genitourinary_system_diagnosis | bag_bagprs_cov | 0.81021749 | 0.71332878 | 0.92757642 | 0.49908114 | 0.83077437 | 0.72216824 | 103313 |

B) **Independent test dataset**

| **Disease_category** | **Feature_combination** | **accuracy** | **balanced_accuracy** | **sensitivity** | **specificity** | **ppv** | **npv** | **train_N** |
| --- | --- | --- | --- | --- | --- | --- | --- | --- |
| infectious_parasitic_disease_diagnosis | bagprs | 0.5135 | 0.50061459 | 0.79888804 | 0.20234114 | 0.52198422 | 0.47992067 | 10000 |
| neoplasms_diagnosis | bagprs | 0.7084 | 0.500216 | 0.99802567 | 0.00240633 | 0.7091893 | 0.33333333 | 10000 |
| blood_and_immune_system_diagnosis | bagprs | 0.6752 | 0.5 | 1 | 0 | 0.6752 | 0 | 10000 |
| endocrine_nutritional_metabolic_disease_diagnosis | bagprs | 0.7112 | 0.5 | 1 | 0 | 0.7112 | 0 | 10000 |
| mental_behavioural_disorder_diagnosis | bagprs | 0.4643 | 0.49423136 | 0.15564131 | 0.83282141 | 0.52641392 | 0.45238946 | 10000 |
| nerve_system_diagnosis | bagprs | 0.5435 | 0.50435452 | 0.86748511 | 0.14122394 | 0.55639185 | 0.46187683 | 10000 |
| eye_diagnosis | bagprs | 0.5485 | 0.50422611 | 0.75578231 | 0.2526699 | 0.59072179 | 0.42026645 | 10000 |
| ear_diagnosis | bagprs | 0.7282 | 0.5 | 0 | 1 | 0 | 0.7282 | 10000 |
| circular_system_diagnosis | bagprs | 0.7791 | 0.5 | 1 | 0 | 0.7791 | 0 | 10000 |
| respiratory_system_diagnosis | bagprs | 0.6536 | 0.49924605 | 0.99588038 | 0.00261172 | 0.65505821 | 0.25 | 10000 |
| digestive_system_diagnosis | bagprs | 0.8009 | 0.5 | 1 | 0 | 0.8009 | 0 | 10000 |
| skin_system_diagnosis | bagprs | 0.5156 | 0.5115506 | 0.55312557 | 0.46997563 | 0.55924083 | 0.46380932 | 10000 |
| musculoskeletal_system_diagnosis | bagprs | 0.7571 | 0.5 | 1 | 0 | 0.7571 | 0 | 10000 |
| genitourinary_system_diagnosis | bagprs | 0.7261 | 0.5 | 1 | 0 | 0.7261 | 0 | 10000 |
| infectious_parasitic_disease_diagnosis | bag | 0.4688 | 0.48939525 | 0.01265337 | 0.96613712 | 0.28947368 | 0.47298404 | 10000 |
| neoplasms_diagnosis | bag | 0.5629 | 0.62103723 | 0.48201946 | 0.760055 | 0.83041788 | 0.37576479 | 10000 |
| blood_and_immune_system_diagnosis | bag | 0.598 | 0.63592287 | 0.5276955 | 0.74415025 | 0.81087847 | 0.4311452 | 10000 |
| endocrine_nutritional_metabolic_disease_diagnosis | bag | 0.322 | 0.49753139 | 0.08197413 | 0.91308864 | 0.69904077 | 0.28769365 | 10000 |
| mental_behavioural_disorder_diagnosis | bag | 0.4765 | 0.51370861 | 0.09279677 | 0.93462045 | 0.62889166 | 0.46319452 | 10000 |
| nerve_system_diagnosis | bag | 0.6634 | 0.66238726 | 0.67178191 | 0.6529926 | 0.70620611 | 0.61572606 | 10000 |
| eye_diagnosis | bag | 0.5909 | 0.60857192 | 0.50816327 | 0.70898058 | 0.71363745 | 0.50249441 | 10000 |
| ear_diagnosis | bag | 0.7226 | 0.49696198 | 0.00257542 | 0.99134853 | 0.1 | 0.72698892 | 10000 |
| circular_system_diagnosis | bag | 0.5959 | 0.57022148 | 0.61622385 | 0.5242191 | 0.82040328 | 0.27917068 | 10000 |
| respiratory_system_diagnosis | bag | 0.5984 | 0.59543538 | 0.60497406 | 0.58589669 | 0.73534866 | 0.43815104 | 10000 |
| digestive_system_diagnosis | bag | 0.7872 | 0.65089889 | 0.87738794 | 0.42440984 | 0.85978221 | 0.46250684 | 10000 |
| skin_system_diagnosis | bag | 0.5913 | 0.60181106 | 0.49389466 | 0.70972745 | 0.67412935 | 0.53561873 | 10000 |
| musculoskeletal_system_diagnosis | bag | 0.5369 | 0.58133966 | 0.49491481 | 0.66776451 | 0.82279315 | 0.29783327 | 10000 |
| genitourinary_system_diagnosis | bag | 0.6557 | 0.64785986 | 0.66519763 | 0.63052209 | 0.82677165 | 0.41534392 | 10000 |
| infectious_parasitic_disease_diagnosis | bag_bagprs | 0.4681 | 0.4883174 | 0.02032209 | 0.95631271 | 0.33650794 | 0.47237997 | 10000 |
| neoplasms_diagnosis | bag_bagprs | 0.6032 | 0.62928121 | 0.56691581 | 0.69164661 | 0.81757169 | 0.39582923 | 10000 |
| blood_and_immune_system_diagnosis | bag_bagprs | 0.6372 | 0.64633723 | 0.62026066 | 0.67241379 | 0.79741051 | 0.45998315 | 10000 |
| endocrine_nutritional_metabolic_disease_diagnosis | bag_bagprs | 0.3166 | 0.49795087 | 0.06861642 | 0.92728532 | 0.6991404 | 0.28789508 | 10000 |
| mental_behavioural_disorder_diagnosis | bag_bagprs | 0.4645 | 0.50425134 | 0.05457552 | 0.95392716 | 0.58579882 | 0.4580217 | 10000 |
| nerve_system_diagnosis | bag_bagprs | 0.6531 | 0.65298048 | 0.65408919 | 0.65187178 | 0.69996136 | 0.60281924 | 10000 |
| eye_diagnosis | bag_bagprs | 0.5953 | 0.61369378 | 0.50918367 | 0.71820388 | 0.72057762 | 0.50624465 | 10000 |
| ear_diagnosis | bag_bagprs | 0.7282 | 0.5 | 0 | 1 | 0 | 0.7282 | 10000 |
| circular_system_diagnosis | bag_bagprs | 0.6551 | 0.58778059 | 0.70838147 | 0.46717972 | 0.82422342 | 0.31234867 | 10000 |
| respiratory_system_diagnosis | bag_bagprs | 0.5896 | 0.5938136 | 0.58025633 | 0.60737086 | 0.73758728 | 0.43208092 | 10000 |
| digestive_system_diagnosis | bag_bagprs | 0.7792 | 0.65062202 | 0.86427769 | 0.43696635 | 0.86062415 | 0.444558 | 10000 |
| skin_system_diagnosis | bag_bagprs | 0.5882 | 0.59930086 | 0.48532896 | 0.71327277 | 0.67298458 | 0.53268244 | 10000 |
| musculoskeletal_system_diagnosis | bag_bagprs | 0.5262 | 0.5767897 | 0.47840444 | 0.67517497 | 0.821129 | 0.29343353 | 10000 |
| genitourinary_system_diagnosis | bag_bagprs | 0.6941 | 0.66566226 | 0.72854979 | 0.60277474 | 0.82941361 | 0.45582551 | 10000 |
| infectious_parasitic_disease_diagnosis | bag_bagprs_cov | 0.4941 | 0.51324071 | 0.07016871 | 0.95631271 | 0.63652174 | 0.48541114 | 10000 |
| neoplasms_diagnosis | bag_bagprs_cov | 0.6449 | 0.68483774 | 0.5893386 | 0.78033689 | 0.86737235 | 0.43805481 | 10000 |
| blood_and_immune_system_diagnosis | bag_bagprs_cov | 0.6017 | 0.65887466 | 0.49570498 | 0.82204433 | 0.85273885 | 0.43950617 | 10000 |
| endocrine_nutritional_metabolic_disease_diagnosis | bag_bagprs_cov | 0.5022 | 0.60570991 | 0.36065804 | 0.85076177 | 0.85614152 | 0.35079954 | 10000 |
| mental_behavioural_disorder_diagnosis | bag_bagprs_cov | 0.4996 | 0.53655397 | 0.1185226 | 0.95458534 | 0.75704225 | 0.47562309 | 10000 |
| nerve_system_diagnosis | bag_bagprs_cov | 0.705 | 0.70133522 | 0.73533129 | 0.66733916 | 0.73294943 | 0.67004276 | 10000 |
| eye_diagnosis | bag_bagprs_cov | 0.7158 | 0.72360643 | 0.6792517 | 0.76796117 | 0.80686869 | 0.62653465 | 10000 |
| ear_diagnosis | bag_bagprs_cov | 0.765 | 0.61762015 | 0.29470199 | 0.94053831 | 0.64910859 | 0.78131417 | 10000 |
| circular_system_diagnosis | bag_bagprs_cov | 0.5074 | 0.60586216 | 0.4294699 | 0.78225441 | 0.87431408 | 0.27992872 | 10000 |
| respiratory_system_diagnosis | bag_bagprs_cov | 0.6007 | 0.62842817 | 0.53921269 | 0.71764364 | 0.7841136 | 0.45020936 | 10000 |
| digestive_system_diagnosis | bag_bagprs_cov | 0.8072 | 0.67432946 | 0.89511799 | 0.45354093 | 0.86823301 | 0.51807229 | 10000 |
| skin_system_diagnosis | bag_bagprs_cov | 0.6711 | 0.6792287 | 0.59577182 | 0.76268558 | 0.75322581 | 0.60812721 | 10000 |
| musculoskeletal_system_diagnosis | bag_bagprs_cov | 0.6412 | 0.64882287 | 0.63399815 | 0.66364759 | 0.85454869 | 0.36778462 | 10000 |
| genitourinary_system_diagnosis | bag_bagprs_cov | 0.6686 | 0.67822984 | 0.65693431 | 0.69952537 | 0.85285178 | 0.43476288 | 10000 |

**eTable 9: The survival analysis to predict mortality**

| **BAG** | **hazard_ratio** | **CI_lower_bound** | **CI_upper_bound** | **p_value** | **n_case** | **n_noncase** |
| --- | --- | --- | --- | --- | --- | --- |
| Endocrine | 1.10863922 | 1.09409609 | 1.12337567 | 6.77E-53 | 22672 | 236339 |
| Digestive | 1.0737658 | 1.05925262 | 1.08847782 | 1.18E-24 | 23458 | 242535 |
| Hepatic | 0.92964539 | 0.91664418 | 0.94283101 | 3.24E-24 | 23371 | 241206 |
| Immune | 1.02070991 | 1.00690334 | 1.03470579 | 0.00317718 | 23893 | 245587 |
| Metabolic | 1.1697998 | 1.15424749 | 1.18556167 | 1.00E-116 | 21650 | 226170 |
| Endocrine_PRS | 0.99928418 | 0.98681731 | 1.01190855 | 0.91098781 | 24152 | 248023 |
| Digestive_PRS | 1.01757121 | 1.00487361 | 1.03042925 | 0.00655142 | 24152 | 248023 |
| Hepatic_PRS | 0.99375711 | 0.98136958 | 1.00630101 | 0.32781987 | 24152 | 248023 |
| Immune_PRS | 1.00196212 | 0.98942741 | 1.01465563 | 0.7602299 | 24152 | 248023 |
| Metabolic_PRS | 1.01898682 | 1.00624122 | 1.03189386 | 0.00340289 | 24152 | 248023 |

**eTable 10: The survival analysis to predict ICD-based disease incidence**

| **MetBAG** | **DE** | **hazard_ratio** | **CI_lower_bound** | **CI_upper_bound** | **p_value** | **N_case** | **N_noncase** |
| --- | --- | --- | --- | --- | --- | --- | --- |
| Endocrine_MetBAG | I10 | 1.10402207 | 1.07245081 | 1.13652274 | 2.31E-11 | 4259 | 4019 |
| Endocrine_MetBAG | Z864 | 1.0917383 | 1.04894659 | 1.13627569 | 1.69E-05 | 2331 | 4019 |
| Endocrine_MetBAG | E780 | 1.23591911 | 1.18437298 | 1.28970862 | 1.94E-22 | 2027 | 4019 |
| Digestive_MetBAG | E780 | 1.10297112 | 1.05529712 | 1.15279884 | 1.38E-05 | 2027 | 4019 |
| Immune_MetBAG | E780 | 1.1014757 | 1.05559813 | 1.14934716 | 8.48E-06 | 2027 | 4019 |
| Endocrine_MetBAG | Z922 | 1.17036849 | 1.10670804 | 1.23769084 | 3.53E-08 | 1249 | 4019 |
| Endocrine_MetBAG | E119 | 1.41853524 | 1.33294986 | 1.50961585 | 3.36E-28 | 872 | 4019 |
| Digestive_MetBAG | E119 | 1.2391879 | 1.16111571 | 1.32250958 | 1.05E-10 | 872 | 4019 |
| Metabolic_MetBAG | E119 | 1.21050226 | 1.1331496 | 1.29313527 | 1.43E-08 | 872 | 4019 |
| Endocrine_MetBAG | I251 | 1.23585013 | 1.1543566 | 1.32309682 | 1.17E-09 | 851 | 4019 |
| Endocrine_MetBAG | E669 | 1.19208938 | 1.1185887 | 1.27041967 | 6.25E-08 | 875 | 4019 |
| Endocrine_MetBAG | I209 | 1.21125173 | 1.12025024 | 1.30964556 | 1.51E-06 | 652 | 4019 |
| Endocrine_MetBAG | I259 | 1.25161983 | 1.1499759 | 1.36224785 | 2.06E-07 | 557 | 4019 |
| Endocrine_MetBAG | N40 | 1.16999987 | 1.09464122 | 1.25054644 | 3.80E-06 | 1000 | 4019 |
| Endocrine_MetBAG | N179 | 1.29136378 | 1.15089936 | 1.44897153 | 1.35E-05 | 295 | 4019 |
| Endocrine_MetBAG | I252 | 1.28662296 | 1.16863692 | 1.41652091 | 2.81E-07 | 425 | 4019 |
| Endocrine_MetBAG | Z955 | 1.29680268 | 1.16339828 | 1.44550427 | 2.70E-06 | 340 | 4019 |
| Endocrine_MetBAG | C61 | 1.2775609 | 1.16362077 | 1.40265789 | 2.76E-07 | 514 | 4019 |
| Endocrine_MetBAG | E785 | 1.33021432 | 1.20679932 | 1.46625053 | 9.26E-09 | 387 | 4019 |
| Endocrine_MetBAG | G473 | 1.35070819 | 1.20263809 | 1.51700885 | 3.88E-07 | 256 | 4019 |
| Endocrine_MetBAG | Z466 | 1.26293197 | 1.12357523 | 1.41957309 | 9.11E-05 | 303 | 4019 |
| Endocrine_MetBAG | Z951 | 1.54592583 | 1.33792635 | 1.78626175 | 3.45E-09 | 186 | 4019 |
| Endocrine_MetBAG | M109 | 1.56995004 | 1.38929228 | 1.77409977 | 4.78E-13 | 218 | 4019 |
| Endocrine_MetBAG | F101 | 1.39897462 | 1.19294875 | 1.64058177 | 3.62E-05 | 136 | 4019 |
| Endocrine_MetBAG | K760 | 1.31211588 | 1.14184549 | 1.50777676 | 0.00012794 | 174 | 4019 |
| Endocrine_MetBAG | E149 | 1.51905241 | 1.26047976 | 1.83066821 | 1.13E-05 | 92 | 4019 |
| Hepatic_MetBAG | M720 | 1.38550478 | 1.22940372 | 1.56142645 | 8.98E-08 | 255 | 4019 |
| Immune_MetBAG | M720 | 1.27299829 | 1.13289809 | 1.43042404 | 4.96E-05 | 255 | 4019 |
| Digestive_MetBAG | E109 | 1.51706808 | 1.23251663 | 1.86731401 | 8.41E-05 | 61 | 4019 |
| Endocrine_MetBAG | K900 | 0.66830847 | 0.54759493 | 0.81563249 | 7.34E-05 | 112 | 4019 |
| Hepatic_MetBAG | K900 | 0.61398504 | 0.51511164 | 0.73183675 | 5.19E-08 | 112 | 4019 |
| Endocrine_MetBAG | M1099 | 1.56392493 | 1.26042766 | 1.94050104 | 4.85E-05 | 70 | 4019 |
| Digestive_MetBAG | E114 | 1.78914634 | 1.32882475 | 2.40892912 | 0.00012644 | 19 | 4019 |
| Digestive_MetBAG | G632 | 1.81507001 | 1.39937605 | 2.35424864 | 7.05E-06 | 21 | 4019 |
| Digestive_MetBAG | K703 | 1.91238922 | 1.37246781 | 2.66471278 | 0.00012787 | 9 | 4019 |

We present the detailed statistics from our survival analysis using the 5 MetBAGs to predict the ICD-based single disease entity incidence. Significant results after Bonferroni correction are presented (P-value<0.05/304). The DE was coded by ICD-10 in the UK Biobank: <https://biobank.ndph.ox.ac.uk/ukb/field.cgi?id=41270>.

**eTable 11: The incremental R2 of the 5 MetBAG-PRS to predict the MetBAG**

| **BAG** | ***R^2^*** | **P** | **BETA** | **SE** |
| --- | --- | --- | --- | --- |
| Digestive | 0.08464422 | 0 | 1.71952623 | 0.01754702 |
| Endocrine | 0.08764909 | 0 | 1.64682324 | 0.01670874 |
| Hepatic | 0.13305813 | 0 | 1.66471788 | 0.01322075 |
| Immune | 0.11903302 | 0 | 1.65576424 | 0.01388848 |
| Metabolic | 0.05148487 | 0 | 1.65826343 | 0.02287789 |

Detailed statistics of the linear regression using the 5 MetBAG-PRSs to predict their respective MetBAG, on top of the age and sex as baseline features (null model). The alternative model included the MetBAG-PRS as an additional feature. Incremental *R^2^* values are used to quantify the prediction power.

**eFolder 1: Sensitivity analyses for the MR analyses**

Due to the large size, we share the raw figure for our extensive MR sensitivity analysis via the following Google Drive link during the peer-review process: <https://drive.google.com/drive/folders/1z8a-gw6XJbLJXGlXadcEMQiafwR0JpbL?usp=sharing>. All the data will be publicly available, together with the GWAS summary statistics, after the paper’s acceptance and publication on Synapse.
